# A lifespan transcriptomic atlas of the human prefrontal cortex at single-cell resolution

**DOI:** 10.1101/2024.11.06.24316592

**Authors:** Hui Yang, Tereza Clarence, Madeline R. Scott, Xinyi Wang, N.M. Prashant, Milos Pjanic, Sanan Venkatesh, Aram Hong, Clara Casey, Sarah R. Murphy, Alexander Kawah Yu, Zhiping Shao, Marcela Alvia, Stathis Argyriou, PsychAD Consortium, Nadejda M. Tsankova, Pavan K. Auluck, Stefano Marenco, Vahram Haroutunian, Georgios Voloudakis, Jaroslav Bendl, Colleen A. McClung, Donghoon Lee, John F. Fullard, Gabriel E. Hoffman, Kiran Girdhar, Panos Roussos

## Abstract

The dorsolateral prefrontal cortex (DLPFC) underpins higher cognitive functions and is highly susceptible to age-related decline. However, a comprehensive, lifespan-resolved map of its cellular and molecular programs has been lacking. Here, we constructed the first single-nucleus transcriptomic atlas of the human DLPFC, spanning the full lifespan, profiling over 1.3 million nuclei from 284 postmortem samples ranging in age from 0-97 years. This unprecedented resource reveals three distinct transcriptomic phases: dynamic developmental remodeling, midlife stability, and late-life molecular reactivation. Non-linear modeling of age trends uncovers ten distinct trajectories, including a neuronal resilience program peaking in early adolescence and glial aging programs marked by immune activation in late adulthood. Pseudotime analyses reconstruct lineage maturation from fetal progenitors to aged states, identifying gene modules linked to neurodevelopmental and neurodegenerative disease risk. Spatial transcriptomics confirm these dynamic programs, mapping excitatory neuron modules to specific cortical layers and glial signatures to distinct gray–white matter domains. Notably, we identify circadian reprogramming in late adulthood, with loss of neuronal core clock rhythmicity and emergence of stress-adaptive glial rhythms. Together, this study provides the first anatomically resolved, cell-type–specific, and lifespan-wide reference for the human DLPFC, establishing a foundational resource for understanding brain development, aging, and disease vulnerability.

## Main

The dorsolateral prefrontal cortex (DLPFC) plays a pivotal role in higher cognitive functions such as executive processes, decision-making, and working memory, which are critical for mental health and cognitive integrity throughout life^1–3^. However, the DLPFC is also highly susceptible to age-related decline, contributing to neuropsychiatric and neurodegenerative disorders including schizophrenia (SCZ)^4,5^, major depressive disorder (MDD)^6^, and Alzheimer’s disease (AD)^7^. Despite its clinical importance, our understanding of how the DLPFC transcriptome evolves across the human lifespan at single-cell resolution remains limited. Most existing studies have been constrained to narrow age ranges providing only snapshots of development or aging^8–11^. Moreover, many incorporate donors with neuropsychiatric or neurological conditions, making it difficult to disentangle normative aging trajectories from disease-related effects.

To address this gap, we present a single-nucleus RNA sequencing (snRNA-seq) atlas of the human DLPFC derived from over 1.3 million nuclei from 284 neurotypical samples, encompassing ages from birth to 97 years (**Fig. 1a**). This dataset, a subset of the PsychAD Consortium effort (**Supplementary Notes “PsychAD dataset”**)^12,13^, represents the largest and most age-diverse snRNA-seq resource for the human prefrontal cortex to date. Crucially, it provides a normative reference map for dissecting continuous transcriptomic trajectories across all major neuronal and glial cell types. Leveraging this resource, we conducted four integrative analyses to build a unified, cell-type-resolved framework capturing transcriptomic dynamics across the human lifespan in the DLPFC (**Fig. 1b**). First, we quantified changes in nuclei composition from development to aging. Second, we modeled gene expression trajectories to characterize transcriptional changes across distinct life stages: development, midlife, and aging. Third, we mapped age-dependent neuronal and glial lineage dynamics and integrated genetic risk for neuropsychiatric and neurodegenerative disorders, validated by spatial mapping of lineage-specific gene signatures. Finally, we investigated cell-type-specific changes in circadian rhythmicity during adulthood to capture temporal reprogramming associated with aging. Together, these analyses reveal both shared and cell type–specific mechanisms shaping DLPFC transcriptomes across lifespan and establish a foundational resource of understanding how cellular programs transition from resilience to vulnerability in human brain aging and disease.

**Figure 1.**
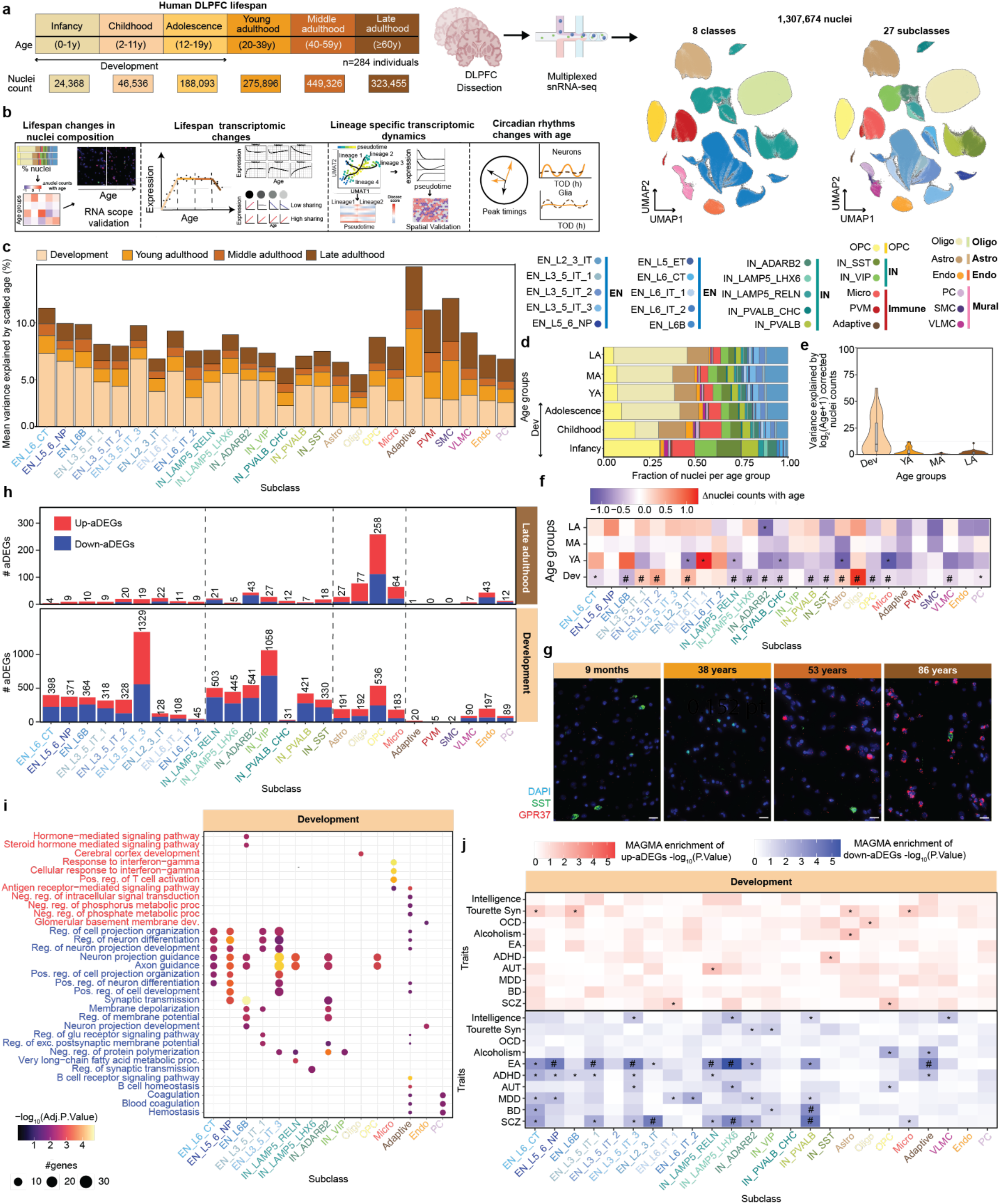
Human lifespan DLPFC transcriptomic atlas. **a**, Study design illustrating age distribution and total nuclei profiled across six age groups, grouped into four broad life stages: development (infancy, childhood, adolescence), young adulthood, middle adulthood, and late adulthood. The study is focussed on DLPFC region collected from 284 postmortem human control donors and profiled using snRNA-seq. UMAP representation of the dataset colored by eight classes and 27 defined subclasses. **b**, Overview of major data analysis conducted in the study, including lifespan changes in cell composition, gene expression, lineage-specific cellular dynamics, and age-related alterations in circadian rhythmicity. **c**, Stacked barplot showing age-associated gene expression variance across four life stages, with subclasses color-coded from beige to brown to indicate age groups. **d**, Stacked barplot representing fraction of nuclei across six age groups, colored by subclass level of taxonomy. **e**, Violin plot to show variance explained by age in nuclei abundance separately for each life stage. **f**, Estimated effect size of age-associated changes in nuclei counts across four age groups. * and # indicate nominal p-value < 0.05 and FDR < 0.05, respectively. **g**, RNAscope images showing IN_SST (green), OLIGO (red), and DAPI nuclei (blue) at four representative ages (9 months, 38 years, 53 years, and 86 years). White scale bars represent 20 μm. **h**, Counts of aDEGs colored by directionality, where red indicates genes increasing with age (Up-aDEGs) and navy indicates genes decreasing with age (Down-aDEGs), shown for development (top) and late adulthood (bottom) life stages. **i**, Functional pathway analysis of Up (in red) and Down-aDEGs (in navy) during development, highlighting subclasses significantly enriched for GO biological processes with an adjusted p-value < 0.05. **j**, Association of Up and Down-aDEGs with risk genes for a subset of brain related traits using MAGMA. * and # indicate nominal p-value < 0.05 and FDR < 0.05.

## Results

### A single nuclei RNA-seq atlas of the DLPFC across the human lifespan

To construct a lifespan-resolved transcriptomic atlas of the DLPFC at single-nucleus resolution, we profiled snRNA-seq libraries from 284 neurotypical donors (87 female and 197 male), spanning infancy (0-1 years), childhood (2-11 years), adolescence (12-19 years), and young (20-39 years), middle (40-59 years), and late adulthood (≥ 60 years) (**Fig. 1a, Supplementary Fig. 1a, Supplementary Data 1** and **Supplementary Notes “PsychAD dataset”**)^12,13^. For downstream analyses (**Fig. 1b**), infancy, childhood, and adolescence were consolidated into a single development group to increase the nucleus counts for early stage, thus yielding four broad age groups. After stringent quality control (QC) filtering^12,13^ (**Supplementary Fig. 2a** and **Methods**), we retained 1,307,674 single-nucleus transcriptomes annotated into eight major cell classes: excitatory (EN) and inhibitory (IN) neurons, astrocytes (Astro), immune cells, mural cells, endothelial cells (Endo), oligodendrocytes (Oligo) and oligodendrocyte progenitor cells (OPC) which were further resolved into 27 subclasses and 65 subtypes^12,13^(**Fig. 1a, Supplementary Fig. 1b** and **Methods**). Subclass identities were validated using canonical marker genes and alignment with external references, including the dataset from Mathys et al. 2023^14^ (**Supplementary Fig. 3**).

To quantify age-related transcriptional variation across subclasses and age groups, we applied a linear mixed model using the variancePartition^15^ tool (**Methods**) on pseudobulk gene expression aggregated at the donor-by-subclass level. This approach estimates the relative contribution of biological covariates (e.g., age, sex) and technical covariates to expression variance for each gene across all subclasses. Technical covariates in the model included a broad set of QC metrics, such as number of detected genes (n_genes), percentage of mitochondrial content (percent_mito) and other mitochondrial and ribosomal gene content metrics that were summarized per subclass and included in the model. These covariates typically accounted for < 10% of variance across most subclasses (**Supplementary Fig. 2b**) and showed minimal correlation with age (**Supplementary Fig. 2c**). Among biological factors, age explained the greatest transcriptional variance during development across all subclasses (mean: 4%), likely reflecting dynamic transcriptomic shifts associated with postnatal neurogenesis and gliogenesis^8,16^ (**Fig. 1c** and **Supplementary Fig. 2d)**. The second-largest age-associated effect was observed during late adulthood (mean: 2%) (**Supplementary Fig. 2d** and **Supplementary Data 3**), consistent with emerging transcriptional alterations linked to aging^17–19^.

We next investigated the enrichment subclass-level transcriptomes for traits relevant to brain health (**Supplementary Fig. 4** and **Methods**). Among 24 traits grouped into psychiatric, neurological and other (metabolic and immunological) categories (**Supplementary Table 2** and **Supplementary Data 2**), psychiatric traits were predominantly enriched in neuronal subclasses. MDD, autism spectrum disorder (ASD), attention deficit hyperactivity disorder (ADHD), and educational attainment also demonstrated significant enrichment in OPCs. In contrast, neurological and other traits were most strongly associated with immune, mural, and astrocytic subclasses, with immune cells (adaptive, microglia (Micro), and perivascular macrophage (PVM)) demonstrating particularly pronounced effect^20,21^. Notably, obesity showed enrichment in both EN and IN classes, highlighting a shared genetic architecture between metabolic and psychiatric traits^22,23–25^. Together, this DLPFC lifespan atlas offers a dynamic framework to decode how transcriptional programs evolve from development to late adulthood group, serving as a powerful reference for investigating the molecular underpinnings of disease risk and cellular resilience across the human lifespan.

### Age-related changes in nuclei composition across the lifespan

To investigate age-associated changes in DLPFC nuclei composition, we first examined subclass-level abundances across age groups. This revealed prominent shifts in EN and IN subclasses during infancy, childhood and adolescence (**Fig. 1d**), suggesting that compositional changes are most pronounced during development, a period marked by extensive neuronal and glial remodeling^26^. Consistent with this, quantification of variance in nuclei composition explained by age showed a peak during development (9.7%) with substantially lower variance in young, middle, and late adulthood (0.22-1.50%) (**Fig. 1e**).

We next applied the *crumblr*^*27*^ framework (**Methods**) to model nuclei abundance changes across each age group while adjusting for sex, postmortem interval, and sample source (**Supplementary Fig. 5a**). This analysis identified development and young adulthood as the most dynamic periods of compositional changes (**Fig. 1f** and **Supplementary Table 3B-E**). To further interpret this shift, we performed breakpoint analysis (**Methods**), which revealed a sharp decline in the number of subclasses showing significant changes after age 23 years, indicating a transition to relative compositional stability in DLPFC (**Supplementary Fig. 5c**). Based on this inflection point, we selected age 24 as the cutoff and modeled subclass-specific compositional changes across two groups: development (<24 years) and adulthood (≥24 years) (**Supplementary Fig. 5d**). This transition likely reflects the completion of neurodevelopmental processes and stabilization of DLPFC cellular composition. However, a subset of subclasses including IN_SST and OPC continued to exhibit significant abundance shifts in late adulthood, indicating that specific cell types remain dynamic and may be selectively vulnerable or adaptive during brain aging.

To capture trajectories across the lifespan, we modeled subclass abundances using nonlinear functions of age (**Methods** and **Supplementary Fig. 5b**). A logarithmic age model outperformed linear fits for nearly all subclasses (**Supplementary Fig. 6**). Furthermore, this analysis revealed two major patterns: log-increasing trajectories (e.g., ENs, Astro, Oligo) and log-decreasing trajectories (e.g., INs, OPCs, Micro) (**Extended Data Fig. 1a**). The observed log-shaped trends in nuclei composition align with the biology of brain maturation, wherein rapid postnatal proliferation, differentiation, and synaptic remodeling (**Extended Data Fig. 1b**) produce an accelerated rate of change during development. After accounting for cellular hierarchy, crumblr identified significant compositional shifts in 24 of 26 subclasses (**Extended Data Fig. 1c** and **Supplementary Table 3A**).

To validate these findings, we performed RNAscope assays targeting SST (marking IN_SST) and GPR37 (marking oligodendrocytes) across a representative series of four age groups, development (4 months, 9 months, and 9 years), young adulthood (38 years), middle adulthood (51 and 53 years), and late adulthood (65 and 86 years). These assays revealed a progressive decline in SST+ cells and a marked increase in GPR37+ cells with age (**Fig. 1g** and **Methods**). Quantification of the RNAscope data (**Extended Data Fig. 1d)** further confirmed the opposing lifespan trends for IN_SST and oligodendrocytes (**Extended Data Fig. 1e**), matching the log-linear model predictions.

Importantly, overlaying molecular programs on these trajectories revealed that log-increasing trends were dominated by neuronal development and synaptic programs, with strong enrichment for synaptic organization and neurogenesis-related pathways, particularly in ENs. INs showed lower enrichment magnitudes for these developmental pathways, with notable high enrichment for neurotransmitter metabolic and biosynthetic processes consistent with their specialized roles in inhibitory circuit formation. Glial subclasses in the log-decreasing trends were marked by moderate-to-high enrichment for glial proliferation, apoptotic processes, and neuroinflammatory processes (**Extended Data Fig. 1f**). Together, these findings support the existence of coordinated biological programs of cellular expansion and attrition that orchestrate the temporal remodeling of DLPFC cytoarchitecture and define discrete windows of neurodevelopmental plasticity and age-related stabilization.

**Extended Data Figure 1:**
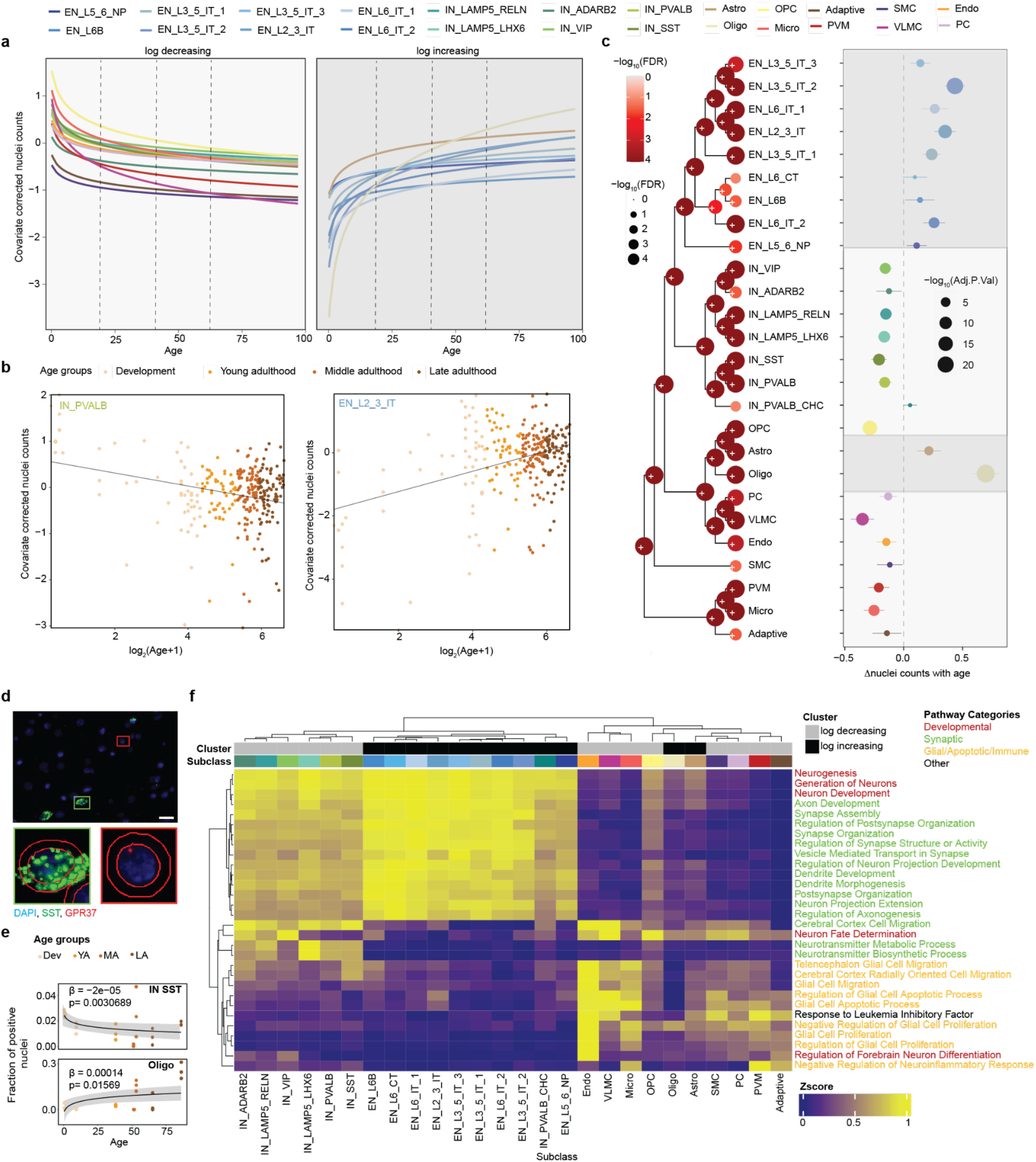
Nuclei composition changes across the lifespan. The top row displays the color scheme for each subclass. **a**, Stratification of lifespan trajectories of nuclei counts from all subclasses into logarithmic (log) increasing and decreasing groups. **b**, Examples of log-decreasing and log-increasing trends in IN PVALB (left) and EN_L2_3_IT (right) subclasses, respectively. **c**, *crumblr* results of univariate hypothesis testing on the leaves and multivariate hypothesis testing on the internal nodes shown on the hierarchical clustering, based on gene expression (left panel). Color and size of each node shows the FDR value from multivariate hypothesis testing. Right panel shows estimated effect size of age-associated changes in nuclei counts across the lifespan for all subclasses. Size of each dot depicts -log_10_ adjusted p-value from univariate testing. **d**, Representative RNAscope image showing two regions of interest from 9 months old tissue highlighted by green and red boxes. Zoomed-in views in the bottom row display segmented nuclei classified as SST+ in green (left) and GPR37+ Oligos in red (right) used for quantification. Yellow scale bar = 20 μm. **e**, Quantification of the fraction of SST+ and GPR37+ nuclei across age groups from 3 regions of interest per sample as measured by RNAscope. **f**, Heatmap showing the z-scaled pathway enrichment scores of nuclei from development groups (x-axis: subclass) and pathways (y-axis). Subclasses are annotated by cluster trends (log-increasing vs log-decreasing with age).

### Transcription remodelling of DLPFC across the lifespan

Significant age-related variance in subclass transcriptomes (**Fig. 1c)** suggested dynamic transcriptional remodeling across the human lifespan. Using dreamlet^28^ at the pseudobulk level (**Methods**), we identified three distinct transcriptional phases in the DLPFC within each age group (**Supplementary Fig. 7a-b** and **Supplemental Data 4**). The first phase, spanning development, exhibited widespread transcriptional remodeling across all subclasses, with 8,223 age-associated differentially expressed genes (aDEGs) (**Supplementary Fig. 7b**). These changes were dominated by neuronal subclasses, which accounted for 41.2% (3,389) and 40.5% (3,329) of aDEGs in EN and IN subclasses, respectively (**Fig. 1h**). The second phase, encompassing young and middle adulthood, was marked by relative transcriptomic stability, with only 27 and 1 aDEGs detected, respectively (**Supplementary Fig. 7b**). In contrast, the third phase during late adulthood showed renewed remodeling, primarily affecting glia subclasses (426/735 aDEGs) compared to neurons (246/735 aDEGs) (**Fig. 1h**). Pronounced age-associated effect sizes during development compared to late adulthood further highlighted magnitude of early-life changes (**Supplementary Fig. 7c**).

Next, to ensure adequate sensitivity for detecting aDEGs across subclasses, we performed both theoretical and empirical power assessments (**Supplementary Fig. 8a-g**). Power modeling indicated that our design with ≥50 donors per age group and ∼11,700 median expressed genes across subclasses (and minimum across age groups), is sufficiently powered to detect age-associated effects across all four age groups, confirming that the observed triphasic transcriptional architecture reflects true biological differences rather than technical or power-related artifacts.

To validate these findings, we integrated an independent replication dataset of neurotypical donors from three published sources^11,29,30^ (“Lifespan replication dataset”; **Methods** and **Supplementary Fig. 9**), totaling 306 donors spanning gestational week 22 to 101 years and 2,039,078 nuclei. For the validation analysis, three fetal donors and their nuclei were excluded to match the age range of the discovery dataset. This analysis confirmed robust replication of aDEGs across subclasses and age groups, with consistent directionality and effect sizes between datasets, particularly during development and late adulthood (high Spearman correlations and π1 statistics; **Supplementary Fig. 10**). These results support the robustness and generalizability of the triphasic model, reinforcing the biological relevance of age-related transcriptional transitions.

To interpret the functional relevance of these transcriptional changes, we performed gene set pathway analysis for the developmental aDEGs (**Supplemental Data 5)**. Downregulated genes (Down-aDEGs) were enriched for neurodevelopmental processes such as axon guidance and neuronal differentiation (**Fig. 1i**), while upregulated genes (Up-aDEGs) were associated with immune and metabolic pathways across EN_L6B, Oligo, Adaptive, Micro and Endo subclasses (**Fig. 1i**). Psychiatric trait risk genes, including those for SCZ and MDD, were significantly enriched among Down-aDEGs but not Up-aDEGs^31,32^ (**Fig. 1j** and **Supplemental Data 6**). Down-aDEGs, particularly in EN_L3_5_IT_1/3, also exhibited significantly higher intolerance to loss-of-function mutations (pLI scores^33^) compared to Up-aDEGs, suggesting stronger purifying selection (**Supplementary Fig. 7d)**.

In late adulthood, glial Up-aDEGs were enriched for processes such as leucine metabolism, DNA binding regulation, and protein localization, whereas Down-aDEGs in OPC and IN subclasses were associated with synaptic transmission and postsynaptic modulation (**Supplementary Fig. 7f** and **Supplemental Data 5**). Notably, glial and IN_ADARB2 aDEGs showed significant enrichment for AD and other neurological and immune-related traits (**Supplementary Fig. 7g** and **Supplemental Data 6**). Unlike developmental aDEGs, late adulthood aDEGs showed no significant association with pLI scores, except for OPCs (**Supplementary Fig. 7e**).

Taken together, these analyses reveal temporally segregated transcriptional programs: an early developmental phase shaped by neuronal specification and under strong evolutionary constraint; a quiescent midlife period of relative transcriptional stasis; and a late-life reactivation of glial gene networks linked to immune regulation and neurodegenerative risk. This triphasic framework establishes a molecular foundation for understanding how temporal shifts in gene expression contribute to structural maturation, cognitive function, and age-related vulnerability in the human prefrontal cortex.

### Non-linear gene expression trajectories across the lifespan of the DLPFC

After observing significant transcriptomic changes during development and late adulthood, we next examined the temporal trajectories of aDEGs to characterize their expression dynamics across the lifespan, pinpoint periods of heightened transcriptional remodeling, and identify age-associated molecular programs within individual subclasses. To model non-linear age-dependent expression patterns, we used subclass-level pseudobulk matrices encompassing 334,689 gene-subclass combinations (median=13,227 genes per subclass). and evaluated twelve alternative aging models. These included linear, log-transformed, and polynomial forms of age. As illustrated in **Fig. 2a**, we selected the optimal model based on median BIC across subclasses and extracted gene-wise model coefficients for downstream clustering and trajectory inference. The model using a second-degree polynomial of log-transformed age consistently provided the best fit, particularly in neuronal subclasses (**Supplementary Fig. 11a** and **Methods**). Importantly, power analysis confirmed that this model is sufficiently powered to detect non-linear age-associated effects across subclasses (**Supplementary Fig. 8h-j**).

**Figure 2.**
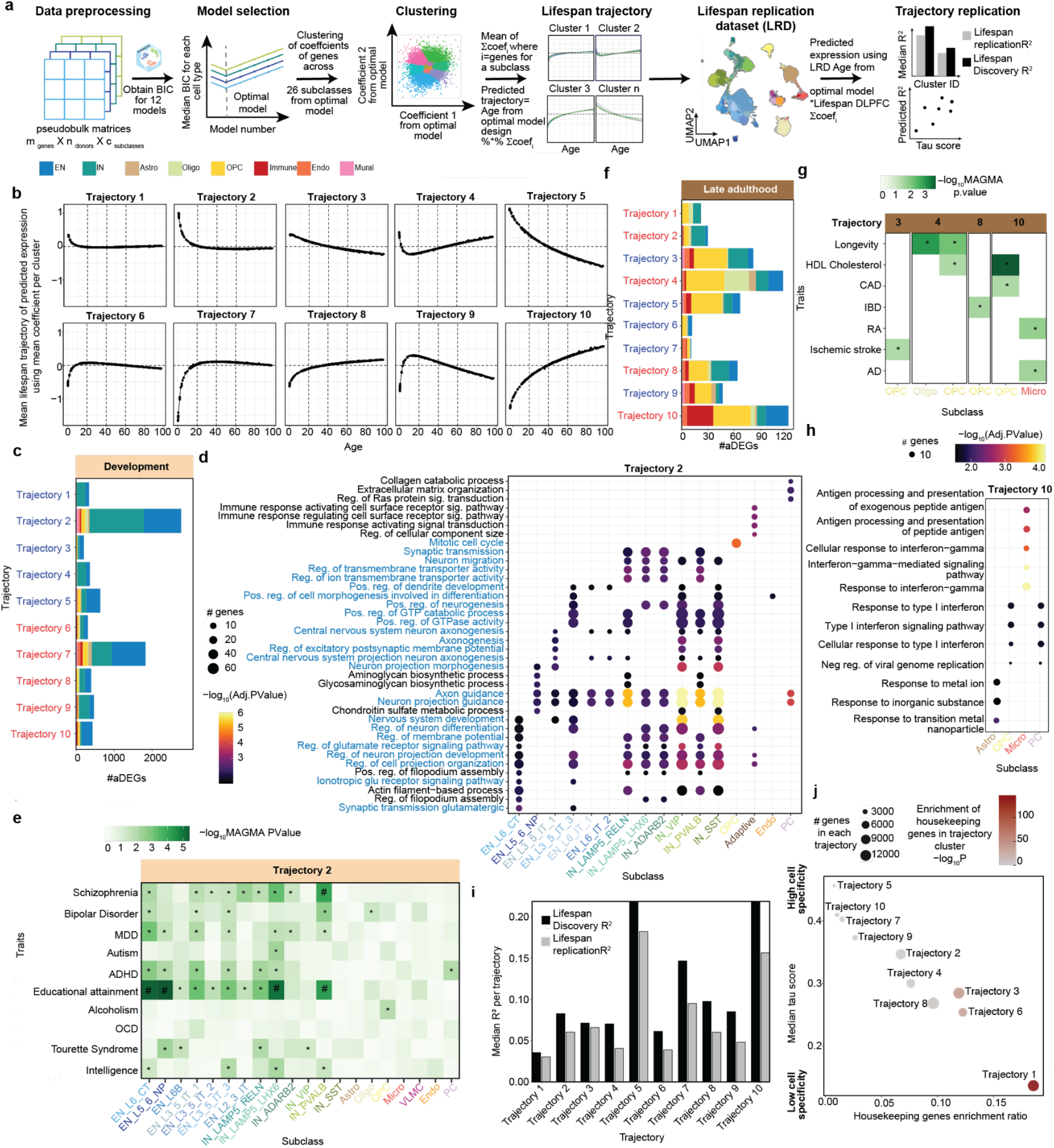
Lifespan transcriptomic trajectories. **a**, Schematic of the workflow illustrating the steps: data preprocessing, model selection for final trajectory analysis, clustering, visualization and validation of trajectories. **b**, Ten characteristic trajectories were identified, derived from the average gene expression of cellular subclasses within each cluster. The trajectories are organized by upward and downward trends. **c**,**f**, Stratification of development aDEGs (**c**) and late adulthood aDEGs (**f**) across the trajectories. Colors indicate the number of genes within each class. Trajectory names colored in red and blue indicate upregulation and downregulation, respectively, during development (**c**) and late adulthood (**f**). **d, h**, Pathway enrichment for genes from trajectory 2 and 10 highlighting subclasses significantly enriched for GO biological processes with an adjusted p-value < 0.05. Pathways in blue text in the d panel denote synaptic and glutamatergic signaling processes, which were specifically enriched in EN and IN subclasses. **e, g**, MAGMA enrichment of genes within development and late adulthood aDEGs highlighting their relevance to neuropsychiatric and late age diseases. * and # indicate nominal p-value < 0.05 and FDR < 0.05 from MAGMA enrichment. **i**, Bar plot showing the predicted median R^2^ of aging trajectories across all genes within each cluster, separately for the lifespan DLPFC discovery (black) and replication (gray) datasets **j**, Scatter plot displaying the the ratio of housekeeping genes in each trajectory, and median tau score (cell specificity) per trajectory. Point size corresponds to the number of genes in the trajectory, and color indicates fisher test enrichment (–log_10_P value) for housekeeping genes in each trajectory cluster.

Approximately 40.3% (135,120/334,689) of these gene-subclass combinations showed significant age-related changes, which clustered into 10 distinct lifespan trajectories (**Fig. 2b, Supplementary Fig. 11b-d, Supplementary Fig. 12a,b** and **Supplemental Data 7**). Functional enrichment analysis revealed a spectrum of biological processes, ranging from basic cellular functions to brain-specific synaptic activities (**Supplemental Data 8**). For example, Trajectory 1 (16,188 genes from 23 subclasses) highlighted basic cellular processes, while Trajectory 5 (5,172 genes from 26 subclasses) was enriched for Wnt signaling, extracellular matrix organization, and synaptic transmission, with contributions from both glial and neuronal subclasses (**Supplementary Fig. 13a**). These findings reveal divergent molecular programs associated with age-related dynamics across the lifespan.

Mapping development aDEGs onto these trajectories showed that EN and INs genes were predominantly represented in Trajectories 2 and 7 (**Fig. 2c**). Notably, Trajectory 2, containing most downregulated developmental aDEGs, was enriched for neuronal pathways related to synaptic signaling, glutamatergic transmission, and central nervous system development (**Fig. 2d**). Genes in Trajectory 2 (**Fig. 2e**) specifically showed significant overlap with brain-related GWAS traits, including schizophrenia, bipolar disorder, and educational attainment compared to genes in other trajectories (**Supplementary Fig. 12c** and **Supplemental Data 9**). Remarkably, most of the development aDEGs (67.6% in Trajectories 1, 2, 6, and 7) peaked around adolescence (∼12-13 years) and remained stable thereafter, suggesting that early-life transcriptomic changes in neurons establishes a foundation for later brain function and psychiatric risk.

Late adulthood aDEGs were concentrated in Trajectories 3–5 and 8–10 (**Fig. 2f**), which exhibited three distinct patterns: (a) pronounced changes during development, followed by midlife stabilization and an onset of changes during late adulthood (Trajectories 3, 8), (b) progressive change across lifespan (Trajectories 5, 10), and (c) reversal trends after early peaks (Trajectories 4, 9) (**Supplementary Fig. 13b** and **Supplemental Data 9**). Trajectories with upward trends, particularly Trajectories 4, 8, and 10, were enriched for risk genes associated with neurological and immune-related disorders (**Fig. 2g**). Trajectory 10 was notable for enrichment of subclass-specific immune activation and interferon signaling pathways (**Fig. 2h**), highlighting glial immune responses in late adulthood neuroinflammation. Microglial genes within trajectory 10 were further enriched for processes such as protein localization to the plasma membrane and calcium homeostasis (**Supplementary Fig. 14a,b**) and showed significant overlap with AD risk genes (**Supplementary Fig. 14c**). Interestingly, microglial Trajectories 3 (chromatin modification) and 6 (autophagy), both characterized by age-related decline, also showed AD risk gene enrichment, suggesting additional microglial programs may contribute to AD vulnerability even in the absence of statistically significant late-life differential expression (**Supplementary Fig. 14d** and **Supplemental Data 10**). Among neuronal late adulthood aDEGs, only the IN_ADARB2 subclass showed significant pathway enrichment, with genes assigned to Trajectory 2 exhibiting a downregulation pattern during development and late adulthood (**Supplementary Fig. 14e, f** and **Supplemental Data 10**), suggesting diminished synaptic plasticity, a hallmark of neuronal aging^34,35^.

To assess the reproducibility of gene expression trajectories, we applied the nonlinear age coefficients derived from the Lifespan DLPFC dataset to predict pseudobulk gene expression in the independent Lifespan replication dataset based on donor age (**Methods**). This cross-dataset prediction yielded high concordance between predicted and observed expression values, as quantified by R^2^ (**Fig. 2i** and **Supplementary Fig. 15)**. Among the modeled trajectories, Trajectories 5 and 10 exhibited the highest R^2^ values in both discovery and replication datasets, indicating strong robustness and generalizability of these age-associated expression patterns.

We next examined which biological features distinguished reproducible trajectories. A comparison of median R^2^ values (**Fig. 2i**) with cell-type specificity (tau scores) and housekeeping gene enrichment (**Fig. 2j**), revealed that the most reproducible trajectories (e.g., Trajectories 5 and 10) were also the most cell-type specific and the least enriched for housekeeping genes. In contrast, Trajectory 1 showed the lowest reproducibility and was most enriched for housekeeping genes while being least cell-type specific. These findings suggest that lifespan transcriptional programs that are restricted to specific cell types are more consistently recapitulated across individuals and datasets, whereas trajectories dominated by housekeeping genes may capture greater inter-individual variability. Together, these analyses highlight the robustness of specific non-linear lifespan trajectories, particularly those anchored in cell-type-specific programs and point to a potential role for housekeeping gene programs in mediating individual-level variability in DLPFC.

### Transcriptional convergence in excitatory neurons emerges with age in the DLPFC

While transcriptomic convergence with age has been described in bulk human brain tissue^36^ and in mouse models^37^, its cell type-specific dynamics within the human DLPFC remain poorly characterized. To address this, we leveraged our comprehensive lifespan dataset to systematically assess gene expression convergence from development through late adulthood across ENs, INs and glial subclasses. To quantify convergence, we applied the *mashr*^*38*^ framework, an empirical Bayes approach that models shared effect sizes across conditions. For each age group, we estimated composite probabilities (P_E_, P_I_, P_G_) summarizing the degree of shared age-associated transcriptional changes across 9 EN, 7 IN, and 4 glial subclasses, respectively (**Extended Data Fig. 2a**). Analyses focused on genes with nominally significant age effects (from dreamlet; **Supplementary Fig. 16a**) in at least two subclasses (**Methods** and **Supplemental Data 11**).

We observed a marked increase in gene sharing with age, particularly in EN, where P_E_ > 0.9 rose substantially between development and late adulthood (**Extended Data Fig. 2b**). Similar but less pronounced trends were detected in IN and glial subclasses (**Supplementary Fig. 17a**). To better understand this trend, we examined genes at the extremes of the sharing spectrum. Low sharing genes (P_E_ < 0.01), such as *KCNH4* (P_E_ = 0), exhibited divergent age-associated effect sizes (**Supplementary Fig. 17b)**, whereas highly shared genes like *PARP2* (P_E_ = 0.99) showed consistent expression changes across all EN subclasses (**Extended Data Fig. 2c**).

When averaging composite probabilities (P_E_, P_I_, P_G_) across age groups, we observed a clear increase in EN convergence from development through late adulthood (**Extended Data Fig. 2d**), with more moderate changes in IN and glia (**Supplementary Fig. 17c** and **Supplementary Fig. 18a,b**). Highly shared genes (P_E_, P_I_, P_G_ > 0.9) exhibited significantly lower tau scores^39^ than low-sharing genes, indicating reduced cell type specificity (**Supplementary Fig. 19, Supplementary Fig. 18c** and **Supplementary Table 4**). This suggests that transcriptional convergence during aging may preferentially involve genes with ubiquitous roles in cellular maintenance rather than highly cell-specific programs. Functional enrichment analyses revealed that shared genes in EN and IN during development and young adulthood were enriched for neuronal adhesion and neurotransmitter transport (**Supplemental Data 12**). In contrast, shared genes in late adulthood were enriched for canonical aging processes, including DNA repair in EN, protein degradation via ubiquitination in IN, and antigen processing and immune surveillance in glia (**Extended Data Fig. 2e**).

To probe molecular mechanisms underlying convergence, we constructed protein-protein interaction (PPI) networks using STRING-db^40^ (**Methods**). In young adulthood, EN networks highlighted mitochondrial and axonal projection genes, while IN networks featured synaptic proteins (**Supplemental Data 13**). By late adulthood, EN and IN networks revealed clusters involved in DNA repair, RNA transcription, and RNA processing, whereas glial networks showed clusters related to apoptosis, MHC class I signaling, and RNA splicing (**Extended Data Fig. 2f–h, Supplementary Fig. 20**). Notably, most genes in these networks were upregulated with age, consistent with increased demands for genomic stability and immune surveillance during aging. Together, our findings reveal a transition from divergent developmental programs to shared aging molecular programs in human DLPFC, with transcriptional convergence most pronounced in excitatory neurons.

**Extended Data Figure 2.**
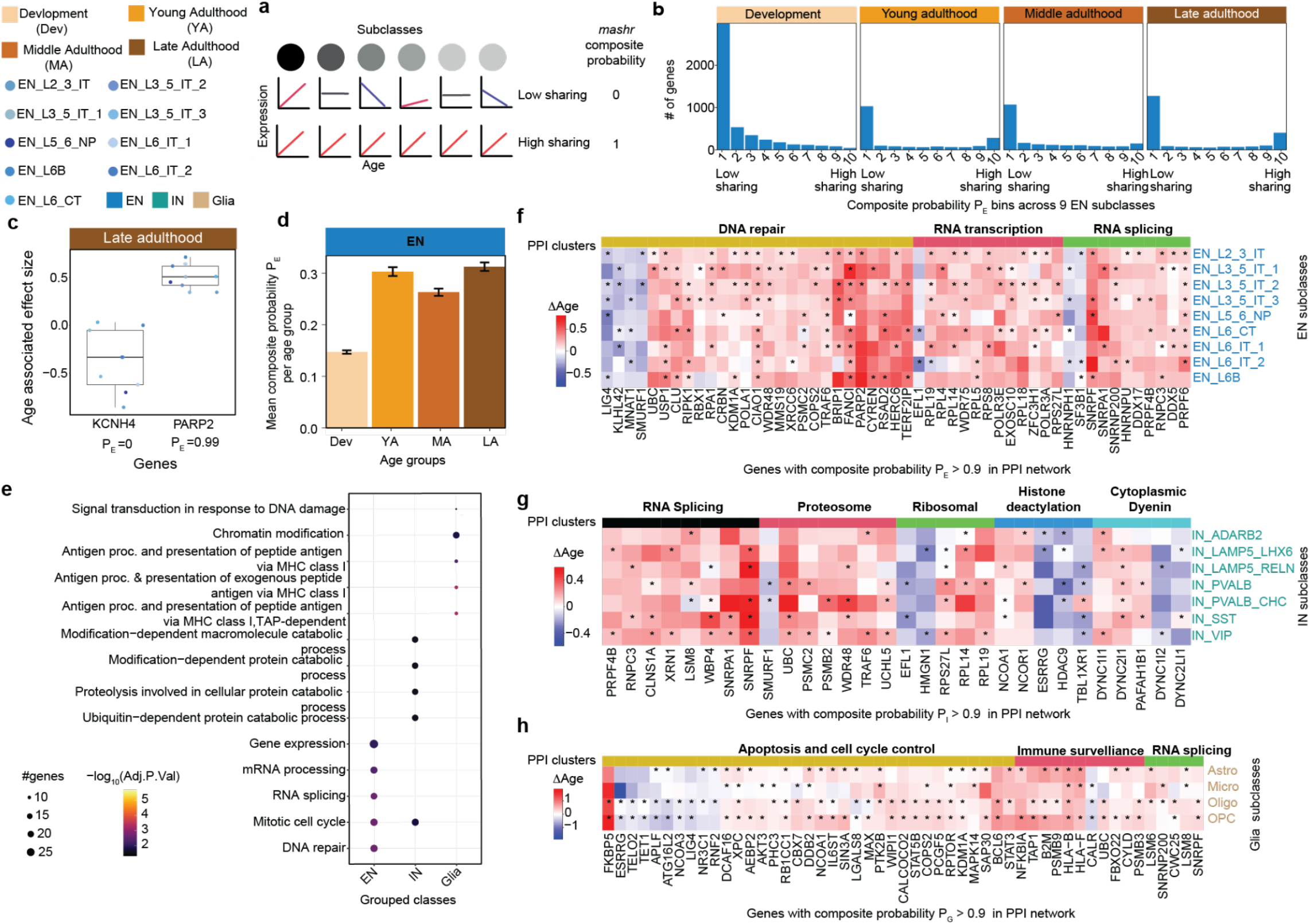
Degree of sharing in age effect sizes and associated biological mechanisms across cell subclasses. The top-left panel displays the color gradient and abbreviations for age group, subclasses and classes: development (Dev), young adulthood (YA), middle adulthood (MA), late adulthood (LA), 9 EN subclasses and EN, IN and glia. **a**, A schematic illustrating low degree sharing with heterogeneous age effect sizes (composite probability = 0) and high degree sharing with concordant effect sizes across subclasses (composite probability = 1) **b**, Number of genes stratified into 10 equally sized bins based on composite probability (P_E_) values for EN, ranging from 0 to 1, from development to late adulthood. **c**, Distribution of late adulthood age-associated effect sizes for KCNH4 and PARP2 genes with P_E_ = 0 and P_E_ = 0.99, respectively, across EN subclasses. **d**, Mean P_E_ of genes in each age group (4,767 Dev, 1,926 YA, 2,092 MA, 2,271 LA). This plot is the mean of probabilities of genes shown in the b panel. **e**, Biological mechanisms of shared genes during late adulthood across 9 EN, 7 IN and 4 glia that show significant enrichment for GO terms after an adjusted p-value < 0.05. **f-h**, Heatmap of age-associated effect sizes of 51, 30 and 51 shared genes across 9 EN, 7 IN and 4 glia classes which showed significant PPI interactions (score > 0.9) and had at least 5 genes within the PPI network. Color bar on the top of the heatmap shows associated mechanisms obtained from k-means clustering of genes within the PPI network. “*” denotes genes that are nominally significant with p-value < 0.05 from age groups analysis using *dreamlet*.

#### Cellular dynamics of astrocytes across the lifespan

Recognizing the variance observed during development and late adulthood (**Fig. 1c** and **Extended Data Fig. 2b**), we performed pseudotime trajectory analysis to delineate cellular dynamics of DLPFC lineages across the lifespan. To capture early developmental transitions, we integrated our dataset with published snRNA-seq spanning gestation to adulthood^11^, resulting in a combined dataset of 1,454,617 nuclei from 311 individuals (**Supplementary Fig. 21a-d**, see **Methods**). We utilized UMAP of MATuration (UMAT)^11^, which restricts neighbor selection to adjacent stages, thereby enhancing the precision of representing transitional processes across the lifespan.

In astrocytes, we identified two distinct age-related patterns (**Fig. 3a**) corresponding to fibrous astrocytes (FA; high *GFAP* expression) and protoplasmic astrocytes (PA; low *GFAP*, high *SLC1A2*) (**Extended Data Fig. 3a**). Pseudotime trajectories revealed that PA matured later than FA (**Fig. 3b,c**), reflecting their distinct microenvironments: FA in white matter interacts with myelinated axons and oligodendrocytes, while PA in gray matter engages with neurons and synapses^41,42^. Notably, PA exhibited consistently higher single cell disease relevance scores (scDRS) for migraines across pseudotime, especially during maturation and aging (**Fig. 3d**). These findings underscore the significance of cellular interactions and temporal dynamics in understanding astrocytes functions and their role in disease susceptibility.

**Figure 3.**
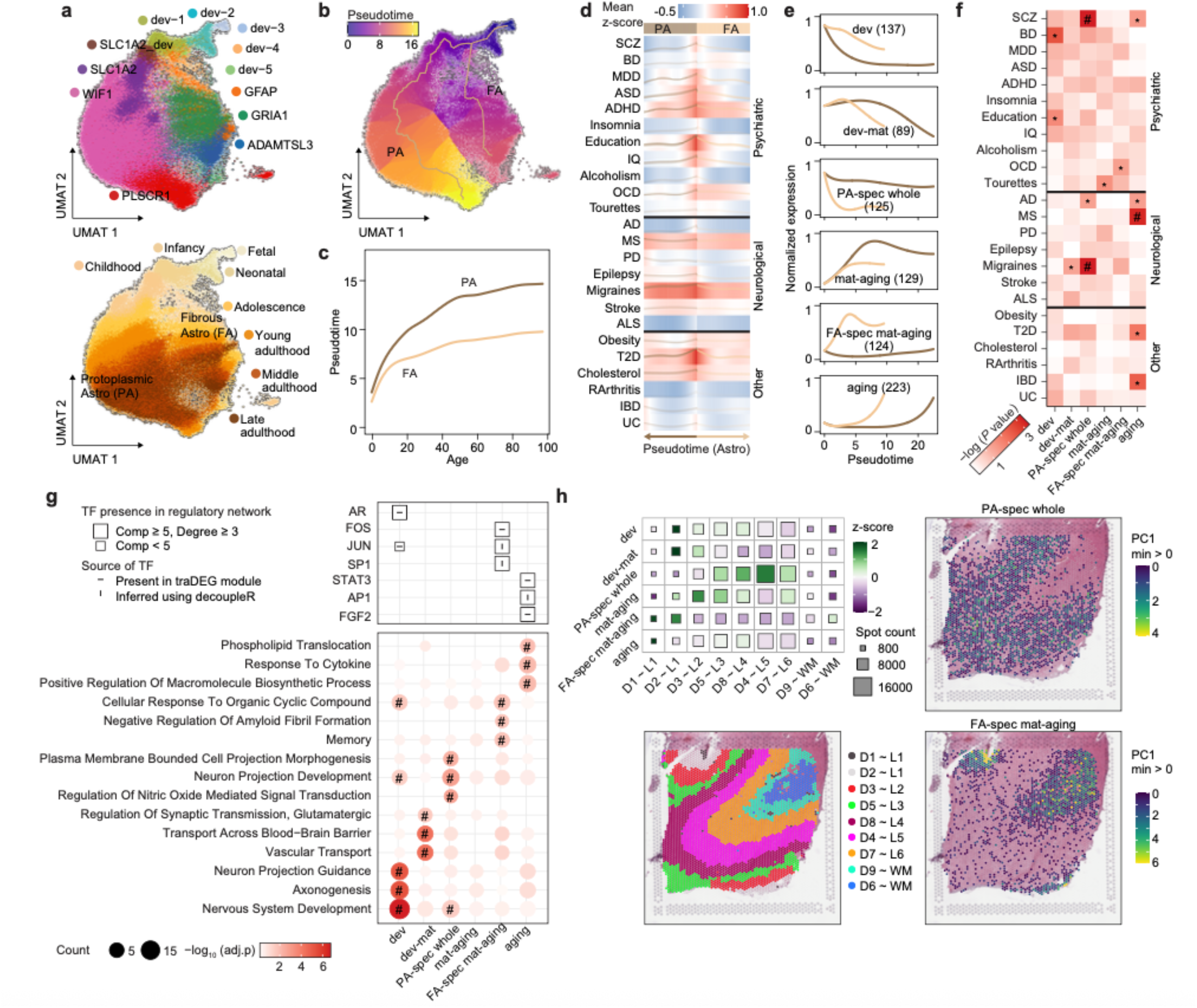
Cellular dynamics of astrocytes lineages. **a,b**, UMAT representation of astrocyte lineage colored by subtype (**a**, upper), stage (**a**, bottom) and pseudotime (**b**). **c**, Maturation rates of two astrocyte categories. **d**, Fitted disease scores along the pseudotime trajectory for the two astrocyte categories (**Methods**). Traits that were significantly enriched are shown in bold and visualized in detail using the line plots below. **e**, Scaled (0-1) expression of differentially expressed genes along trajectories (traDEGs) in the astrocyte lineage clustered into six modules. Fibrous astrocytes (FA) are indicated in light brown and protoplasmic astrocytes (PA) are indicated in dark brown. **f**, Enrichment adjusted *P* value of different classes of GWAS traits corresponding to traDEG clusters in Astro lineage. *: p*-*value < 0.05, #: adjusted p*-*value < 0.05. **g**, Enriched GO terms corresponding to traDEG clusters in the astrocyte lineage, with TF presence and network properties indicated (top). Squares represent TFs, with size reflecting network properties, and internal symbols denoting source. Circles (bottom) represent enriched GO terms, with size proportional to gene count and color indicating enrichment significance: #: adjusted p-value < 0.05. **h**, Multi-gene spatial analysis of traDEG modules. z-score heatmap (upper left) show expression across 10 DLPFC samples, while cortical layer annotation (bottom left) and PC1 score maps for PA (upper right) and FA (bottom right) modules are displayed for a representative DLPFC sample (Br2743_mid), highlighting region-specific patterns.

To resolve molecular programs shaping astrocyte trajectories, we identified 827 differentially expressed genes along pseudotime trajectories (traDEGs; FDR < 0.05, Moran’s I ≥ 0.05) clustered into six modules reflecting developmental (dev), mature (mat), and aging processes (**Fig. 3e, Extended Data Fig. 3b, Supplemental Data 14**, and **Methods**). The dev module was enriched for psychiatric traits including bipolar disorder and education attainment (**Fig. 3f, Methods** and **Supplemental Data 15**), and functional analysis revealed involvement in nervous system development and axonogenesis (**Fig. 3g** and **Supplemental Data 16**), emphasizing astrocytic contributions to cortical development and mental health^43–45^. In contrast, the aging module was enriched for neurological and immunological traits and linked to macromolecule biosynthesis and cytokine response, implicating astrocytic programs in neuroinflammation and late-life vulnerability (**Fig. 3f,g**)^46–48^. The PA-specific (spec) module showed strong enrichment for SCZ and migraine risk genes (**Fig. 3f**), consistent with scDRS results (**Fig. 3d**), suggesting functional interactions between neurons and PA may contribute to disease etiology^29,49^. Moreover, FA-spec mat-aging module was linked to memory, negative regulation of amyloid fibril formation (**Fig. 3g**), and cellular response to organic cyclic compounds, underscoring their involvement in cognitive support, neuroprotection, and cellular defense mechanisms^50^.

To elucidate regulatory mechanisms shaping these astrocytic programs, we inferred transcription factor (TF) activity for each traDEG module using the univariate linear model (ULM) in decoupleR^51^ (**Methods**). Candidate TFs were integrated with module genes into PPI networks constructed with STRING-db, and prioritized based on their presence in connected components (size ≥ 5) and high node degree (≥ 3). Among astrocytic modules, robust TF-gene networks were identified for the dev, FA-spec mat-aging and aging modules (**Extended Data Fig. 3c**), reflecting stage- and lineage-specific regulatory architectures. For example, AR emerged as a key developmental regulator, coordinating a network linked to protein folding (**Fig. 3g** and **Extended Data Fig. 3c**). Validation against enhancer-driven regulons (eRegulon) from SCENIC+ in the developing neocortex^52^ further supported AR’s role, along with additional regulators identified in astrocytic modules (**Extended Data Fig. 3d,e**). These findings reveal dynamic, stage-specific TF networks that coordinate astrocytic contributions to brain development and disease vulnerability.

To validate the anatomical relevance of pseudotime modules, we analyzed Visium spatial transcriptomics data from human DLPFC^53^ (**Methods**). This approach allowed us to assess whether distinct pseudotime modules exhibit laminar or region-specific enrichment, thereby linking temporal transcriptional trajectories to spatially organized astrocyte functions *in situ*. PA-specific genes were broadly distributed across cortical layers (**Fig. 3h** and **Extended Data Fig. 3f**), aligning with neuronal support functions and psychiatric trait associations (**Fig. 3f,g**). By contrast, FA-specific genes were enriched in white matter (**Fig. 3h** and **Extended Data Fig. 3f**), consistent with neuroprotective roles (**Fig. 3g**). Together, these findings reveal temporally and spatially resolved astrocytic programs, underscoring subtype- and stage-specific roles in shaping brain health and disease vulnerability across the human lifespan.

**Extended Data Figure 3.**
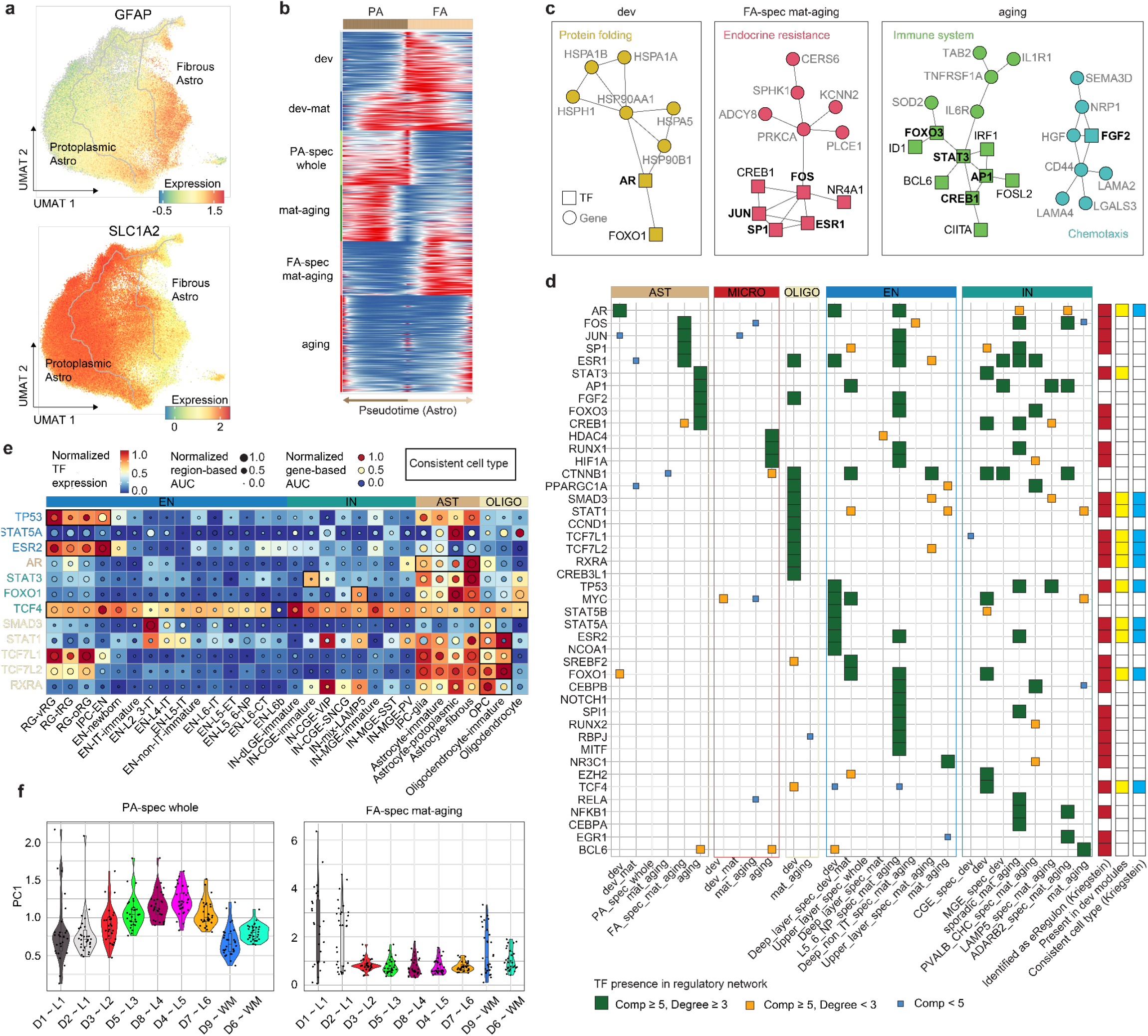
Characteristics of astrocytes. **a**, Expression of fibrous (GFAP) and protoplasmic (SLC1A2) astrocyte markers. **b**, Fitted expression of traDEG clusters along the pseudotime trajectory for the two astrocyte categories. **c**, TF-gene regulatory networks inferred for Astro traDEG modules, with enriched biological processes annotated based on network composition. Squares indicate TFs; circles indicate target genes. TF labels are shown in black; gene labels are shown in gray. Bolded TF names denote key regulators with high connectivity (connected components ≥ 5 nodes and degree > 3). **d**, Key TFs in regulatory networks for each traDEG module, with square size and color reflecting network properties (green: component size ≥ 5, degree ≥ 3; orange: component size ≥ 5, degree < 3; blue: component size < 5). Right columns show external validation: red marks eRegulon in the developing human neocortex^52^, yellow indicates presence in developmental traDEG modules, and blue highlights high expression of these TFs in consistent cell types in the developing human neocortex^52^. **e**, The normalized TF expression levels, region-based AUC scores, and gene-based AUC scores of TFs present in developmental traDEG modules across cell types. Font color denotes the dominant cell type assignment of each TF. **f**, Violin plots of PC1 scores of PA-spec whole (left) and FA-spec mat-aging (right) modules across DLPFC samples, indicating variability across cortical layers.

#### Cellular dynamics of microglia and oligodendrocytes across the lifespan

We next examined the lifespan dynamics of other glial lineages, focusing on microglia and oligodendrocytes. For Micro, pseudotime trajectory analysis revealed a single developmental path (**Fig. 4a** and **Extended Data Fig. 4a**). Trait enrichment analysis showed significant and stable enrichment in several neurological traits, including AD and multiple sclerosis, as well as immunological traits such as rheumatoid arthritis, inflammatory bowel disease, and ulcerative colitis (**Extended Data Fig. 4b**). Similar to astrocytes, developmental genes in Micro were associated with psychiatric traits and neuronal development processes, while aging genes were linked to immunological functions (**Fig. 4b-d, g, Extended Data Fig. 4c** and **Supplemental Data 14-16**). These findings highlight the dual roles of microglia in supporting brain development and contributing to neurodegeneration and immune-related conditions across the lifespan^54,55^. Notably, the progression from developmental to aging modules in Micro revealed genetic associations with AD that emerge later in life, offering insights beyond those identified in previous GWAS studies focused on aged donors^56^ (**Fig. 4b-d**).

**Figure 4.**
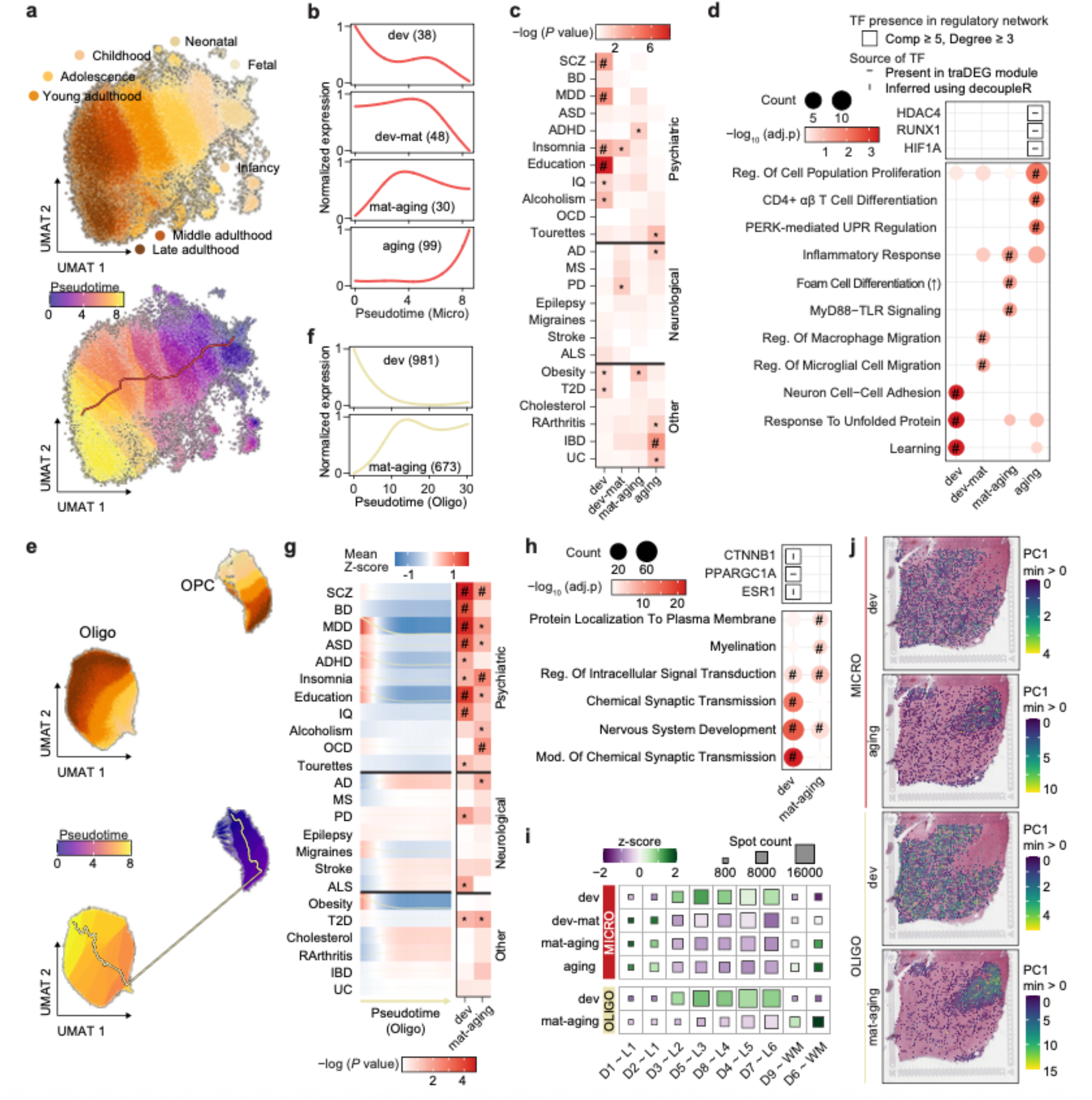
Cellular dynamics of Micro and Oligo. **a,e**, UMAT representation of the Micro (**a**) and Oligo (**e**) lineages, colored by stage (upper) and pseudotime (bottom). **b,f**, Clusters of traDEG modules in Micro (**b**) and Oligo (**f**) lineages. **c**, Enrichment adjusted *P* value of different classes of GWAS traits corresponding to traDEG clusters in Micro lineage. *: p-value < 0.05, #: adjusted p-value < 0.05. **d,h**, Enriched GO terms corresponding to traDEG clusters in the Micro (**d**) and Oligo (**h**) lineage, with TF presence and network properties indicated (top). Squares represent TFs, with size reflecting network properties, and internal symbols denoting source. Circles (bottom) represent enriched GO terms, with size proportional to gene count and color indicating enrichment significance: #: adjusted p-value < 0.05. **g**, Disease association along Oligo pseudotime trajectories (left) and traDEG modules (right). *: p-value < 0.05, #: adjusted p-value < 0.05. **i,j**, Multi-gene spatial analysis of Micro and Oligo traDEG modules. z-score heatmap shows expression across 10 DLPFC samples (**i**), while PC1 score maps for selected modules are displayed for a representative DLPFC sample (**j**, Br2743_mid), highlighting region-specific patterns.

For the Oligo lineage, we identified a trajectory from OPC to mature Oligos (**Fig. 4e-h** and **Extended Data Fig. 4a**). Trait enrichment along this trajectory revealed distinct transitions: OPC showed significant associations with psychiatric traits and obesity, while mature Oligos exhibited enrichment for AD (**Fig. 4g**). Additionally, developmental genes in Oligos were associated with multiple psychiatric disorders and synaptic transmission, whereas aging genes were linked to myelination, protein localization to plasma membrane, and obsessive compulsive disorder (**Fig. 4f-h** and **Supplemental Data 14-16**). These patterns highlight the stage-specific functions of Oligos in maintaining neural circuitry and their roles in lifespan vulnerability to psychiatric and neurological traits.

TF-gene network analysis identified CTNNB1, PPARGC1A and ESR1 as key regulators within the Oligo developmental module, orchestrating networks tied to synaptic signaling and axon development (**Fig. 4h** and **Supplementary Fig. 22**). In Micro, the aging module featured HDAC4, RUNX1 and HIF1A-centered networks associated with immune response pathways (**Fig. 4d** and **Extended Data Fig. 4d**). Spatial transcriptomic validation revealed that developmental modules in both lineages were broadly distributed across cortical layers, while aging modules were enriched in white matter regions (**Fig. 4i, j** and **Extended Data Fig. 4e**). Together, these findings underscore the temporally and spatially distinct regulatory programs of Micro and Oligo, linking their lineage-specific dynamics to lifespan disease susceptibility.

**Extended Data Figure 4.**
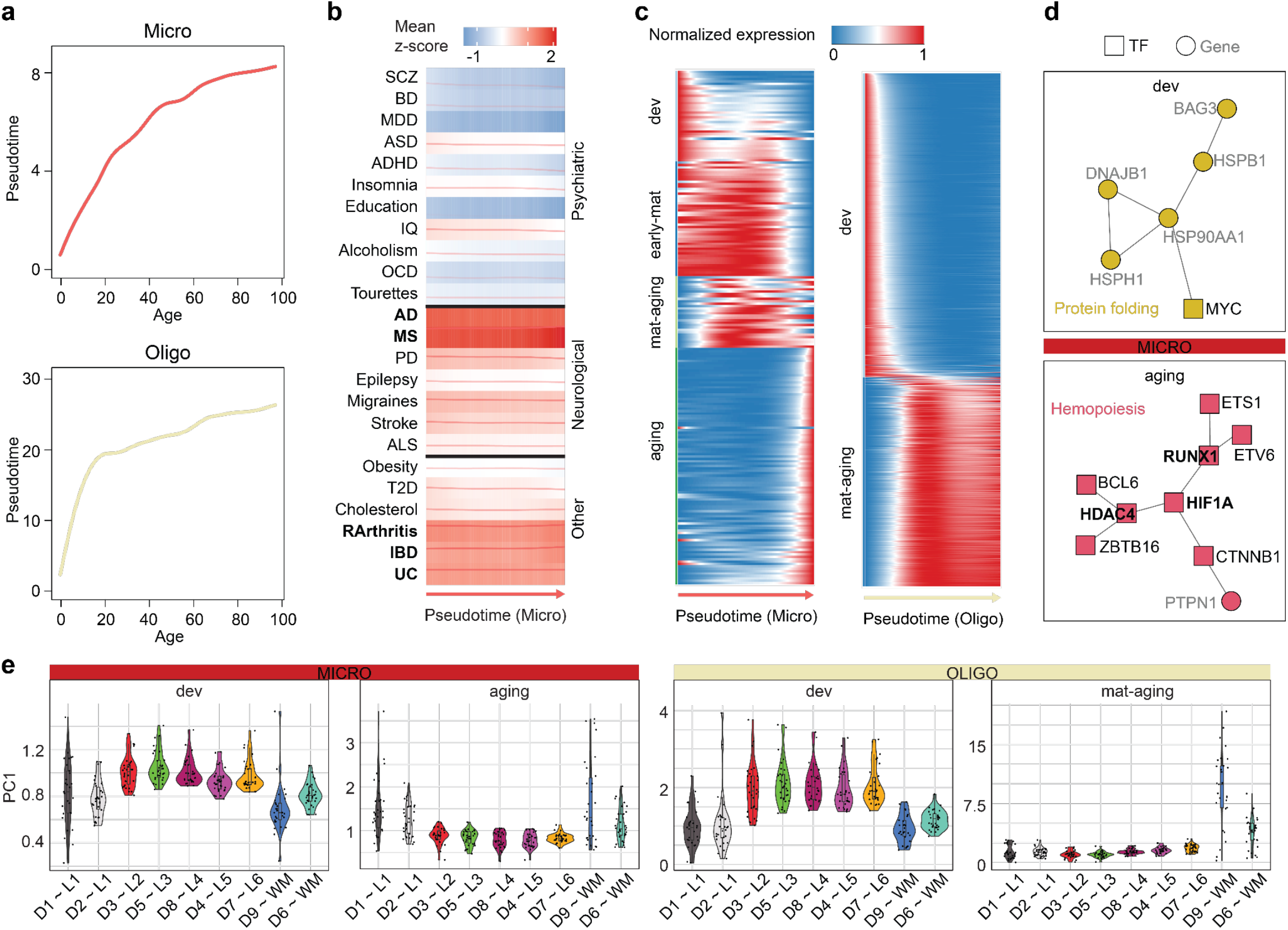
Characteristics of Micro and Oligo lineages. **a**, Maturation rates of the Micro (upper) and Oligo (bottom) lineages. **b**, Fitted disease scores along the pseudotime trajectories for Micro lineage. **c**, Fitted expression of traDEG clusters along the pseudotime trajectory for the Micro (left) and Oligo (right) lineages. **d**, TF-gene regulatory networks inferred for Micro traDEG modules, with enriched biological processes annotated based on network composition. Squares indicate TFs; circles indicate target genes. TF labels are shown in black; gene labels are shown in gray. Bolded TF names denote key regulators with high connectivity (connected components ≥ 5 nodes and degree > 3). **e**, Violin plots of PC1 scores of selected traDEG modules in Micro (left) and Oligo (right) lineages across DLPFC samples, indicating variability across gray and white matter.

### Cellular dynamics of neuronal lineages across the lifespan

We next focused on neuronal lineages, including excitatory and inhibitory neurons (**Fig. 5** and **Extended Data Fig. 5**). For ENs, pseudotime trajectory analysis revealed nine developmental paths grouped into three categories based on cortical distribution: upper-layer intratelencephalic projection neurons (Upper-IT), deep-layer intratelencephalic projection neurons (Deep-IT), and deep-layer non-intratelencephalic projection neurons (Deep-non-IT) (**Fig. 5a** and **Extended Data Fig. 5a**). These categories displayed distinct temporal patterns, with Deep-non-IT ENs maturing earliest, followed by Deep-IT and Upper-IT ENs (**Fig. 5a,b** and **Extended Data Fig. 5b**), consistent with the canonical inside-out pattern of cortical development^57,58^.

**Figure 5.**
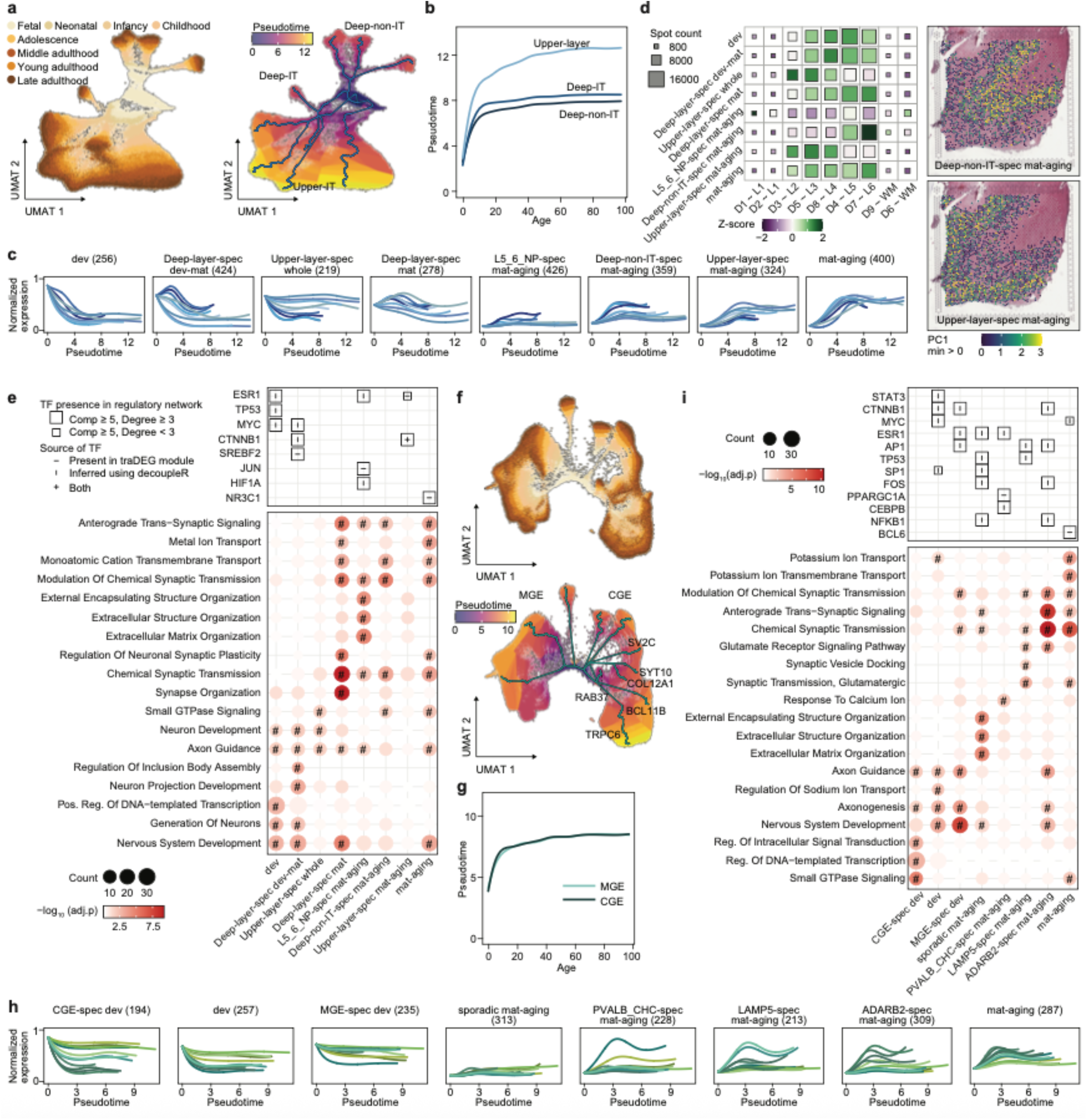
Cellular dynamics of neuronal lineages. **a**,UMAT representation of EN lineage colored by stage (left) and pseudotime (right). **b**, Maturation rates of three EN categories. **c**, Scaled (0-1) expression of the traDEGs in EN lineage clustered into eight modules. **d**, Multi-gene spatial analysis of EN traDEG modules. z-score heatmap (left) show expression across 10 DLPFC samples, and PC1 score maps for Deep-non-IT-spec mat-aging (upper right) and upper-layer-spec mat-aging (bottom right) modules are displayed for a representative DLPFC sample (Br2743_mid), highlighting cortical layer-specific patterns. **e,i**, Enriched GO terms corresponding to traDEG modules in EN (**e**) and IN (**i**) lineages, with TF presence and network properties indicated (top). Squares represent TFs, with size reflecting network properties, and internal symbols denoting source. Circles (bottom) represent enriched GO terms, with size proportional to gene count and color indicating enrichment significance: #: adjusted p-value < 0.05. **f**, UMAT representation of IN lineage, colored by stage (upper) and pseudotime (bottom). **g**, Maturation rates of two IN categories. **h**, Clusters of traDEGs in IN lineage clustered into eight patterns.

Across these trajectories, we identified 2,686 genes exhibiting dynamic expression, which clustered into eight distinct modules (**Fig. 5c, Extended Data Fig. 5c** and **Supplementary Data 14**). Spatial transcriptomics revealed striking laminar enrichment of these modules: upper-layer-specific modules were localized predominantly to cortical layers II-IV, while deep-layer-specific modules were enriched in layers V-VI (**Fig. 5d** and **Extended Data Fig. 5d**). This laminar segregation underscores the spatial distinction of transcriptional programs across EN subclasses and highlights their developmental divergence and functional specializations within cortical circuits.

Functional enrichment analysis revealed that developmental genes were associated with neuron generation and development, while mature and aging genes were enriched for synapse transmission and ion transport (**Fig. 5e** and **Supplementary Data 15**), essential for synaptic function and structural integrity throughout life^59,60^. Deep-layer modules included developmental and maturation programs enriched in neuron projection development and protein folding, with cholesterol metabolism uniquely regulated by SREBF2-centered networks (**Fig. 5e** and **Supplementary Fig. 23**). Aging modules encompassed processes such as extracellular matrix organization and circadian entrainment, with key regulators including NR3C1 and NCOR2. Across upper- and deep-layer modules, shared regulators such as CTNNB1, MYC, and ESR1 orchestrated these programs, reflecting both common and trajectory-specific transcriptional control. Disease enrichment analysis revealed robust associations between excitatory neuron and psychiatric traits (**Extended Data Fig. 5e** and **Supplementary Data 16**). Schizophrenia, for example, showed significant associations during developmental and mature stages, suggesting critical windows of vulnerability^32,61–63^. Tourette’s syndrome and obesity exhibited progressive associations during EN maturation, indicating roles for neuronal development in these traits. These results extend previous findings that primarily emphasized IN involvement^64,65^. Together, these findings underscore how trajectory-dependent transcriptional programs shape excitatory neuron development, synaptic maintenance, and disease associations.

For INs, pseudotime analysis identified eleven trajectories, grouped into two categories based on origin: medial ganglionic eminence (MGE)-derived and caudal ganglionic eminence (CGE)-derived INs (**Fig. 5f** and **Extended Data Fig. 5f**). While overall maturation patterns of CGE- and MGE-derived INs were similar, CGE-derived INs exhibited greater variability across trajectories (**Fig. 5g** and **Extended Data Fig. 5g**), suggesting distinct temporal dynamics across these IN categories. We identified 2,036 traDEGs forming eight distinct modules (**Fig. 5h, Extended Data Fig. 5h**, and **Supplementary Data 14**). Developmental modules were enriched for genes associated with nervous system development, axonogenesis and axon guidance (**Fig. 5i** and **Supplementary Data 15)**. Additionally, CGE-specific developmental genes were involved in regulation of intracellular signal transduction. Mature and aging genes were enriched in synaptic transmission and ion transport, contributing to the stability and adaptability of neural circuits throughout life^59,60^. Specifically, sporadic trajectories in mature and aging modules showed enrichment for extracellular matrix maintenance, echoing patterns observed in L5_6_NP ENs (**Fig. 5i**). TF-gene network analysis revealed modular architectures with distinct functional enrichments, reflecting stage- and subtype-specific regulation (**Fig. 5i** and **Supplementary Fig. 24**). Furthermore, enrichment in psychiatric disorders was also observed for inhibitory neuron traDEGs (**Extended Data Fig. 5i** and **Supplementary Data 16**), with the enrichment being more pronounced in developmental genes. Notably, PVALB_CHC-specific mature and aging genes showed significant enrichment in alcoholism and stroke, potentially due to their involvement in response to calcium ions (**Fig. 5i**). Together, these findings reveal distinct temporal dynamics and disease associations within inhibitory neuron lineages, underscoring their contributions to both developmental and late-life brain disorders.

**Extended Data Figure 5.**
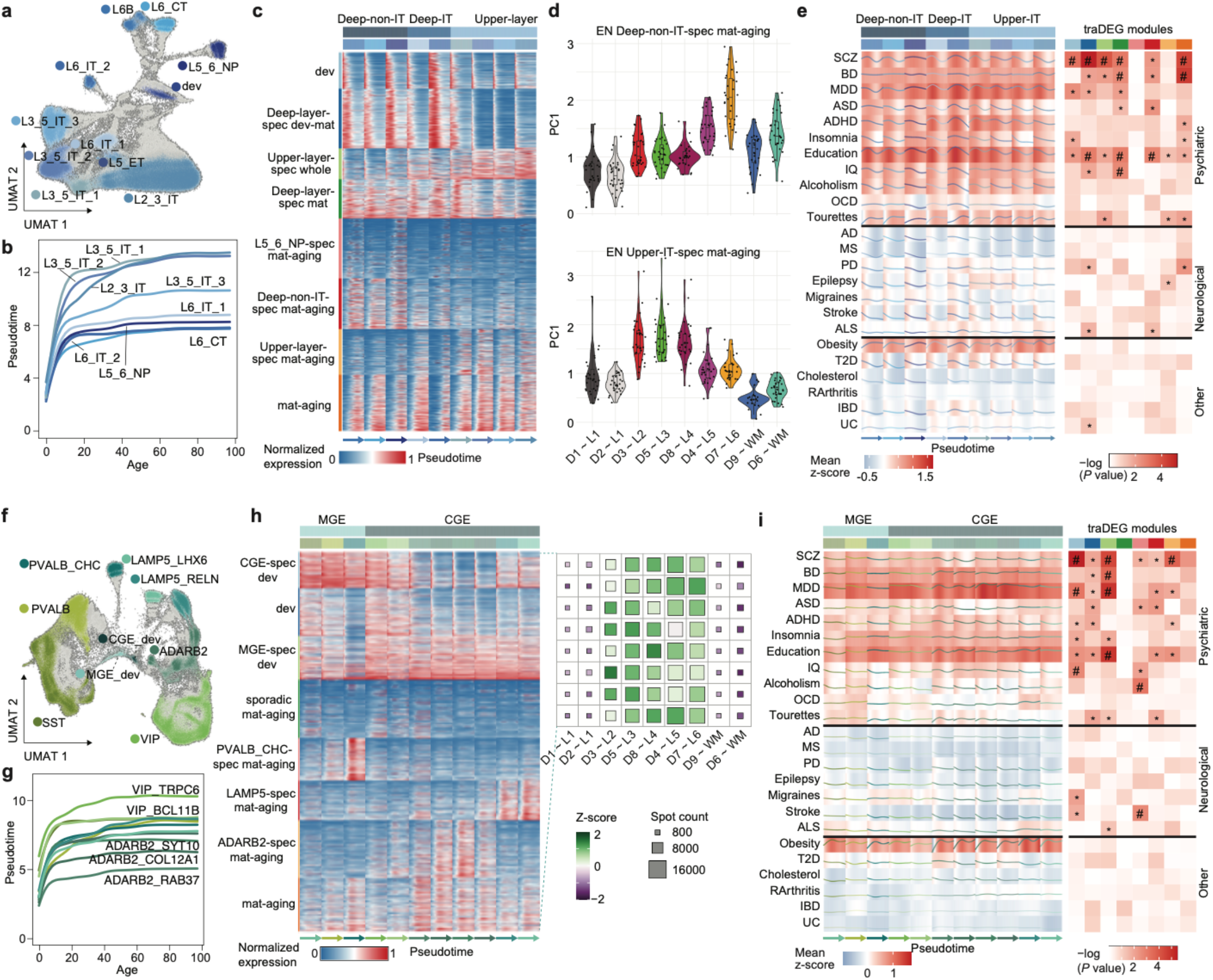
Cellular dynamics of EN and IN lineages. **a,f**, UMAT representation of the EN (**a**) and IN (**f**) lineages colored by subclass. **b,g**, Maturation rates of nine EN (**b**) and eleven IN (**g**) trajectories. **c**, Fitted expression of traDEG clusters along the pseudotime trajectory for the EN lineage. **d**, Violin plots of PC1 scores of Deep-non-IT-spec mat-aging (upper) and Upper-layer-spec mat-aging (bottom) EN traDEG modules across DLPFC samples, indicating variability across cortical layers. **e,i**, Disease association along EN (**e**) and IN (**i**) pseudotime trajectories (left) and traDEG modules (right). *: p-value < 0.05, #: adjusted p-value < 0.05. traDEG modules are colored by identity, as in panel **c** (EN) and **h** (IN). **h**, Fitted expression of traDEG clusters along the pseudotime trajectory for the IN lineage.

#### Circadian reprogramming in the late adulthood DLPFC transcriptome

Given the enrichment of transcription factor networks for circadian entrainment pathways and the established link between circadian rhythm disruption and aging^66,67^, we investigated how age influences circadian transcriptional programs in the DLPFC. Using a cosinor model, we identified 24-hour gene expression rhythms in subjects with known times of death (TOD) (**Fig. 6a, Extended Data Fig. 6** and **Methods**). This allowed us to compare rhythmicity in young and middle adulthood (YA+MA, n = 119) to late adulthood (LA, *n* = 77) (**Fig. 6a**).

**Figure 6.**
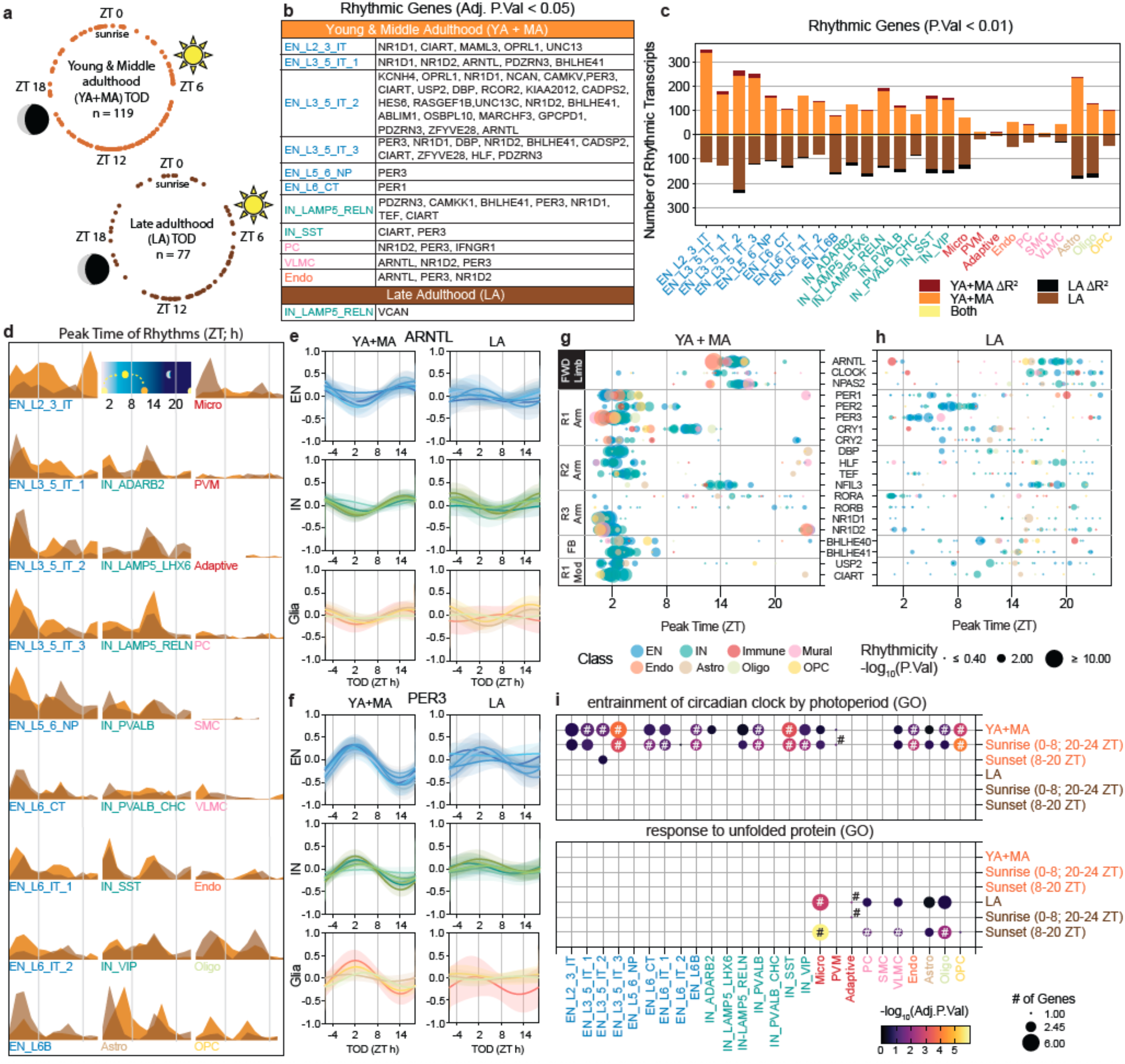
Circadian reprogramming in late adulthood. **a**, Time of death (TOD) information for subjects used in the rhythmicity analysis. Each dot is an individual’s TOD in zeitgeiber time (ZT). For full 24 h coverage, Young and Middle adulthood groups were combined (YA+MA; 21 - 60 years), and then compared to the Late adulthood (LA; ≥ 61 years) group. **b**, Genes identified as rhythmic (FDR < 0.05). **c**, Number of genes identified as rhythmic (p < 0.01) for YA+MA, LA or Both groups. Rhythmic genes with significantly different R^2^ values (p < 0.01) are denoted as dark red or black. **d**, Peak expression times for all rhythmic genes. **e,f**, Example rhythms for ARNTL (**e**) and PER3 (**f**), two canonical molecular clock genes, in EN, IN, and glia, including microglia, astrocytes, oligodendrocytes and OPC subclasses. Solid lines are the calculated 24 h oscillation, transparent area is the 95% confidence interval. Individual plots are available in **Supplementary Fig. 27-28. g,h**, Rhythmicity and timing of genes associated with the circadian molecular clock in YA+MA (**g**) and LA (**h**). Represented pathways include the forward limb (FWD limb), the primary PER/CRY regulatory arm (R1), the secondary D-Box regulatory arm (R2), the secondary ROR/NR1D regulatory arm (R3), the BHLHE40/41 feedback loop (FB), and modifiers of the R1 arm. (R1 mod). **i**, Important pathways identified by enrichR. # represents FDR < 0.05.

In YA+MA, we detected 64 significant rhythmic genes (FDR < 0.05) across 11 subclasses, with 69% (44/64) found in upper-layer IT ENs (**Fig. 6b** and **Supplementary Data 17**). Many of these genes are associated with the circadian molecular clock (**Extended Data Fig. 6b**). By contrast, only *VCAN* exhibited significant rhythmicity in LA (**Fig. 6b**). Permutation testing confirmed the robustness of these rhythmic signatures (empirical p-value 0.0099, **Supplementary Fig. 25**, and **Supplementary Data 18**). Applying a relaxed threshold (p-value < 0.01) revealed more rhythmic genes in YA+MA than LA across most subclasses, except for EN_L6_CT, EN_L6B, IN_LAMP5_LHX6, Micro, and Oligo, where rhythmic genes were abundant in LA (**Fig. 6c** and **Extended Data Fig. 6c-d**). Comparative analyses showed few overlapping rhythmic genes between age groups within each subclass, indicating that the identity of transcripts with rhythmic expression shifts substantially from YA+MA to LA (**Fig. 6c** and **Supplementary Data 17**). Rhythmic genes peaked either shortly after sunrise (∼2 ZT) or in the evening (∼12-14 ZT), a bimodal pattern observed across subclasses and age groups (**Fig. 6d**). Of the few genes rhythmic in both age groups, 49% (29/59) showed significantly different peak times (FDR < 0.05; **Supplementary Data 19**), emphasizing age-related reprogramming of rhythmic expression. These findings indicate widespread circadian reprogramming during late adulthood, with both the identity and timing of rhythmic transcripts altered.

In YA+MA neuronal subclasses, core clock genes displayed synchronized peak expression: forward limb genes peaked at night, while regulatory arms peaked in the morning (**Fig. 6e-h, Extended Data Fig. 6b,e**, and **Supplementary Data 17**). In LA, these rhythms were largely lost, and peak times became inconsistent across subclasses (**Fig. 6e-h, Extended Data Fig 6d**, and **Supplementary Data 20**). Pathway enrichment analyses mirrored these patterns, with circadian clock pathways highly enriched in YA+MA but absent in LA (**Fig. 6i, Supplementary Fig. 26** and **Supplementary Data 21**). Interestingly, Micro and Oligo in LA showed enrichment for Response to Unfolded Protein, suggesting adaptive transcriptional programs in response to aging-related stress (**Fig 6i** and **Supplementary Fig. 26**). Further analysis of gene peak timing revealed that sunrise-peaking genes in YA+MA were enriched for circadian pathways, while sunset-peaking genes in LA were associated with response to unfolded protein in Micro, PC, VLMC, and Oligo (**Fig 6i** and **Supplementary Data 21**). Together, these results demonstrate that while circadian transcriptional signatures are robust in young and middle adulthood neuronal subclasses, they are largely lost in late adulthood, with glial subclasses acquiring novel rhythmic programs associated with cellular stress responses during aging^68^.

**Extended Data Figure 6.**
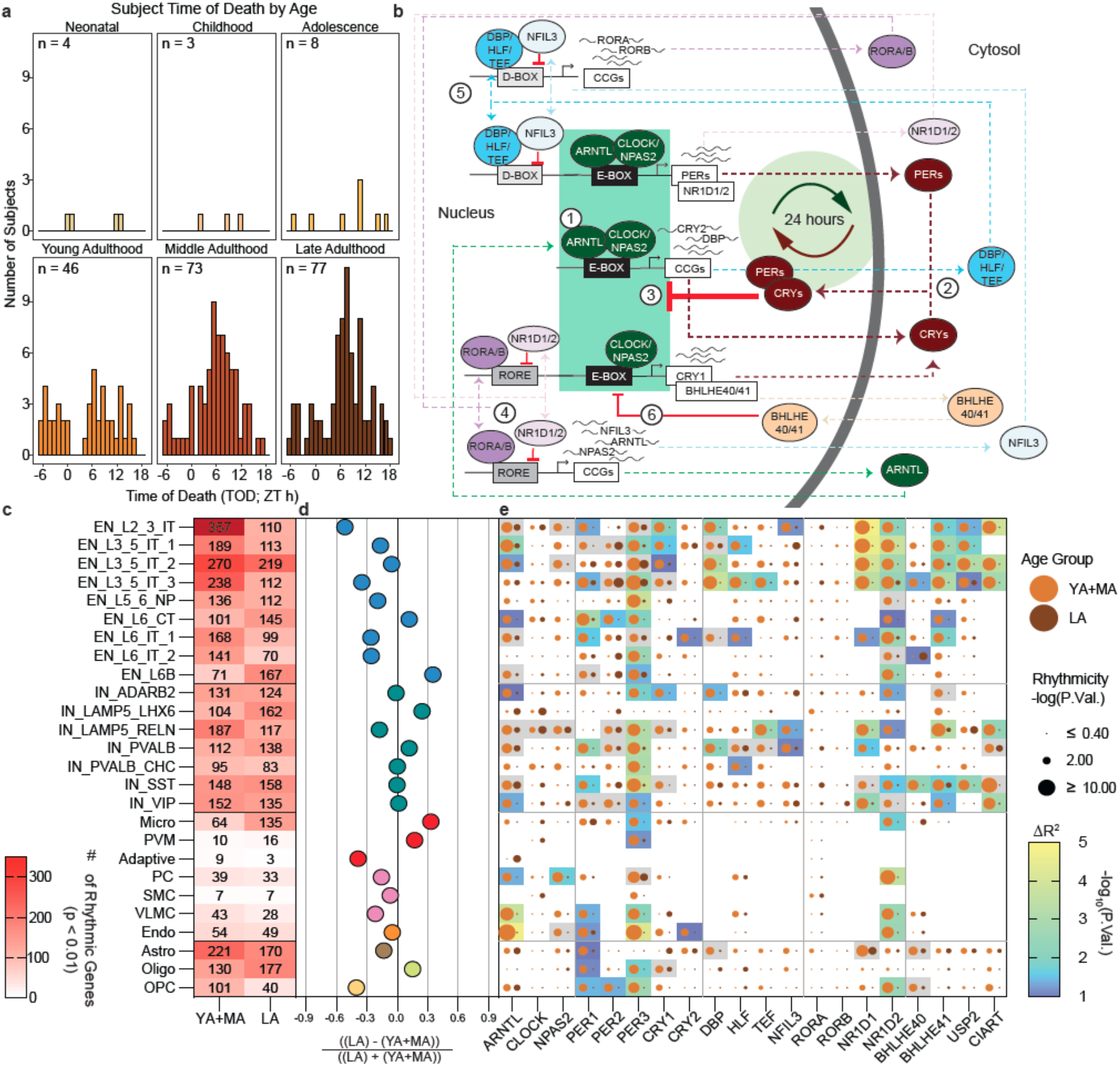
Age associated changes in 24 h gene expression rhythms. **a**, Time of Death (TOD) distributions within each age group. **b**, The core of the molecular clock is a forward limb (1) that drives 3 major regulatory arms (2/3, 4, 5) in an interconnected series of transcription-translation feedback loops (reviewed in^69^). **c**, Number of rhythmic genes (p < 0.01) in each subclass in YA+MA and LA. **d**, Percent difference in the number of rhythmic genes within each subclass between YA+MA and LA. **e**, Comparing rhythmicity (size) of circadian clock genes between YA+MA and LA. Additionally, the results of testing for difference in rhythmicity (ΔR^2^) is included for genes that were significantly rhythmic (p < 0.01) in at least one group.

## Discussion

We present the first cell-type–resolved, lifespan-spanning single-nucleus transcriptomic atlas of the human DLPFC, profiling over 1.3 million nuclei from 284 neurotypical donors aged 0 to 97 years. This resource captures dynamic, cell-type-specific gene expression changes across the lifespan and provides a normative reference for interrogating molecular deviations in brain disorders.

Our atlas reveals that DLPFC nuclei composition follows a global non-linear trajectory across the lifespan, with the most pronounced changes occurring during early development, a period marked by gliogenesis, synaptic remodeling, and apoptosis, followed by stabilization in adulthood. Age 24 emerged as an inflection point, after which the composition of most neuronal and glial subclasses remained stable, with the exception of IN_SST and OPCs, which continued to exhibit age-associated changes. This pattern suggests that the major wave of compositional changes is during early development, while aging induces selective and limited shifts restricted to specific subclasses. Importantly, these non-linear trends were validated by RNAscope, which confirmed a logarithmic decline in IN_SST and an increase in oligodendrocyte abundance across the lifespan.

Our analyses delineate three distinct phases of transcriptomic change: robust developmental remodeling, midlife stability, and late-life reactivation of molecular programs. During development, 2.9% and 1.7% of aDEGs occurred in neuronal and glial subclasses, respectively. These changes stabilized after age 20 but significantly resurged after age 60, predominantly affecting glial subclasses compared to neurons. While previous studies have noted neuronal vulnerability during development^70^ and glial susceptibility with aging^71,72^, our findings quantitatively compared the aging landscape across cells and demonstrated neurons are transcriptionally more resilient to aging. Strikingly, aging modules across glia—microglia, astrocytes, and oligodendrocytes— converged on stress-response and immune pathways, suggesting a coordinated glial reprogramming in late life.

By quantifying transcriptomic similarity across cell types, we uncovered patterns of divergence during development reflecting tightly regulated neurodevelopmental processes^73^ that contrasted with increasing convergence in late adulthood. This convergence was most pronounced in excitatory neurons and enriched for pathways involved in DNA repair and RNA splicing, potentially reflecting shared mechanisms of resilience in post-mitotic cells ^74–76^. To our knowledge, this is the first demonstration of cell subclass–specific transcriptional divergence during development and convergence in late adulthood across the human DLPFC.

To capture global patterns of transcriptomic change beyond discrete age-group comparisons, we modeled continuous gene expression dynamics across the lifespan and uncovered ten distinct non-linear trajectories spanning all major DLPFC cell types (**Fig. 2**). Notably, Trajectory 2, a neuronal resilience program, was characterized by early downregulation of genes enriched for neurodevelopmental disorder risk, peaking around ages 12–13 and remaining stable thereafter. This trajectory accounted for 67.6% of developmentally regulated genes and aligns with normative brain growth patterns observed in MRI studies^77^, suggesting that perturbations prior to this critical window may have long-term impacts on brain function^78^. In contrast, Trajectory 10 captured a glial aging program defined by late-life upregulation of genes involved in immune function and the unfolded protein response, highlighting distinct temporal windows of transcriptional vulnerability and resilience across cell types.

Pseudotime analyses across glial and neuronal lineages uncovered dynamic transcriptional programs and disease association maps. Psychiatric trait-linked genes showed persistent expression in neuronal lineages across lifespan (**Extended Data Fig. 5e,i**), indicating roles in both early development and adult brain maintenance^70,77,79^. These genes are also highly expressed during the development of astrocyte and oligodendrocyte lineages (**Fig. 3d** and **Fig. 4g**), highlighting the crucial glial-neuronal interplay necessary for proper brain function^29^. Importantly, our analysis reveals that convergence is most pronounced in excitatory neurons during late adulthood and is associated with aging-related mechanisms such as DNA repair and RNA splicing to preserve integrity in post-mitotic cells. In contrast, neurodegenerative disease-associated genes were highly expressed in microglia and oligodendrocyte during aging (**Fig. 4c,g**), suggesting the evolving functions of glia from developmental support to late-life homeostasis and immune responses^14,55^. The emergence of these late-life glial programs—converging across subclasses—implicates shared aging mechanisms underlying diverse neurodegenerative conditions.

The lineage- and stage-specific programs were further validated using spatial transcriptomics: excitatory neuron modules localized to expected cortical layers, while glial (microglia and oligodendrocytes) modules exhibited gray-to-white matter enrichment during aging **(Fig. 4i,j** and **Fig. 5d**), supporting predicted compartmental transitions of glial subtypes. These spatial patterns reinforce the anatomical relevance of pseudotime-inferred programs and highlight region-specific vulnerability. Together, these integrated analyses provide the first anatomically resolved, lifespan-wide map of human DLPFC gene regulatory programs, offering critical insights into temporal windows of cellular vulnerability and resilience.

Circadian rhythm analysis uncovered a striking reprogramming of molecular rhythms in late adulthood (**Fig. 6**). While rhythmic expression of core clock genes was robust in young and middle adulthood, it diminished in neurons during late adulthood. Conversely, microglia and oligodendrocytes gained rhythmicity in unfolded protein response pathways, suggesting an adaptive response to age-related stress^66,67,80^. These findings provide a cell-type–resolved perspective on how circadian disruption in aging may impact sleep and cognition. To our knowledge, this presents the first single-cell characterization of circadian gene expression across adult stages of the human DLPFC.

Despite these advances, our study is limited by its reliance on transcriptomic data alone, which does not capture post-transcriptional, proteomic, or epigenomic regulation. A promising avenue for future research is to integrate this atlas with orthogonal modalities such as proteomics, epigenomics, and multi-region spatial transcriptomics to elucidate how transcriptional aging trajectories translate to functional and structural changes in the brain. Future integration with regulatory and functional genomic layers may also clarify how resilience mechanisms in development relate to vulnerability in aging.

In conclusion, this lifespan-resolved atlas provides a foundation for understanding how cellular programs transition from resilience to vulnerability. Our findings highlight the transcriptional stability of neurons across adulthood, the late-life reprogramming of glia, and the interplay between aging, circadian rhythm disruption, and disease risk architecture. The coordinated temporal and spatial organization of these programs suggests that distinct inflection points during development and aging may represent windows for therapeutic intervention. This work offers a critical resource for the community and opens avenues for targeted interventions aimed at preserving brain health and mitigating age-related cognitive decline and neurodegeneration.

## Methods

### DLPFC lifespan study design

Brain tissue specimens were obtained from NIMH-IRP Human Brain Collection Core (HBCC): 172 samples, ages 0.2-85 years and The Mount Sinai NIH Neurobiobank (MSSM): 112 samples, ages 20-97 years. In total, 284 neurotypical controls of age range 0-97 years from “PsychAD dataset” were included in this study (**Supplementary Fig. 1a**). The majority of the samples are from European descent (*n* = 158) followed by African (*n* = 95), American (*n* = 26), East Asian and South Asian (n=5). **Supplementary Fig. 1a** shows the demographic information at donor level, including sex, age, time of death and ancestry, stratified by corresponding brain bank. The data was categorized into four groups: 1) developmental which constitutes infancy (0-1 year), childhood (2-11 years), adolescence (12-19 years), 2) young (20-39 years), 3) middle (40-59 years) and 4) late adulthood (≥ 60 years) samples.

We utilized the available neuropathology details on MSSM samples for control samples selection. The selection criterion for MSSM included the following:

1. CERAD scores: For neuritic plaque density of MSSM samples, only those with a score of 1 (no AD) were included.
2. Braak stage: Samples with Braak stages 0, 1, or 2 were retained.
3. Secondary diagnoses: Donors with any additional brain-related diagnosis, including neurodegenerative (e.g. AD and PD) and neuropsychiatric diseases (e.g. SCZ and BD) as well as the presence of mild cognitive impairment, were not kept.

In principle, we applied equivalent selection criteria to the HBCC samples. Although the HBCC cohort did not provide specific Braak and CERAD values, we confirmed through review of neuropathological reports that the selected donors did not exhibit significant plaque or tangle pathology. Thus, all selected donors who lacked brain-related diagnoses were deemed reliable neurotypical controls, despite the absence of detailed neuropathological data.

The link to the complete demographic and clinical information of the present study population is provided in **Supplemental Table 1** and **Supplemental Data 1** and at synapse platform **syn52396927**.

#### Fluorescence-activated nuclear sorting (FANS) protocol and snRNA-seq hashing from frozen brain tissue

All libraries from the “PsychAD” dataset were prepared using a standardized protocol for nuclei isolation and hashing^32^. The lifespan dataset was generated after completing snRNA-seq preprocessing and the taxonomy step, as described in this and succeeding sections. The process involved isolating and sorting nuclei from frozen brain specimens using fluorescence-activated nuclear sorting (FANS). 25 mg of frozen postmortem human brain tissue was homogenized in a cold lysis buffer with RNAse inhibitors. The homogenate was filtered through a 40 µm cell strainer, and the flow-through was underlaid with sucrose solution before centrifugation at 107,000 xg for 1 hour at 4 °C. The resulting pellets were resuspended in PBS with 0.5% bovine serum albumin (BSA). Six samples were processed simultaneously, with up to 2 million nuclei per sample pelleted at 500 xg for 5 minutes at 4° C. The nuclei were then resuspended in 100 µl staining buffer and incubated with 1 µg of a unique TotalSeq-A nuclear hashing antibody (BioLegend) for 30 minutes at 4 °C. Prior to FANS, the volumes were adjusted to 250 µl with PBS, and 7-Aminoactinomycin D (7-AAD) was added according to the manufacturer’s instructions. The 7-AAD positive nuclei were sorted into tubes pre-coated with 5% BSA using a FACSAria flow cytometer (BD Biosciences).

After FANS, the nuclei were washed twice with 200 µl of staining buffer, resuspended in PBS and quantified using the Countess II (Life Technologies). The concentrations were adjusted, and equal volumes of differentially hash-tagged nuclei were combined. Using 10x Genomics single cell 3’ v3.1 reagents, 60,000 nuclei (10,000 per donor) were processed in each of two lanes to create technical replicates. During cDNA amplification for library preparation, 1 µl of a 2 µm HTO cDNA PCR additive primer^81^ was included. The supernatant from a 0.6x SPRI selection was reserved for HTO library generation. Both cDNA and HTO libraries were prepared according to manufacturer’s instructions (10x Genomics and BioLegend, respectively). Sequencing was performed at the NYGC using the Novaseq platform (Illumina).

#### Preprocessing of snRNA-seq dataset

The DLPFC lifespan cohort analyzed in this study is a subset of the broader “PsychAD dataset,” which comprises over 6 million nuclei and was defined in a separate manuscript (Lee et al., 2024, Fullard et al., 2024).^12,13^ Below, we summarize the computational processing and initial quality control (QC) applied to the original PsychAD dataset.

##### Computational Processing and Demultiplexing

Sequencing reads from all pools of multiplexed samples were aligned to the hg38 reference genome using *STARsolo*^*82*^. To assign cells from each pool to their respective donors, a genotype-based demultiplexing pipeline was employed, followed by genotype concordance checks. Specifically:

1. <u>Pile-up of Alleles</u>: Using *cellSNP*^*83*^, alleles were aggregated from polymorphic sites overlapping snRNA-seq reads within expressed genes (genes expressed by at least 10 cells were included). Polymorphic sites required a minimum minor allele frequency of 0.1 and a minimum aggregated UMI count of 20.
2. <u>Donor Assignment:</u> *Vireo*^*84*^ was used to cluster cells into six distinct donor groups per pool based on allele pile-ups. Each cluster’s identity was assigned through genotype concordance analysis, comparing cell clusters to reference genotyping data using QTLtools-mbv^85^.
3. <u>Baseline QC for Genotyping:</u> Only cells meeting baseline QC thresholds (expressing at least 1,000 genes and with a mitochondrial read fraction below 5%) were included in this analysis.

This pipeline detected and corrected occasional sample swaps and mislabeling, ensuring accurate donor assignment for the majority of pools.

##### Rigorous QC Pipeline

After genome alignment and demultiplexing, downstream QC and processing were conducted using *Pegasus* (v1.7.0)^86^ and *Scanpy* (v1.9.1)^87^, with additional steps to ensure robust data integrity:

1. <u>Individual Cell QC</u>: Cells were excluded if their UMI counts were outside the range of 1,500 ≤ UMI ≤ 110,000, gene counts were outside 1,100 ≤ genes ≤ 12,500, or if their mitochondrial content exceeded 5% (mitochondrial fraction > 0.05). Ambient RNA contamination was mitigated using CellBender^88^, as well as the proportion of reads mapped to non-mRNA categories like rRNA, sRNA, and pseudogenes, in addition to examining confounding factors such as the lncRNA MALAT1. Doublets were identified and removed using Scrublet^89^.
2. <u>Feature-Level QC</u>: Genes not expressed in at least 0.05% of nuclei were removed.
3. <u>Donor-Level QC</u>: Donors with fewer than 50 nuclei were excluded to avoid introducing noise into downstream analyses.

##### Data Integration

To correct for (non-biological) variance, including tissue dissection biases or brain bank resources, we employed Canonical Correlation Analysis using the *Harmony* (v0.1)^90^. After identifying highly variable features based on mean-variance constructed a k-nearest-neighbor (kNN) graph using Harmony-corrected PCA embeddings. The Leiden algorithm was applied to cluster cells by type.

Basic QC of the DLPFC lifespan dataset is presented in **Supplementary Fig. 2a**. These include the distribution of nuclei across donors and a histogram of nuclei per sample, as well as distribution plots for mitotic ratio, ribosomal gene content, total counts (n_counts), and the number of genes (n_genes), all split by brain bank.

##### Joint cellular taxonomy

The DLPFC lifespan cohort was prepared as a subset from the broader “PsychAD dataset,”^12,13^ which we defined in a separate manuscript (Lee et al., 2024 and Fullard et al., 2024). The “PsychAD dataset” includes a comprehensive cellular taxonomy with all levels of classification-class, subclass, and subtype.

The taxonomy for the “PsychAD dataset” was established using a modular and iterative clustering approach, as described in Lee et al., 2024 and Fullard et al. 2024. This process involved marker validation, label transfer using scANVI^91^ based on the reference dataset from Mathys et al., 2023^14^, and subclass-level validation through Xenium spatial experiments. The hierarchy of taxonomy labels - class, subclass, and subtype - was derived from classifications outlined in Mathys et al., 2023^14^. For the current study, the DLPFC lifespan cohort was isolated as a subset of this annotated dataset for further analysis. The subclass level correlation of taxonomy with Mathys et al., 2023^14^ as well as visualization of key gene markers is displayed in **Supplementary Fig. 3**, respectively.

##### UMAP

Initially, 6,000 highly variable genes (HVGs) were selected based on mean and dispersion trends, using default parameters (min_mean=0.0125, max_mean=3, min_disp=0.5) and excluding sex chromosomes, mitochondrial genes, and using “MT” as batch variables. A k-nearest-neighbor (kNN) graph was generated using the harmony-corrected PCA embedding space, which facilitated clustering of nuclei by cell type using the Leiden clustering algorithm. UMAP projection was then utilized to visualize these clusters. For each class-level cluster, data were subsetted, and HVGs were recalculated within each class to fine-tune the feature space relevant to that class. A new kNN graph was generated based on the harmony-corrected PCA of these HVGs, followed by Leiden clustering to annotate subclass-level identities. This iterative process resulted in the identification of 65 subtypes of DLPFC cells. Canonical markers of major cell types, reference datasets^14,92^ as well as spatial validation were utilised for class/subclass/subtype-level labels of the joint “PsychAD dataset”^13^. The DLPFC lifespan cohort was then extracted and the UMAP was re-calculated using the same approach; for the full dataset in current study, 6,000 HVGs were identified using the `hvg` function in *Pegasus* (v1.7.0)^86^, while excluding mitochondria and sex-specific genes. Principal component analysis (PCA) was performed on the gene expression data, followed by batch correction using *Harmony* (v0.1)^90^. Subsequently, UMAP dimensionality reduction was generated based on the first 30 harmony-adjusted principal components^93^ (**Fig. 1a**). Finally, a nearest-neighbour graph using the first 30 harmony-adjusted principal components was calculated using the `sc.pp.leiden` function. UMAT calculation (**Supplementary Fig. 1b**) was run based on the approach introduced in^11^.

#### Single-cell polygenic disease risk score

We utilized *scDRS* package (v1.0.1)^87^ to evaluate the combined expression of potential disease-associated genes obtained from GWAS summary statistics through MAGMA analysis^94^ **(Supplementary Fig. 4)**. Each gene’s contribution was weighted by its MAGMA z-score from GWAS and inversely weighted by its gene-specific technical noise level in the single-cell data. This process was performed across each cell of the Aging single nuclei dataset to generate raw disease scores specific to each. Additionally, we generated 200 sets of raw control scores, matched in gene set size, mean expression, and expression variance to the disease-associated genes. Subsequently, we normalized both the raw disease scores and raw control scores for each cell, resulting in normalized scores. These calculations were executed using the `scdrs.score_cell` function, with the following parameters:

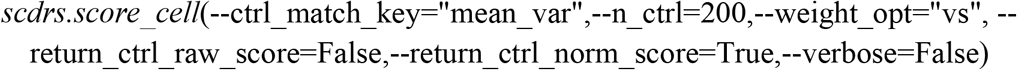

For further analysis, subclass-level examinations were conducted to link predefined subclasses to disease and evaluate the heterogeneity in disease association across cells within each predefined subclass level of taxonomy. This was achieved using the `scdrs.method.downstream_group_analysis` function with default settings. The output from this step is provided in **Supplemental Data 2**. All 24 GWAS traits utilized in the study are listed in **Supplemental Table S2**.

scDRS analysis for the pseudotime section was performed consistently across each cell of the integrated single nuclei dataset. Single-cell disease scores along each trajectory were compressed into 500 meta-cells per trajectory.

Those scores were then modelled using a generalized linear model *disease score* ∼ *splines*: : *ns*(*pseudotime, df* = 3). Unlike MAGMA, which assesses gene-level GWAS enrichment across predefined cell types, scDRS provides per-nucleus scores by weighting gene expression with GWAS-derived effect sizes. This allowed us to pinpoint disease-relevant subpopulations that may not align with standard cell-type annotations, offering finer resolution into how aging intersects with genetic risk.

#### Pseudobulk data aggregation and covariate selection

To quantify the variance explained by age, age-associated changes in the transcriptome and lifespan trends, we pseudobulked gene expression data by aggregating the counts for each donor from 1,307,674 nuclei, stratified by subclass and four age groups (developmental, young, middle and late adulthood) using the `aggregateToPseudoBulk` function from *dreamlet* (v1.1.17) R package. Note, all steps in this section are done using functions from the *dreamlet* (v1.1.17) R package. Subsequently, we applied voom normalization to the age group-subclass-specific gene-by-donor matrix using the `processAssays` function, which filters for samples with at least 5 nuclei per subclass and genes with a minimum of 5 reads per donor. This process generated four lists containing genes x donor matrices for each subclass corresponding to each age group.

With *Source, Sex*, and *PMI* as the base model, technical and biological covariates were identified using a forward stepwise regression approach on subclass-specific pseudobulk expression data on the entire “PsychAD dataset”. Since the lifespan nuclei in this study are a subset of the PsychAD dataset, we implemented the same model for *i* subclass and the *j* gene in our analysis as described in equation (eq) (1). From hereon we use these variables as variable in other models in succeeding sections.

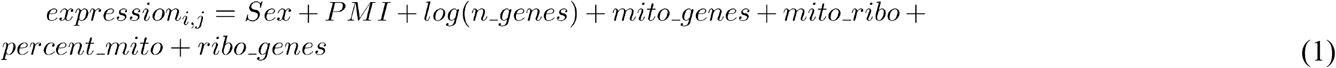

The details of these covariates are as follows: *PMI* (post-mortem interval in hours) is coded as a scaled numerical variable, while *Source* and *Sex* are categorical variables. Source is excluded from the equation for any downstream analyses for the developmental group, since all samples are from a single brain bank (HBCC) (**Supplemental Data1**). The number of genes from each nuclei *n_genes*, and the proportion of mitochondrial genes per donor *percent_mito* are summarized for each donor and were obtained using the `qc_metrics` function from the *pegasus* (v1.8.1)^86^ package. Scores for reference mitochondrial genes from chrM *mito_genes* and mitochondrial ribosomal genes not from chrM *mito*, as well as ribosomal genes from both large and small units *ribo_genes*, are estimated for each nucleus and summarized for each donor using the `calc_signature_score` function from *pegasus* (v1.8.1)^86^. **Supplementary Fig. 2c** shows the pairwise correlation of these covariates with age.

#### Quantification of variance in age

We quantified the variance explained by age for age group-subclass-specific pseudobulk expression data for *i* subclass and the *j* gene as *expression*_*i,j*_ = *Age* + *covariates* in the `fitVarPart` function from *dreamlet* (v1.1.17) R package. VariancePartition^15^ is a statistical framework designed to prioritize drivers of variation in gene expression, utilizing a linear mixed model to quantify contributions from factors such as disease status, sex, cell type, and technical variables. The covariates were obtained from eq(1). **Fig. 1c** highlights the mean contribution of numerical variable age in pseudobulk expression for each subclass across four age groups. **Supplementary Fig. 2d** shows the combined contribution of covariates and age from four groups. The output from this section is provided in **Supplemental Data 3**.

#### Lifespan dynamics of nuclei counts

To quantify changes in nuclei counts as a function of age we utilized `crumblr` function from *crumblr*^*27*^ (Count Ratio Uncertainty Modeling Based Linear Regression) (v0.99.6) R package. Traditional methods (e.g., per-cell-type proportion tests) may miss subtle, coordinated shifts across related subtypes and often overlook covariate effects. Crumblr addresses this by applying precision-weighted linear mixed models within a multivariate framework, improving sensitivity to aging-related cell type composition changes across the cell lineage hierarchy. The package employs a three-step process to quantify the association of changes in nuclei counts with user-defined independent variables. 1) Normalization: Nuclei counts are normalized using a Dirichlet multinomial distribution. 2) Modeling: A standard dream precision-weighted linear mixed model with empirical Bayes estimation is implemented to obtain association statistics for each measurement. 3) Multivariate hypothesis testing: This step enables the joint analysis of internal nodes in a hierarchical clustering of subclasses, which is crucial for accounting for correlations among related subclasses, such as those among EN subclasses. The sample code of these steps section is provided in the repository (**code availability section**).

In this section, we performed two analyses to quantify changes in nuclei counts: (a) as a function of age across the entire lifespan, and (b) within specific age groups. These analyses were limited to subclasses with at least 500 nuclei counts resulting in 26 out of 27 subclasses. For both analyses, we explored the optimal model to represent nuclei counts as a function of age, aiming to determine whether a linear or logarithmic relationship would be optimal. To test these models, we first regressed out the subset of covariates: sex, PMI, and source from nuclei counts for subclass across 284 donors using the equations (2) and (3). The other covariates are more relevant to gene expression analysis so we kept them out for nuclei composition analysis.

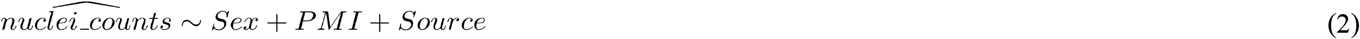

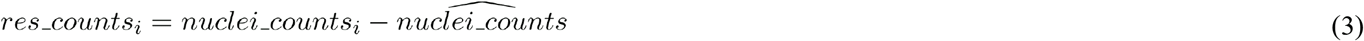

After this, we tested two models *res_counts*_*i*_∼*Age, res_counts*_*i*_ *= log*_2_(*Age*+1) and obtained the difference of Bayesian Information Criterion (BIC) Δ*BIC*_*i*_ for *i* subclass. Using Δ*BIC*_*i*_ metric, we found *log*_2_(*Age*+1) showed optimal relationship between age and changes in nuclei counts for most subclasses as shown in **Supplementary Fig. 6a**. Applying k-means clustering to subclass specific coefficients from the *log*_2_(*Age*+1) model (**Supplementary Fig. 6b)**, we identified two distinct clusters: one with positive coefficients indicating a log-increasing trend, and another with negative coefficients indicating a log-decreasing trend, as illustrated in **Extended Data Fig. 1a**. To further quantify these lifespan associations, we performed *crumblr* enabled modeling and multivariate test analysis using the eq(4). The coefficient of *log*_2_(*Age*+1) is shown in **Extended Data Fig. 1c** and output is provided in **Supplemental Table 3**.

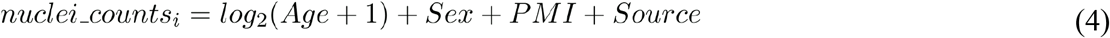

Next, we performed age-group specific analysis by first quantifying the variance explained by age in normalized nuclei counts for each age group using `fitExtractVarPartModel` from *variancePartition*^*15*^ (v1.33.11) R package including eq(4) as a model. **Fig. 1e** shows the distribution of variance explained by *log*_2_(*Age*+1) in 26 subclasses from four age groups. Subsequently, we quantified the coefficients of *log*_2_(*Age*+1) and performed *crumblr* enabled modeling and multivariate hypothesis test using the eq(4) for each age group (“source” covariate was removed from eq(4) for developmental group). **Fig. 1f** shows the heatmap of coefficient of *log*_2_(*Age*+1) for each subclass and age group. The results of this analysis are provided in **Supplemental Table 3**.

To test whether early developmental changes (0–20 years) may dominate the modeling, we performed a breakpoint analysis (**Supplementary Fig. 5c-d**). We iteratively split the dataset at age thresholds ranging from 20 to 30 years and modeled changes in nuclei counts separately for the <cutoff and ≥cutoff groups. This revealed a plateau in significant age-related changes after age 23, suggesting stabilization in adulthood. We then selected age 24 as an optimal breakpoint and visualized subclass-specific trends in development vs adulthood.

Finally, to investigate potential biological mechanisms underlying the most vulnerable age group to compositional changes in nuclei, we performed a pathway activity analysis stratified by subclasses showing log-increasing or log-decreasing age trends. We first curated a set of 374 brain-relevant Gene Ontology Biological Process (GOBP) pathways from MSigDB using keyword-based filtering with terms such as “neuron,” “glia,” “synapse,” “axon,” “oligodendrocyte,” “astrocyte,” “microglia,” and “cortex”. For each nucleus, pathway activity scores were calculated using the over representation analysis (ORA) method implemented in the decoupler python package. This method performs over-representation analysis using a Fisher’s exact test applied to the top 5% most highly expressed genes in each nucleus, relative to the genes annotated to each pathway. The resulting nucleus-by-pathway activity score matrix was aggregated at the subclass level by averaging across all nuclei within each subclass. Subclasses were then annotated with their lifespan trajectory group (log-increasing or log-decreasing), as defined by the compositional modeling described above. To identify biological pathways associated with these aging trends, we performed Welch’s t-tests comparing subclass-level activity scores between the two trajectory groups. Pathways with FDR-adjusted p-values < 0.05 were retained and ranked by the absolute difference in mean activity scores. We selected the top 15 pathways with higher activity in log-increasing subclasses and the top 15 with higher activity in log-decreasing subclasses for visualization. Pathway scores were min-max normalized across subclasses, and subclass annotations were used to group columns by major cell class as shown in the heatmap (**Extended Data Fig. 1f**)

### Validation of nuclei composition changes with age using RNAscope

To validate age-associated changes in nuclei composition, we collected formalin-fixed paraffin-embedded (FFPE) tissue blocks from the DLPFC of eight donors across the lifespan. Samples were obtained from the Neuropathology Brain Bank and Research CoRE at the Icahn School of Medicine at Mount Sinai. Donors were stratified into four life stages: Developmental (n = 3): 0.33, 0.75, and 9 years, young adulthood (n = 1): 38 years, middle adulthood (n = 2): 51 and 53 years and late adulthood (n = 2): 65 and 86 years. To assess subclass-specific changes in nuclei composition across the lifespan, we selected marker genes for IN_SST and Oligo subclasses, which exhibited opposing age-related trends: log-linear decreases for IN_SST and increases for Oligo (**Extended Data Fig. 1e**). IN_SST cells were labeled using the *SST* gene, detected with the RNAscope probe Hs-SST-C2 in channel C2 (Cat. No. 310591-C2), while oligodendrocytes were labeled using the *GPR37* gene, detected with the RNAscope probe Hs-GPR37-C3 in channel C3 (Cat. No. 513631-C3).

#### RNAscope assay and true Black treatment pipeline

RNAscope Multiplex Fluorescent v2 assay (ACD, UM 323100/Rev B) was performed on FFPE DLPFC sections (5 µm). Slides were deparaffinized, rehydrated, and underwent target retrieval using preheated 1X Target Retrieval Reagent (≥99°C), followed by ethanol dehydration and drying. A hydrophobic barrier was drawn around tissue sections, which were then treated with hydrogen peroxide (10 min) and Protease IV (30 min) at room temperature. Gene-specific probes were hybridized at 40°C for 2 h, followed by AMP1–3 incubations (30 min each) with washes in between. Fluorescent signal amplification was performed using sequential HRP-C1, C2, and C3 detection, each with distinct fluorophores. Nuclei were counterstained with DAPI. To reduce lipofuscin autofluorescence, slides were treated with 1X TrueBlack (23007, Biotium) in 70% ethanol for 30 s, then washed with PBS and mounted using ProLong Gold Antifade Mountant (P36930, Thermo Fisher).

#### Imaging

RNAscope images were acquired using the EVOS M7000 microscope (Thermo Fisher) with a 40X objective (OLY XAPO 40X, NA 0.95). For each tissue section, 3–4 high-quality regions of interest (ROIs) were selected to capture representative areas spanning the full grey matter and half of the white matter. Imaging was performed using DAPI, RFP, and Cy5 filter cubes to detect nuclei and RNAscope signals (C2 and C3 channels). ROIs were scanned using the serpentine horizontal pattern in quick scan mode with autofocus on every DAPI field. Each ROI consisted of 100–200 fields of view (FOVs), saved as 16-bit TIFF files.

#### Image analysis

FOVs were stitched into full ROI images using a custom Fiji (ImageJ) macro based on the Grid/Collection Stitching Plugin. DAPI channels were stitched first (snake-by-rows layout with subpixel accuracy), generating a reference TileConfiguration.registered.txt used to stitch the corresponding C2 and C3 channels for alignment. Stitched channel images were combined into hyperstacks using Fiji’s “Images to Stack” and “Stack to Hyperstack” functions. For quantification, stitched ROIs were analyzed in QuPath. Cells were segmented using the Cell Detection tool, and RNAscope puncta were detected with the Subcellular Detection function. Single-cell expression and composition data were exported using saveDetectionMeasurements.

#### Statistical analysis

To quantify subclass-specific expression, we analyzed QuPath output tables from stitched RNAscope images across eight donors. For each region of interest (ROI), we calculated the fraction of positive cells by dividing the number of cells with ≥2 RNA molecules for the target marker (e.g., SST) by the total number of detected nuclei in that ROI. This metric quantifies the relative abundance of marker-positive cells and serves as a proxy for cell composition across age groups. Similar to the lifespan model as described in (**Extended Data Fig. 1a**), age of eight donors was modelled as log_2_(Age + 1). Statistical analysis and visualizations were performed using dplyr, ggplot2 packages.

##### Age-associated transcriptomic changes

To assess the age-related transcriptomic changes for each subclass and age group, we performed *dreamlet* enabled limma modeling on age group-subclass-specific pseudobulk expression data as described in the “Psuedobulk data aggregation and covariates selection”. The model implemented for *i* subclass and the *j* gene is *expression*_*i,j*_ = *Age* +*covariates*. We conducted final multiple testing correction at the study-wide level (26 subclasses × number of genes per subclass) for each group. The total number of study-wide genes from all subclasses were: development (*n* = 328,991), young adulthood (*n* = 333,471), middle adulthood (*n* = 340,084), and late adulthood (*n* = 292,748). After applying a threshold of FDR < 0.05, we obtained age-associated differential expression genes (aDEGs) per age group, as shown in **Fig.1h** and **Supplementary Fig. 7a-b**. We provided the results of this analysis in **Supplemental Data 4**. For gene set pathway analysis to identify biological processes, we used the `enrichr` function from the *enrichR* (v3.2) R library. The database used was GO_Biological_Process_2021, and the results are shown in **Fig. 1i** and **Supplementary Fig. 7f-g**. The pathway enrichment analysis for all aDEGs per subclass is provided in **Supplemental Data 5**. Dreamlet allowed us to model donor-level variation and batch effects using precision-weighted mixed models, outperforming standard methods. Its efficiency and design-aware approach ensured statistically rigorous identification of differentially expressed genes.

##### Lifespan trends of gene expression

To obtain the lifespan trend of each gene (*n* = 334,689 genes) across 26 subclasses, analysis was performed in three steps. 1) model selection, 2) clustering and 3) prediction of average trajectory as depicted in **Fig. 2a**. The details of these steps are explained below.

###### Model selection

Our objective was to identify a single optimal model that could capture age related non-linear trends in gene expression across all 26 subclasses. We achieved this by fitting 12 different models to expression of *i* subclass and the *j* gene for a total of 334,689 genes using the `dream` function as described in eq(5-8).

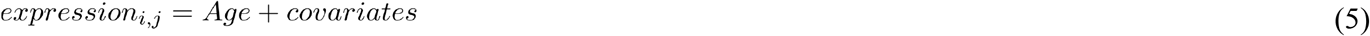

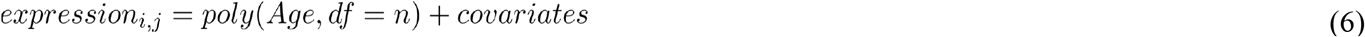

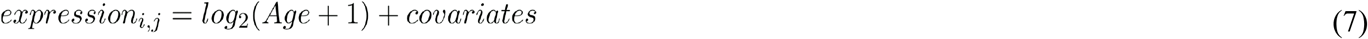

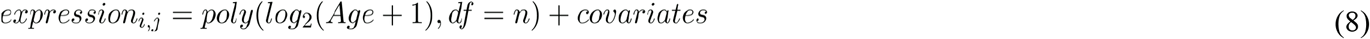

*n* in eq(6,8) represent degrees of freedom which are 2, 3, 4, 5, 6 and `poly` is R function from *stats* base R library, which produces orthogonal polynomials with a given degree of freedom in df argument. To identify the optimal model, we collected the BIC values from each of the 12 models and searched for the minimum values. Although BIC values across all models were not conclusive for glia and other subclasses, a notable decrease in BIC was observed for the *poly* (*log*_2_(*Age*+1),*df* = 2) +*covariates* model specifically in neurons, particularly IN (**Supplementary Fig. 11a**). Since approximately 60% of the subclasses are neuronal, and for simplicity, we adopted the *poly* (*log*_2_(*Age*+1),*df* = 2) +*covariates* model as the optimal lifespan model for all 26 subclasses. All steps in this section were performed using the dream function from *dreamlet* (v1.1.17) R package. The output from optimal model consists of linear 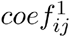 and non-linear coefficient 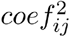 from the model for 334,689 genes.

###### Clustering

Our goal was to identify the optimal number of unique characteristic curves for the lifespan trends of 334,689 genes. To achieve this, we applied k-means clustering to 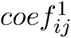 and 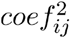 from the dreamlet summary statistics of the optimal lifespan model for 334,689 genes. Clustering all coefficients revealed that k = 10 provided the optimal clusters with non-overlapping lifespan trends (**Supplementary Fig. 11b**). **Supplementary Fig. 11c** shows the stratification of all genes across these 10 clusters. For all downstream analyses, we focused on the 135,120 genes that remained after applying study-wide multiple testing correction (FDR < 0.05) from the optimal model on the 334,689 genes. **Supplementary Fig. 11d** illustrates the stratification of lifespan-aDEGs (135,120 genes) across the 10 trajectories. The output table containing 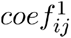 and 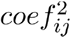, along with other summary statistics from the *dreamlet* tool for each gene and subclass, and trajectory cluster numbers, is provided in **Supplemental Data 7**.

###### Prediction of average lifespan trend

Finally, to visualise an average trajectory per cluster, we calculated the mean of the coefficients for all genes within each cluster, as follows:

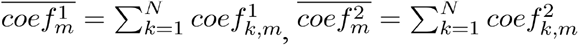, where is total *N* number of genes in *m*^*th*^ cluster and *k* is *j* gene in *j* subclass in *m*^*th*^ cluster. Next we obtained non-linear form of age using 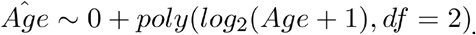 . Using the mean of the coefficients and the polynomial form of age 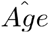, we obtained lifespan trajectory for cluster 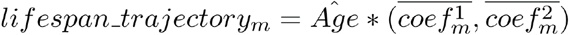 as shown in **Fig. 2b. Supplementary Fig. 12b** shows predicted average trajectory for each subclass in a trajectory cluster using the mean of subclass coefficients 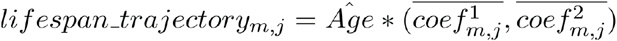. To further obtain biological insight on genes within trajectory clusters, we performed gene set pathway analysis using the `enrichr` function from *enrichR* (v 3.2) R library. The database used was GO_Biological_Process_2021. **Supplementary Fig. 13a** shows an example of pathway enrichment from trajectory 1 and 10. The full table of pathways is provided in **Supplemental Data 8**.

### Integrated analysis of lifespan replication dataset

To validate the age associated transcriptomic changes identified in **Fig.1h**, we integrated human DLPFC snRNA-seq cohort of neurotypical controls from three published datasets^11,29,30^, comprising 306 donors spanning gestational week 22 to 101 years of age.

#### Integration and annotation

Count matrices from multiple snRNA-seq datasets were combined into a single *Scanpy* object (v1.9.3) for joint processing and filtering. Genes expressed in fewer than five nuclei across all batches were excluded. Potential doublets were identified and removed using *Scrublet* (v0.2.3) with 10 principal components per batch. To mitigate sampling bias from variable sequencing depths across studies, we downsampled the integrated data to 1,000 Unique Molecular Identifier counts (UMIs) per nucleus. Downsampling involved random sampling without replacement, and nuclei with less than 1,000 total UMIs were excluded. The count matrix was then scaled to Counts Per Million (CPM) post-downsampling and transformed using natural log plus one. Highly variable genes (n=6,000) were selected from protein-coding genes located in the euchromosome using Scanpy’s `highly_variable_genes` function. Dimensionality reduction was performed via principal component analysis (PCA), retaining components explaining 50% of the variance. A neighborhood graph was constructed using 100 neighbors on the reduced components, followed by generation of a two-dimensional UMAP embedding. Cell type annotation was performed at class and subclass levels using scvi-tools (v1.2.0)^95^, transferring labels from our in-house reference cohort.

For replication analysis, we further excluded three fetal donors (gestational week 22-38) and their nuclei to align the age range with that of the discovery dataset (0-97 year). This ensured comparability between datasets while avoiding potential confounding effects from fetal-specific transcriptomic profiles.

#### Replication of aDEGs

To assess the reproducibility of aDEGs identified in the lifespan DLPFC cohort, we applied the dreamlet-enabled limma modeling framework to subclass- and age group–specific pseudobulk expression profiles in the replication dataset. Our covariate model was expression ∼ Age + batch + sex where batch constitutes dataset resource (**Supplementary Fig. 9**). This analysis followed the same modeling strategy as used in the discovery dataset (see “Pseudobulk data aggregation and covariate selection”), allowing consistent estimation of age effects across datasets. We then evaluated replication at three levels (**Supplementary Fig. 10**): a) using all expressed genes to compute the Spearman correlation of age-associated expression changes between the discovery and replication datasets stratified by subclass and age group, b) restricting the analysis to aDEGs (FDR < 0.05) identified in the discovery dataset to assess consistency of direction and magnitude of effects; and c) calculating π1 statistics, which estimate the proportion of discovery aDEGs that show consistent effects in the replication dataset.

#### Replication of lifespan trajectories

To assess the replication of lifespan transcriptomic trajectories, we implemented two complementary strategies. In the first strategy, we evaluated the replicability of gene-level lifespan trajectories by projecting the age-associated gene expression models derived from the discovery dataset onto the replication dataset. Specifically, for each subclass and gene with a significant age effect (FDR < 0.05) in the discovery cohort, we extracted the age spline coefficients (β_1_, β2) from the dreamlet model and applied them to the transformed ages (log2(Age + 1), df = 2) of the replication donors. We then quantified the agreement between predicted and observed expression levels using R^2^ (squared Pearson correlation) as shown in **Supplementary Fig. 15a**. As a second, independent strategy, we applied the full trajectory discovery pipeline as described in the “Lifespan trends of gene expression” section to the replication dataset (**Supplementary Fig. 15b)**. This included re-aggregating pseudobulk expression data, computing nonlinear coefficients using the optimal model, and re-clustering genes into ten trajectories using the same clustering parameters as in the discovery. We then compared the resulting trajectory shapes and gene memberships between the two datasets to evaluate the concordance of age-associated changes as shown in **Supplementary Fig. 15c-d**. We also quantified replication performance across trajectories by plotting the proportion of true positives (π_1_) recovered in the replication dataset for each trajectory as shown in **Supplementary Fig. 15e**.

### Statistical power analysis

To evaluate the sensitivity of our dataset to detect age-associated changes for each age group and across lifespan, we conducted both theoretical and empirical power assessments.

#### Theoretical power estimation

We first estimated statistical power using Cohen’s d, sample size, and a range of moderated R^2^ values (0.5 to 1), which reflect the proportion of gene expression variance explained by age. Power calculations were Bonferroni-corrected for multiple testing, accounting for 26 subclasses and a minimum of ∼11,700 expressed genes per subclass across age groups and whole lifespan. As shown in **Supplementary Fig. 8a**, with sample size ≥ 50, we achieve >80% power to detect effects of d ≥ 1.2 across a range of biologically plausible R^2^ values. Given that all four of our age-defined groups include 50–100 samples: development (n = 53), young adulthood (n = 54), middle adulthood (n = 95), and late adulthood (n = 82), our design is sufficiently powered to support robust group-level comparisons.

#### Empirical power evaluation

To assess empirical variation in power across cell subclasses, we considered both technical and biological factors that influence sensitivity to detect age-associated differential expression. First, we examined the relationship between the number of expressed genes and the number of donors per subclass. **Supplementary Fig. 8b, e** shows that subclasses with greater donor coverage tend to exhibit higher numbers of expressed genes, reflecting improved transcriptome detection with increased sample size, particularly in the late adulthood group. In contrast, the development group displays substantial variability across subclasses, especially among excitatory neurons, likely due to underlying biological heterogeneity during early brain development.We further evaluated how these factors impact aDEGs detection sensitivity. As shown in **Supplementary Fig. 8c,f** there is a positive association between the number of aDEGs and the number of expressed genes per subclass. Finally, we assessed the relationship between aDEG count and the magnitude of age-associated effect sizes. **Supplementary Fig. 8d,g** reveals an inverse relationship: low-powered subclasses (e.g., SMC, PC, VLMC) with fewer expressed genes and low aDEG counts show inflated effect sizes, consistent with the winner’s curse, while well-powered subclasses such as neurons and glia exhibit smaller, more stable effect estimates.

Finally, we evaluated power using lifespan aDEGs (defined by F-statistics across both linear and non-linear age terms) across subclasses. **Supplementary Fig. 8h** displays the overall distribution of donor coverage and gene expression for each subclass using this lifespan non-linear optimal model. **Supplementary Fig. 8i–j** extend our analysis by correlating lifespan aDEG counts with (i) the number of expressed genes and (j) the mean absolute age effect size.

### Degree of sharing across EN, IN and Glial subclasses

To identify genes with similarity in effect sizes across subclasses, the association statistics from *dreamlet* was not sufficient. For example, if the age-associated effect size for a gene was significant in one subclass but not in another, one might have concluded that the age effect was subclass-specific. However, the absence of a significant effect in another subclass did not necessarily mean that the effect size was zero. This scenario often occurs when statistical power is limited, or varies significantly between subclasses. To overcome this limitation, we utilized *mashr*^*38*^ (v 0.2.79) R library, which used an empirical Bayes approach to learn patterns of similarity in effect sizes across subclasses and then leveraged these prior patterns to improve the accuracy of effect size estimates.

We assess gene sharing across 9 EN, 7 IN and 4 glial subclasses. The EN group included 9 subclasses (EN_L6_CT, EN_L5_6_NP, EN_L6B, EN_L3_5_IT_1, EN_L3_5_IT_2, EN_L3_5_IT_3, EN_L2_3_IT, EN_L6_IT_1, EN_L6_IT_2), the IN group included 7 subclasses (IN_LAMP5_RELN, IN_LAMP5_LHX6, IN_ADARB2, IN_VIP, IN_PVALB_CHC, IN_PVALB, IN_SST), and the glial group included 4 subclasses (Astro, Oligo, OPC, Micro). From hereafter, we refer to EN, IN and Glial subclasses sets as cE,cI and cG respectively.

In this section, we conducted two analyses: 1) we quantified the degree of sharing for each gene based on the similarity of age-associated effect size patterns across EN, IN, and glial subclasses using `run_mash` function built in *dreamlet* adapted from *mashr* (v0.2.79)^38^ R library, and 2) we calculated the tau score to quantify the cell specificity of each gene, allowing us to stratify shared and non-shared genes.

#### Mashr analysis

Using the age-associated summary statistics per age group from **Supplemental Data 4**, we built two matrices with genes as rows and subclasses as columns, containing log_2_FC (*β*_*i,j*_) and standard errors (*SE*_*j,i*_). The number of genes included are: development (*n* = 23,914 genes), young adulthood (*n* = 26,107 genes), middle adulthood (*n* = 26,809 genes), and late adulthood (*n* = 24,795 genes) time points. Any gene without summary statistics for a subclass was replaced with 0. Using the empirical bayesian framework, *mashr* computed posterior mean and variance by combining covariance matrix derived from prior distribution of effect sizes and matrix of observed variance to assess sharing of *j* gene across subclasses sets *c*_*E*_,*c*_*I*_ *and c*_*G*_ . The posterior mean 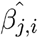 provides a more accurate estimate of the true effect size *β*_*i,j*_, while the posterior variance quantifies the uncertainty in 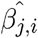. After this step, *mashr* reported the local false positive rate, which is defined as the probability that the true effect size *β*_*j,i*_ had the opposite sign of the estimated effect size 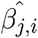 . For example, for *j* gene within *c*_*E*_ set, 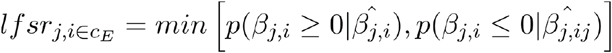 where *β*_*i,j*_ is true age effect size and 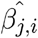 is the estimated effect size. Then *p*_*i,j*_ = 1 − lfsr _*j,i*_ is the probability that the true age effect size and estimated effect size are concordant. Thus, for *j* gene, the degree of sharing or composite posterior probability for concordance in effect sizes of EN, IN, and glial classes for each age group can be quantified as the product of probabilities across their respective subclasses, i.e., 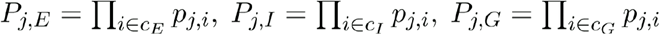. To ensure that each gene has true effect size *β*_*j,i*_ with p-value < 0.05 from *dreamlet* regression as well, we applied an additional filter: a dreamlet p-value of the gene < 0.05 in at least two subclasses in a grouped class. **Supplementary Fig. 16a** shows the number of genes obtained after this step. Composite posterior probability *P*_*E*_, *P*_*I*_, *P*_*G*_ were stratified into ten equally sized bins to display the number of genes in each bin. Bin 1 represented genes with a composite prob. < 0.1, indicating less sharing, while bin 10 included genes with a composite prob. > 0.9, indicating high sharing (**Supplementary Fig. 17a** and **Extended Data Fig. 2b). Supplementary Fig. 17b** demonstrates an example of the distribution of composite posterior probability from EN *P*_*E*_ from 24,795 genes during late adulthood. The heatmap of true age effect sizes *β*_*j,i*_ shows a highly concordant pattern across 9 subclasses of ENs with a composite probability greater than 0.9. In contrast, the effect sizes are heterogeneous across the nine EN subclasses for genes with a probability less than 0.01 (**Supplementary Fig. 17b)**. Using the threshold *P*_*j,E*_ *P*_*j,I*_, *P*_*j,G*_ > 0.9, we obtained the shared genes for each grouped class across four age groups (**Supplementary Fig. 16a)**. Interestingly, no significant overlap was observed in bin 10 across the age groups for each subclass (**Supplementary Fig. 16b-d)**. The output from this step is provided as *P*_*E*_, *P*_*I*_, *P*_*G*_ for each gene per age group in **Supplemental Data 11**.

Using these shared genes per age group per grouped class we identified a network of genes which encode significant protein-protein interactions (PPI) using *STRING-db* ^*40*^ (v 12.0). **Supplementary Fig. 20a-c** show the significant PPI with highest confidence interaction score > 0.9 and genes in networks of at least 5 genes. The clusters of each PPI per grouped class per age group are given in **Supplemental Data 13**.

#### Cells specificity score

Next we reasoned that shared genes with *P*_*j,E*_ *P*_*j,I*_, *P*_*j,G*_ > 0.9 have significantly lower cell specificity compared to genes < 0.9. To validate this we estimated tau score adapted from GTEX ^39^ studies of each gene for each grouped class per age group. Tau score indicates how specifically a gene is expressed across various subclasses. In other words, genes with a tau score close to 1 are more specifically expressed in one subclass, while those with a tau score closer to 0 are equally expressed across all subclasses within a grouped class. To do this, we integrated pseudobulk gene expression data from a grouped class per age group using the `stackedAssays` function, resulting in expression of genes and subclass * donors specifically 17,205 X 2,393; 17,205 X 1,945 and 17,205 X 1,129 for EN; IN and glia respectively. The analysis was limited to protein coding genes. Next, we estimated the median expression of donors across each subclass using the equations below:

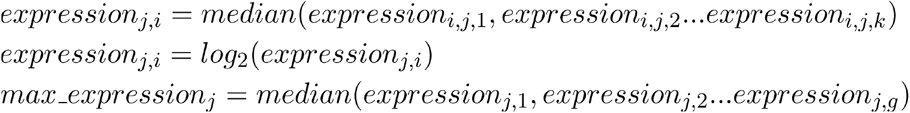

Finally tau score was estimated as 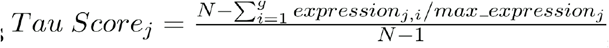, where N is the total number of subclasses. Tau score was kept for cell specificity measurement and was retained for only those genes with sum of median expression value across all subclasses > 10 cpm. **Supplementary Fig. 18c** and **Supplementary Fig. 19** shows the distribution of tau scores stratified by shared and non-shared genes for each grouped across and each age group for a list of genes from *mashr* analysis and aDEGs. The tau score of each age group across 3 grouped classes are provided in **Supplemental Table 4**. The gene set pathways of shared genes, shown in **Extended Data Fig. 2e**, were obtained using the *enrichr* function from the *enrichR* (v 3.2) R library and the full table is provided in **Supplemental Data 12**. The analysis utilized the GO_Biological_Process_2021 database and was limited to protein-coding genes.

### Pseudotime analysis and dynamically expressed gene identification and downstream analysis

For pseudotime analysis, we integrated the lifespan DLPFC dataset (current study) with the published snRNA-seq data spanning gestation to adulthood^11^, following the data integration procedure described in the “Lifespan replication dataset integration and validation of trajectories” Methods section. Unlike the replication analysis, annotation transfer was not performed; instead, we assessed dataset similarity post-integration. This resulted in a combined dataset of 311 donors. The UMAT (Umap of MATuration) embedding was constructed as previously described^11^. Trajectory reconstruction and identification of differentially expressed genes along trajectories (traDEGs) were performed as previously described^8^. For each lineage, corresponding cells were selected and genes not observed in ≥ 5 nuclei were excluded. Six thousand HVGs were selected, data dimensions were reduced via PCA to components explaining 50% of the variance, and the UMAT embedding was recalculated. Pseudotime trajectory analysis was then conducted using *Monocle3* (v1.0.0) based on the UMAT embedding. The shortest path between the developmental node and the node in the mature subclass/subtype clusters was isolated as the corresponding trajectory graph. Cells along the trajectory were selected, and traDEGs were identified using *Monocle3*’s modified `graph_test` function with Moran’s *I* test, including covariates (Sex, Batch, PMI, and log_n_genes) to ensure results were not affected by uneven contributions from different subjects and conditions. Genes with an adjusted p-value < 0.05 and Moran’s *I* ≥ 0.05 were considered statistically significant DEGs. To cluster the DEGs in each lineage, single-cell expression data along each trajectory were compressed using a sliding window along pseudotime, averaging the expression of neighboring cells to generate 500 meta-cells per trajectory. Each gene’s expression was then modeled using a generalized linear model (*expression* ∼ *splines*: : *ns*(*pseudotime, df* = 3)), and k-means clustering was performed on the fitted expressions.

GO-term analysis of traDEG clusters was performed using *enrichR* (v3.2) and the GO Biological Process 2023 dataset. To investigate whether the traDEG clusters play a role in neurological, psychiatric, and other traits, we quantified their colocalization with common risk variants from 24 GWAS (**Supplementary Table 2**) using MAGMA analysis.

### Validation of traDEG modules with spatial transcriptomics

To validate spatial expression patterns of traDEG modules, we analyzed Visium spatial transcriptomics data from 10 adult donors (ages 33-62 years; three cortical sections per donor, 30 sections total) from a publicly available dataset^53^. Data was accessed via the *spatialLIBD* R package (v1.19.11)^96^ using `fetch_data(type=“spatialDLPFC_Visium”`).

For each traDEG module, multi-gene expression was summarized per Visium spot using principal component analysis implemented in `vis_gene` with `multi_gene_method = “pca”` . The first principal component (PC1) was used as a composite score representing module expression. Region-specific enrichment was visualized by computing average PC1 scores per BayesSpace-defined spatial cluster (k = 9).

Spatial validation was performed for all glial and neuronal modules. Visualization outputs included heatmaps of average PC1 scores across donors, PC1 score maps for representative sections (Br2743_mid), and violin plots depicting variability across all sections.

### Enrichment of brain and non-brain related risk genes in age and pseudotime associated genes

All analyses to evaluate the enrichment of brain and non-brain related traits were conducted using MAGMA version 1.08b^94^. MAGMA provided robust gene-set enrichment statistics, reinforcing the biological relevance of our findings in the context of aging and disease, it calculates gene-level P-values for each gene and trait by assessing the joint association of all SNPs within the gene region, while accounting for linkage disequilibrium (LD) between SNPs. Gene regions were defined with a window of 35 kb upstream and 10 kb downstream, and LD estimates were derived from the European panel of the 1000 Genomes Project^97^ (phase 3). MAGMA then applies a linear regression framework to determine whether differentially expressed genes show stronger associations with GWAS traits compared to the rest of the genome. Genes overlapping the MHC region (chr6:25-35 MB) were excluded from the analysis. Heatmaps of brain- and non-brain-related traits were generated using the MAGMA pipeline as described. Outputs are provided in the Supplemental Data: Supplemental Data 6 (Fig. 1k and Fig. S8f), Supplemental Data 9 (Fig. 2g-h and Fig. S12c), and Supplemental Data 16 (Fig. 3f, Fig. 4c,g, and Extended Data Fig. 5e,j). For Supplementary Data 16, we extracted significant genes (FDR < 0.05) from MAGMA gene-level outputs for each trait and overlapped them with pseudotime-inferred gene modules to enable detailed gene-level exploration of trait enrichment.

### Inference of transcriptional regulators of pseudotime gene modules

To identify transcription factors (TFs) regulating pseudotime-driven gene modules, we applied the univariate linear model (ULM) approach in decoupleR (v2.9.7)^51^. This method fits a linear model for each TF, using TF-target interaction weights from the regulatory network CollecTRI^98^, and calculates a *t*-statistic as a measure of TF enrichment. TFs with *p* < 0.05 were considered significant, and inference was performed on meta-cell expression profiles along lineage-specific pseudotime trajectories.

Protein-protein interaction networks were constructed using STRING-db (as described in the “Degree of sharing across EN, IN and Glial subclasses”) by combining genes from each traDEG module with their corresponding decoupleR-inferred TFs. To prioritize key regulators, module-specific networks were filtered to retain TFs within connected components of size ≥ 5 and node degree ≥ 3. The resulting set of 44 key TFs was further evaluated for biological relevance by comparison to 582 enhancer-driven regulons (eRegulons) identified with SCENIC+ in the developing human neocortex^52^.

### Rhythmicity Analysis

Prior to rhythmicity analysis, time of death (TOD) for each subject was normalized to a Zeitgeber Time (ZT) scale as described by Seney *et al*.^99^. Briefly, the TOD for each subject was collected at local time then converted to coordinated universal time by adjusting for time zone and daylight savings time. Coordinated universal time was further adjusted to account for longitude and latitude of death place. Each subject’s TOD was then set as ZT = *t* hours (h) after previous (if *t* < 18) or before next (if *t* ≥ -6) sunrise. The distribution of subject TODs across age groups is shown in (**Extended Data Fig. 6a**). The infancy, childhood and adolescent groups had too few subjects with known TODs to run further rhythmicity analyses, and the young adulthood group had large gaps in their distribution. As such, we excluded the infancy, childhood, and adolescent groups and combined the young and middle adulthood groups (YA+MA; *n* = 119), which was then compared to the late adulthood group (LA; *n* = 77) (**Fig. 6a**).

To measure the rhythmicity in subclass and genes and for other downstream analyses in this section, we first regressed out the effects of identified technical and biological covariates in the previous section along with age using the eq(9) described below.

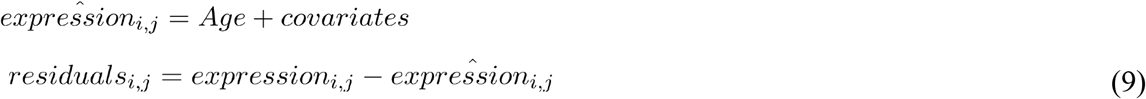

Then, using the DiffCircaPipeline workflow^100^, 24 h rhythms in gene expression were detected and compared between YA+MA and LA groups. First, samples were ordered by TOD and residualized expression for each transcript was fit to a cosinor model separately in each group. The most robust rhythmic genes were identified as those that survived multiple comparison testing (adj.p-value ≤ 0.05). For quality control, we also performed rhythmicity analyses on 100 permutations of our data with scrambled TOD values within both the YA+MA and LA groups (**Supplementary Data 18)**. We then used these permutations to calculate empirical p-values: [(Number of permutation R2 values ≥ Observed R2 value) + 1)]/[(Number of permutations)+1)].

For a deeper analysis, we used a p < 0.01 threshold within the DiffCircaPipeline to categorize the type of rhythmicity (rhythmic in YA+MA, rhythmic in LA, both, or arrhythmic) of each gene (**Supplementary Data 17**). These categories were then used to determine which genes to perform differential goodness of fit and parameter tests on to reduce the likelihood of Type I error^100^. Specifically, differences in goodness of fit (ΔR^2^) between YA+MA and LA were determined through a permutation test (1000 permutations) in transcripts identified as rhythmic in YA+MA, LA, or Both (**Supplementary Data 20**). Additionally, a global differential parameter test was performed in transcripts identified as rhythmic in Both groups, followed by *post hoc* analyses to determine whether differences were due to the rhythmic paramters amplitude, MESOR, or phase (**Supplementary Data 19**). enrichR was then used to identify pathways enriched in rhythmic genes (**Supplementary Data 21**). This was done for all rhythmic genes within a subclass in YA+MA and LA, and then separately for rhythmic genes that peaked between 0-8 h ZT and 20-24 h ZT (the “Sunrise” group) and 8 - 20 h ZT (the “Sunset” group).

## Data Availability

All data can be accessed via Synapse, as part of the PsychAD Study.The data are available under controlled use conditions set by human privacy regulations. To access the data, a data use agreement is needed.

## Data availability

The DLPFC lifespan snRNA-seq profiling data can be accessed via Synapse under accession code syn52396927, as part of the PsychAD Study detailed at syn60084804. The dataset, analysis outputs are available via the AD Knowledge Portal (https://adknowledgeportal.org). The AD Knowledge Portal is a platform for accessing data, analyses, and tools generated by the Accelerating Medicines Partnership (AMP-AD) Target Discovery Program and other National Institute on Aging (NIA)-supported programs to enable open-science practices and accelerate translational learning. The data, analyses and tools are shared early in the research cycle without a publication embargo on secondary use. Data is available for general research use according to the following requirements for data access and data attribution (https://adknowledgeportal.synapse.org/Data%20Access). For access to content described in this manuscript see: https://doi.org/10.7303/syn52396927. The data are available under controlled use conditions set by human privacy regulations. To access the data, a data use agreement is needed. The registration is in place solely to ensure the anonymity of the study participants. In addition, we have a data descriptor manuscript detailing the data processing and data collection. Interactive visualizations of the single-cell data can be explored through CELLxGENE: https://cellxgene.cziscience.com/e/4442d412-91cb-4261-acca-8adf5fa04c11.cxg/.

## Code availability

All the source codes utilized in this study are available via GitHub: https://github.com/gk1610/DLPFC_Lifespan_Atlas

Additional code deposit at Zenodo is available for crumblr analysis: https://doi.org/10.5281/zenodo.12752107

## Acknowledgments

We extend our deep gratitude to the patients and their families for their generous donation of invaluable biological material, which was essential for the success of this study. Their unwavering participation and dedication to advancing scientific knowledge and enhancing human health are deeply appreciated. We also acknowledge the generous support of the National Institute on Aging, who provided funding for this research through the following NIH grants: R01AG067025, R01AG082185, and R01AG065582. Human tissues were obtained from the NIH NeuroBioBank at the Mount Sinai Brain Bank (MSSM; supported by NIMH-75N95019C00049), and NIMH-IRP Human Brain Collection Core (HBCC, project # ZIC MH002903). This research was supported in part by the Intramural Research Program of the National Institutes of Health (NIH). The contributions of the NIH author(s) were made as part of their official duties as NIH federal employees, are in compliance with agency policy requirements, and are considered Works of the United States Government. However, the findings and conclusions presented in this paper are those of the author(s) and do not necessarily reflect the views of the NIH or the U.S. Department of Health and Human Services. The results published here are in whole or in part based on data obtained from the AD Knowledge Portal.

## Author contributions

Conceptualization: PR, KG

Methodology & Software: GEH, KG, HY, MRS

Validation: HY, KG, XW

Formal analysis: KG, HY, MRS, TC

Investigation: JFF, XW, MP, SV, AH, CC, SRM, ZS, MA, SA

Resources: VH, PKA, SM, NMT

Data Curation: JB, PNM, SV, MP, DL, HY

Writing: HY, KG, MRS, TC, PR, JFF, XW, DL with support from all co-authors.

Visualization: KG, HY, MRS, TC

Supervision: PR, KG, JFF, CAM, GV, JB, VH, DL, GEH

Project administration: KG, PR

Funding acquisition: PR, VH

All authors read and approved the final draft of the paper.

## Materials & Correspondence

Correspondence to Kiran Girdhar or Panos Roussos.

## Supplementary Information

This supplementary information includes:

- Supplementary Figs. 1-28
- Captions for Supplementary Tables 1-4
- Captions for Supplementary Data 1-21
- Supplementary Notes

**Supplementary Figure 1.**
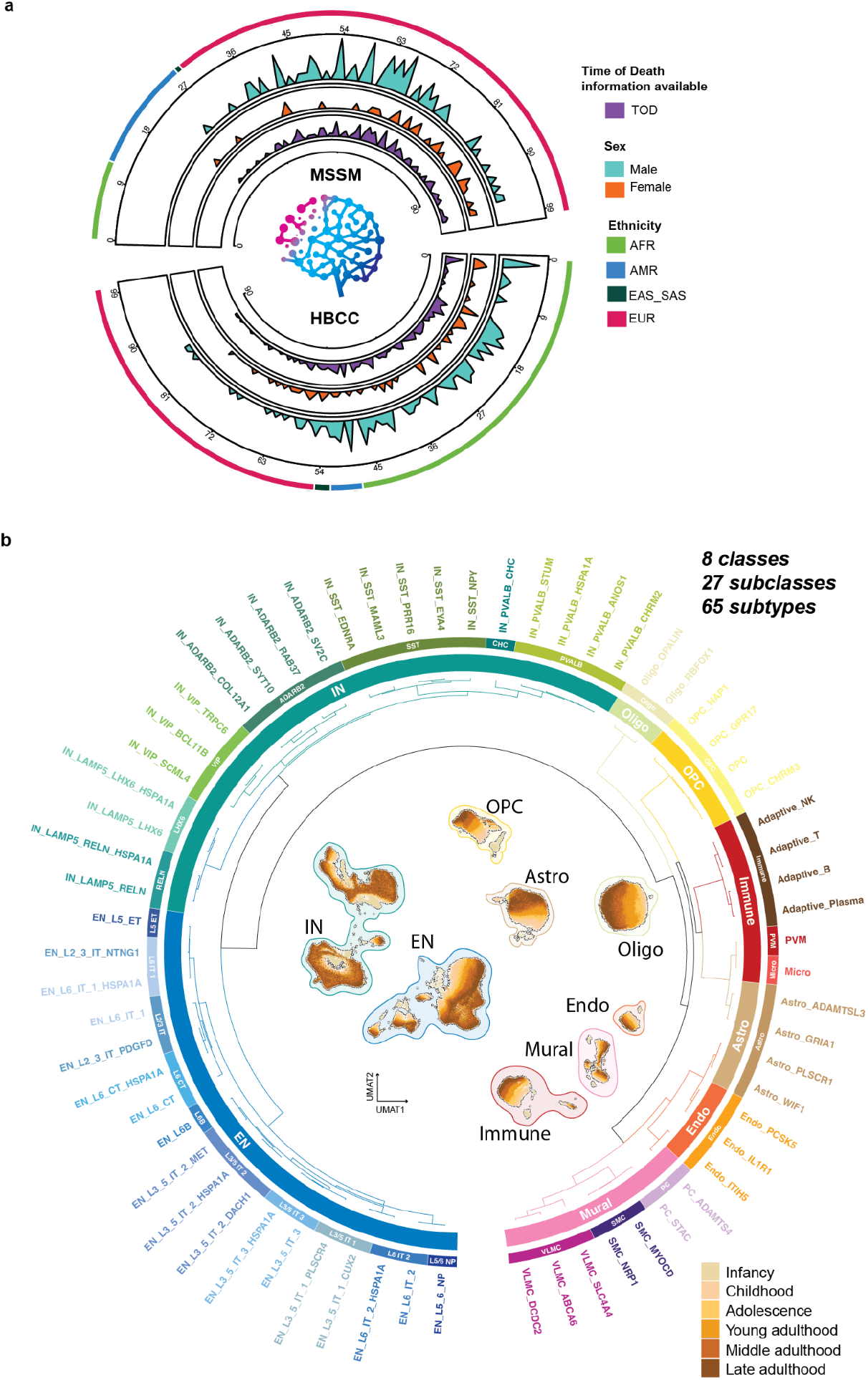
Donor demographics and cell taxonomy in lifespan DLPFC dataset. **a**, Circos plot depicting demographic information of available donors. The plot is split by donors from the MSSM (top) and HBCC (bottom) brain banks, and, from outside-in, indicates: ethnicity (AFR: African, AMR: Ad Mixed American, EAS: East and South Asian, SAS: South Asian, EUR: European), sex (orange Female and cyan Male) and availability of metadata for time of death (purple) are represented as a distribution across age of donors. **b**, Transcriptomic similarities across class, subclass and subtype taxonomic levels. UMAP of maturation (UMAT)^11^ representation of the dataset is positioned within the taxonomy circos plot, colored by age group. The color of the halo encircling each cell class represents class category membership.

**Supplementary Figure 2.**
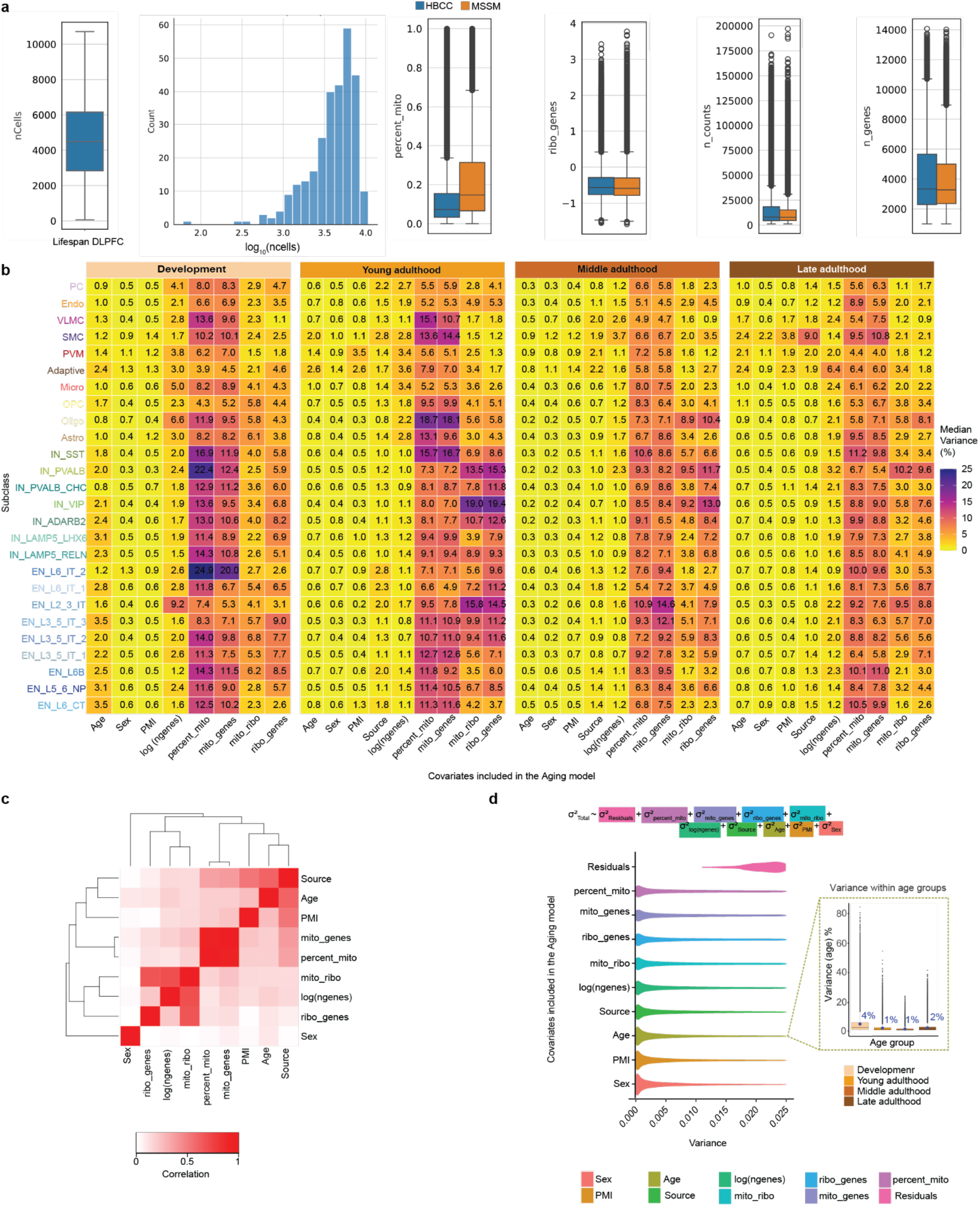
Quality control of snRNA-seq and covariate modeling for pseudobulk gene expression analyses. **a**, Distribution of the number of nuclei per donor (first plot) and log-transformed nuclei per sample (second plot). The third to sixth plots show brain bank-specific distributions for percent mitochondrial reads, ribosomal gene expression, total UMI counts, and total detected genes (HBCC in blue, MSSM in orange). **b**, Heatmap showing the median percent variance explained by each covariate (x-axis) across subclasses (y-axis), stratified by age group. These covariates were included in all age associated and circadian genes analyses done at pseudobulk gene expression level. **c**, Heatmap of pairwise correlation of identified covariates and age. **d**, Combined variance partition^15^ results (from development, young, middle and late adulthood) depicting respective contributions to variance in gene expression explained by listed covariates (Age, source (brain bank), PMI (post mortem interval), mitochondrial and ribosomal properties^87^ (percent_mito, mito_genes, mito_ribo, ribo_genes), log(n_genes) and residuals), ordered from highest to lowest contribution. Highlighted inset boxplot shows distribution of age-contributed variance within the respective age groups. Boxplot in inset shows lower and upper hinges at the 25th and 75th percentiles, with whiskers extending to at most 1.5 times the interquartile range (IQR).

**Supplementary Figure 3.**
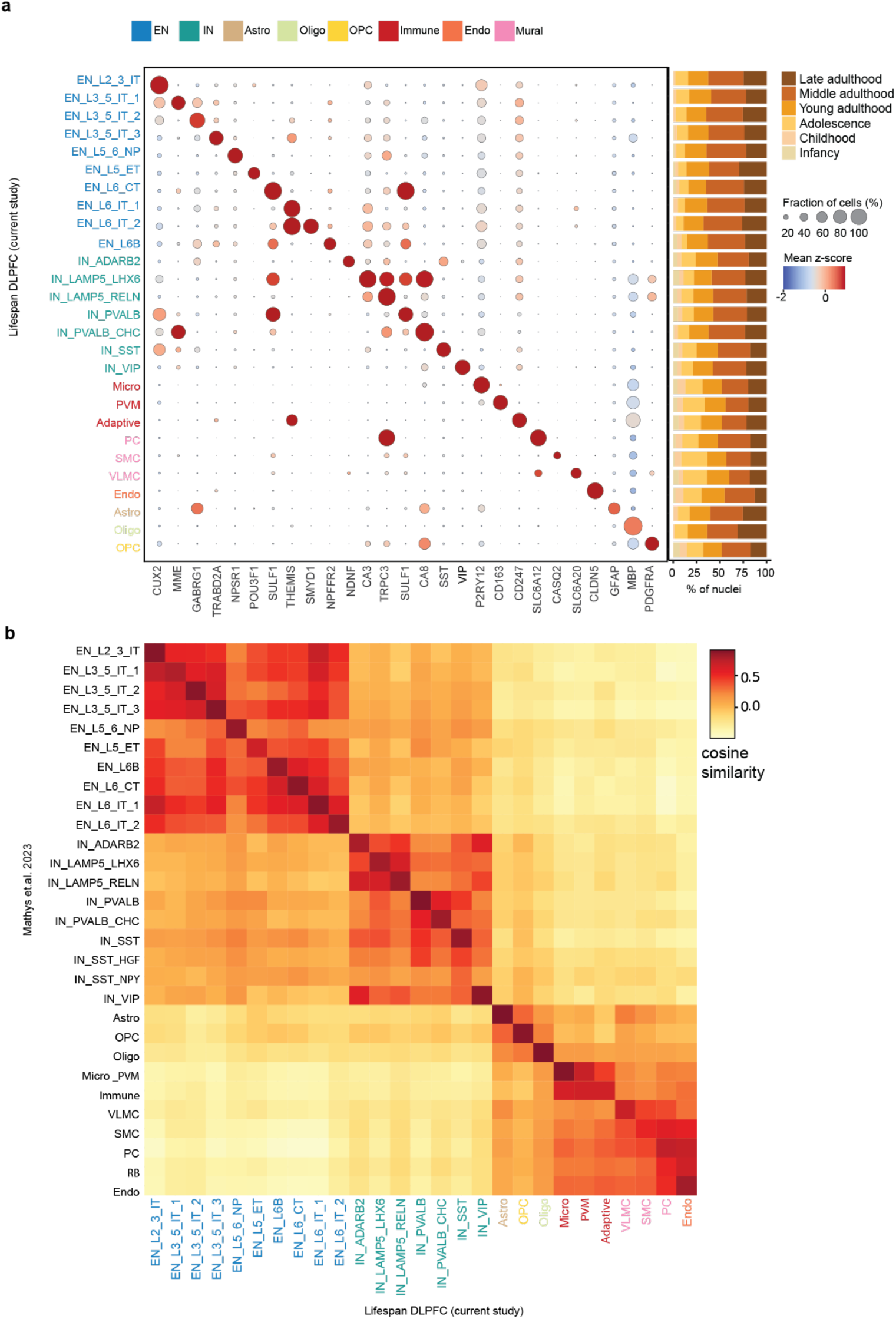
Subclass specific marker validation and cross-dataset similarity analysis. **a**, Dot plot showcasing selected key markers for each subclass, along with the fraction of nuclei observed across different age groups. **b**, Cosine similarity analysis comparing subclass-level cellular taxonomy of the lifespan DLPFC atlas (current study) with the Mathys et al. 2023^14^ dataset.

**Supplementary Figure 4.**
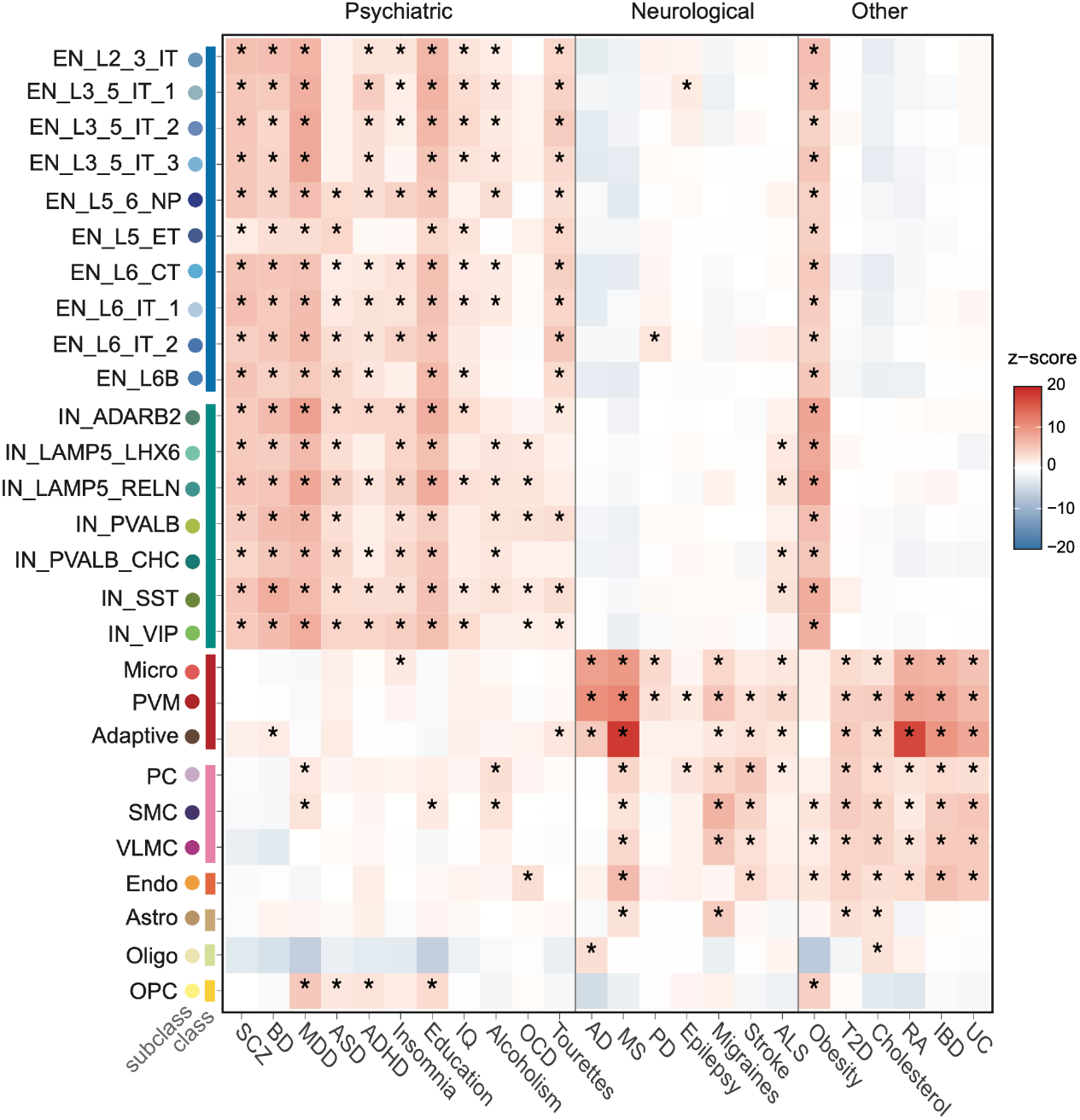
Polygenic diseases risk scores. Disease scores computed using the scDRS package^101^, depicted at the subclass level across various neurological, psychiatric and other selected GWAS traits, presented as the mean association z-score. “*”: indicates significance for enrichment in FDR-corrected P-value.

**Supplementary Figure 5:**
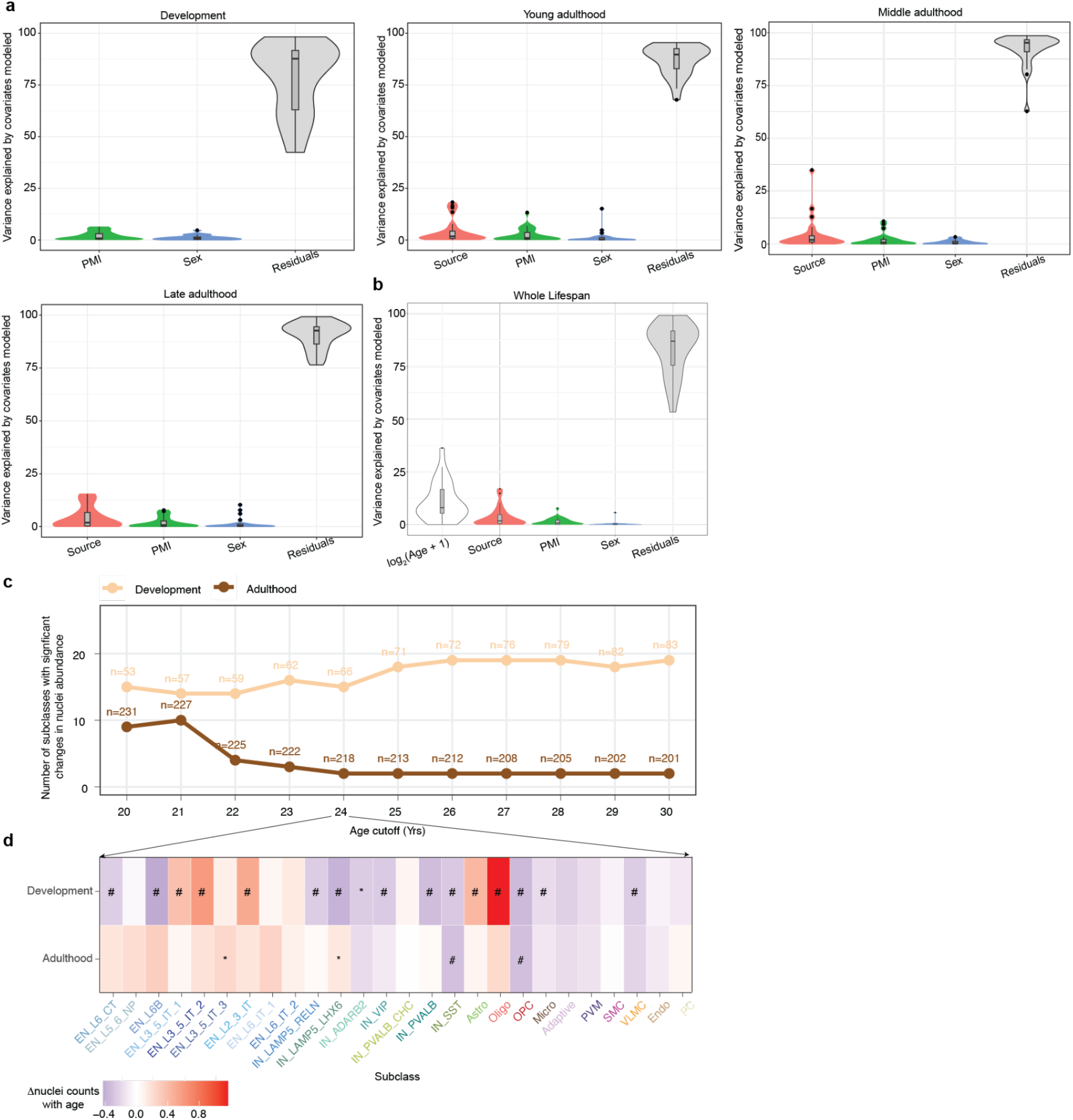
Variance explained by modeled covariates in nuclei abundances across the lifespan and per age group. **a-b**, Violin plots show the distribution of variance explained by individual covariates across all subclasses, as estimated using the variancePartition. Plots are shown for four age groups in a panel and the entire lifespan in b panel. **c**, Number of subclasses showing significant age-related changes in nuclei abundance (FDR < 0.05) as a function of different age cutoffs used to define “development” (< cutoff) vs “adulthood” (≥ cutoff). Sample sizes (n) for each group at each cutoff are added as text. **d**, Heatmap showing estimated effect size of age-associated changes in nuclei abundance across two groups when data are split at age 24 years. * and # indicate nominal p-value < 0.05 and FDR < 0.05, respectively.

**Supplementary Figure 6.**
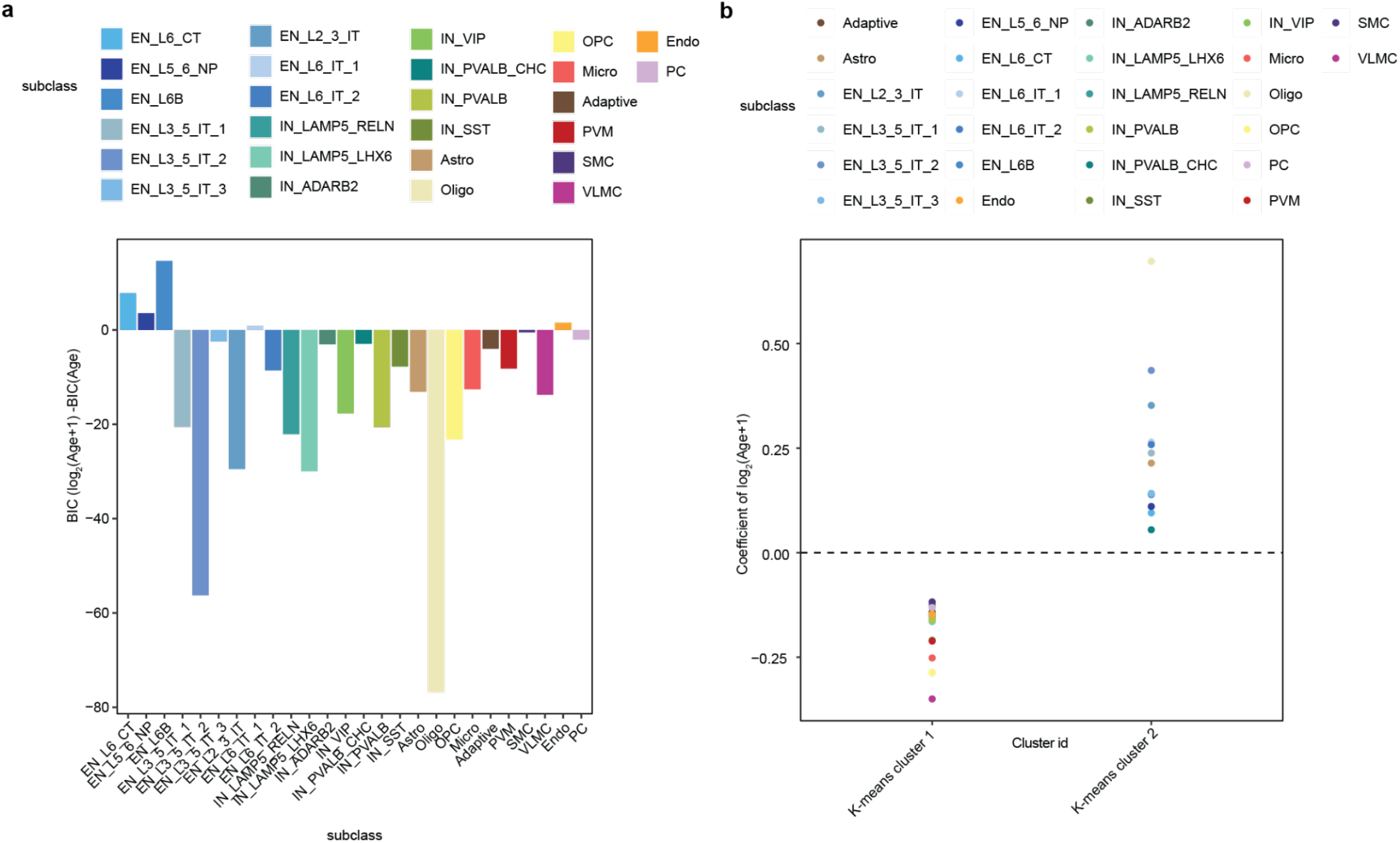
Optimal model selection for nuclei counts. **a**, Bayesian Information Criterion (BIC) obtained from modeling nuclei counts using the formulas: nuclei counts ∼ log_2_(Age+1) and nuclei counts ∼ Age, utilizing crumblr for each subclass. **b**, K-means clustering coefficient from nuclei counts ∼ log_2_(Age+1).

**Supplementary Figure 7.**
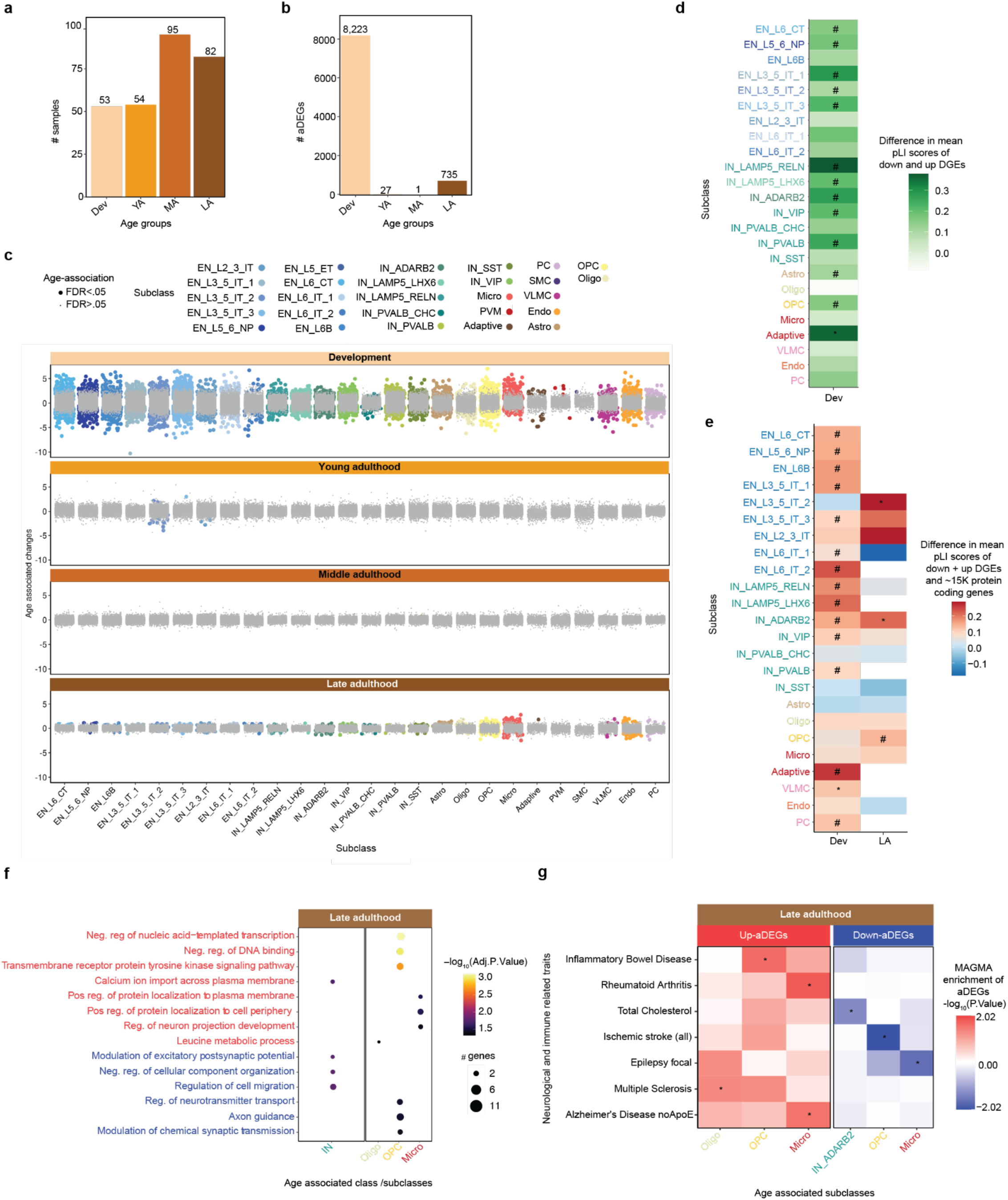
Age-associated gene expression across age groups. **a**, Number of samples and **b**, the number of DEGs with across four age groups. **c**, Beeswarm plot of age-associated effect sizes for 26 subclasses across four age groups. Gray dots are genes with FDR > 0.05 and colored dots are significant genes with FDR < 0.05. **d**, Heatmap showing the mean difference in pLI scores between down-aDEGs and up-aDEGs during development. Colors represent the direction and magnitude of mean pLI differences for each subclass. * indicates nominal p-value < 0.05 (Wilcoxon rank-sum test) and # indicates FDR-adjusted p-values < 0.05 across all subclasses. **e**, Heatmap to show differences in loss-of-function mutation scores (pLI) of down+up-aDEGs and ∼15K protein coding genes during development and late adulthood. * and # indicate nominal p-value < 0.05 and FDR < 0.05 from Wilcoxon test. **f**, Functional pathway analysis of up and down-aDEGs during late adulthood for subclasses that were significantly enriched for GO biological processes with an adjusted p-value < 0.05. **g**, Association of up and down-aDEGs during late adulthood with risk genes for neurological and immune-related traits using MAGMA.* and # indicate nominal p-value < 0.05 and FDR < 0.05 from MAGMA enrichment.

**Supplementary Figure 8.**
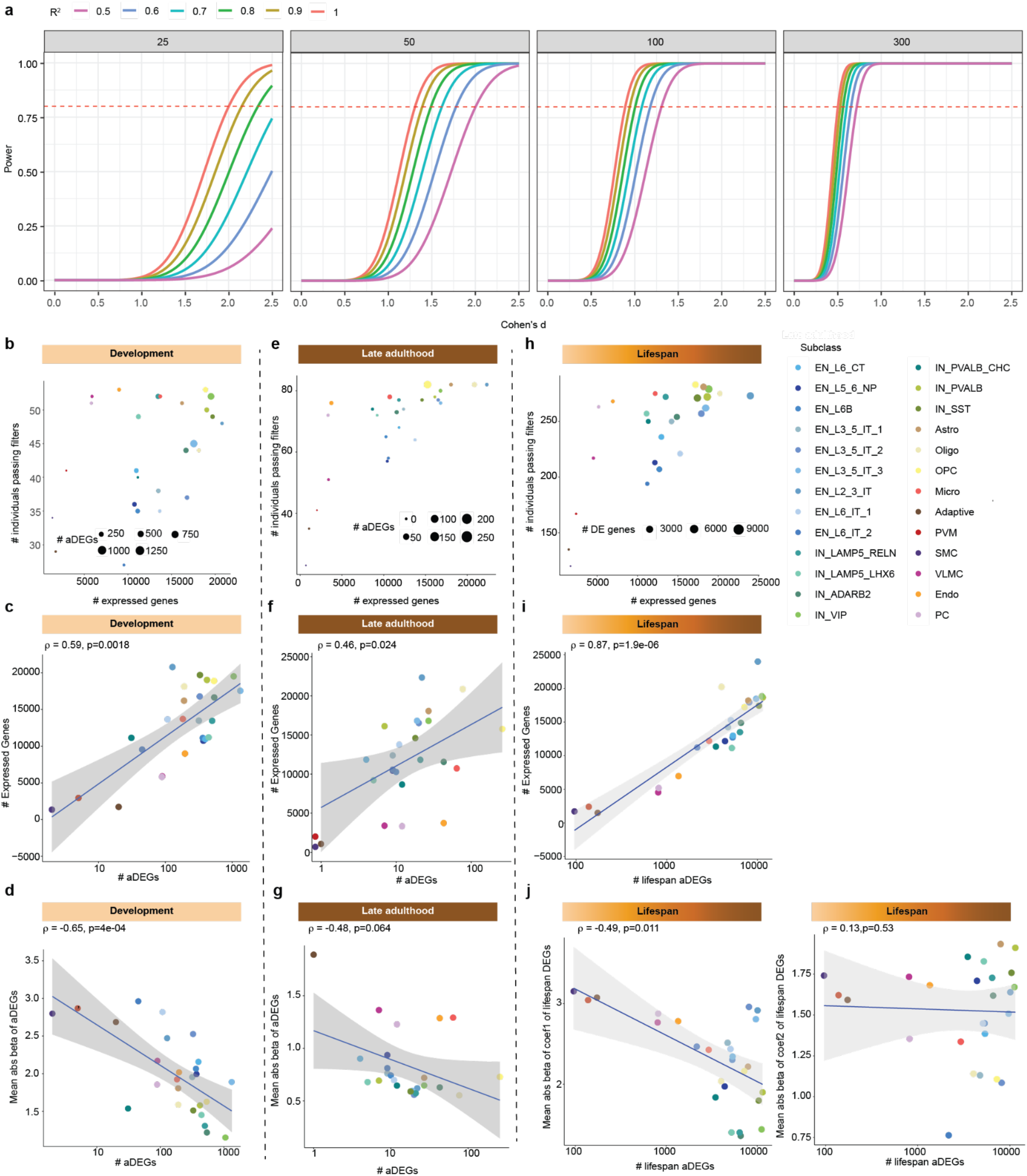
Theoretical and empirical assessment of statistical power across subclasses. **a**, Theoretical power curves illustrating the relationship between effect size (Cohen’s *d*) and statistical power across sample sizes (*n* = 25, 50, 100, 300) and moderated *R*^*2*^ values (0.5–1.0). The horizontal dashed line indicates the 80% power threshold. **b**, Scatter plots showing the number of expressed genes and number of developmental donors passing quality control for each subclass (derived from dreamlet pseudobulk residuals). **c–d**, Spearman correlations between aDEG developmental counts and (c) number of expressed genes and (d) mean absolute age-associated expression changes during development per subclass. **e**, Same as panel b, but based on late adulthood donors. **f–g**, Spearman correlations between late adulthood aDEG counts and (f) number of expressed genes and (g) mean absolute age-associated expression changes during late adulthood per subclass. **h**, Same as panel b, but based on lifespan donors. **i–j**, Spearman correlations between lifespan aDEG counts and (i) number of expressed genes and (j) mean absolute age-associated expression changes during lifespan per subclass. In panel j, the left plot shows correlation with the linear age coefficient (coef_1_), and the right plot shows correlation with the non-linear (coef_2_) coefficient. Points in b, e and h are colored by subclass, and point size reflects the number of aDEGs during development, late adulthood and lifespan respectively.

**Supplementary Figure 9.**
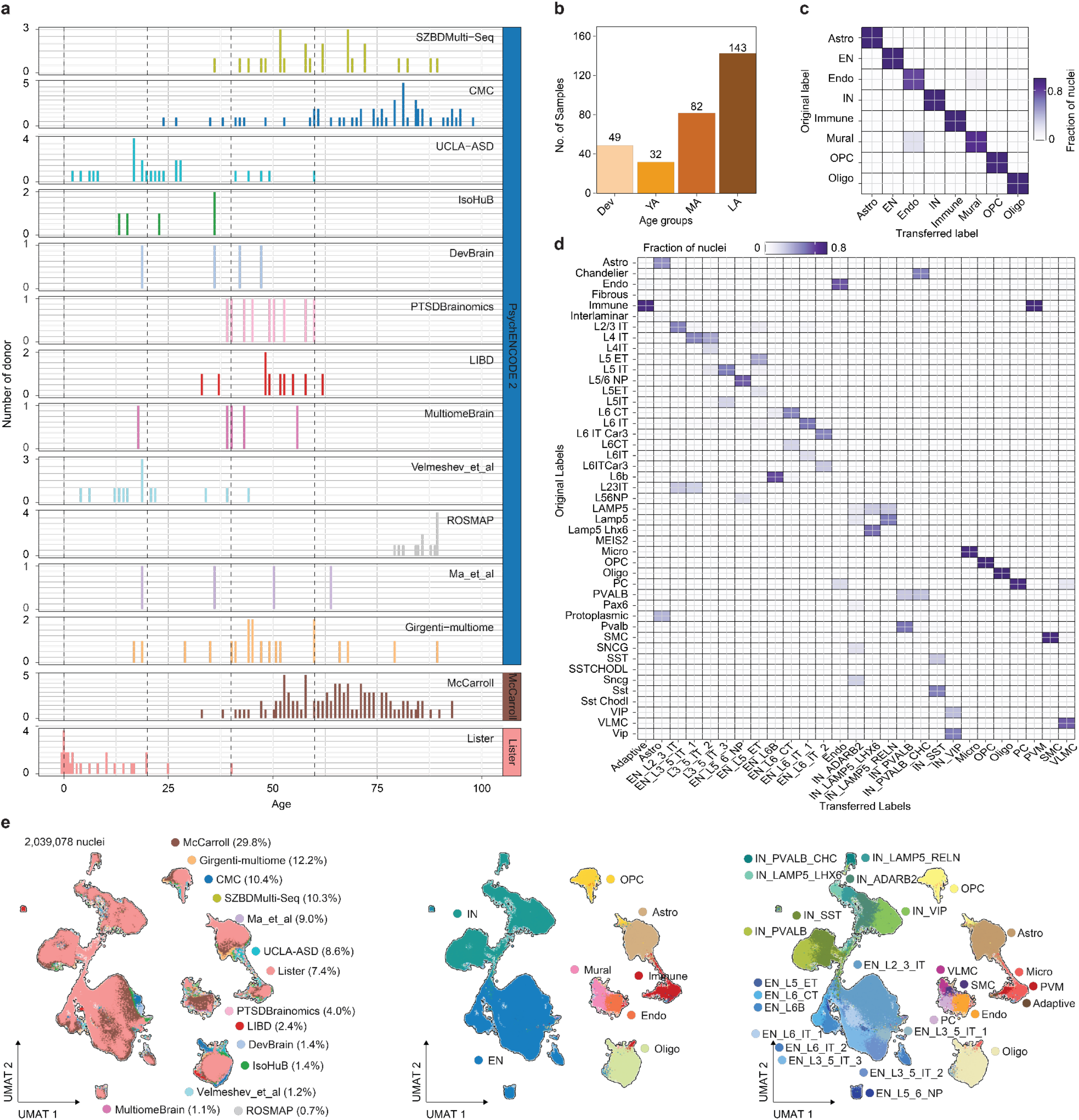
Lifespan replication dataset: **a**, Age distribution of donors (x-axis) across 14 published cohorts (y-axis) included in the lifespan replication dataset across full human lifespan. **b**, Number of donor samples per age group included in the replication dataset. **c-d**, Concordance of class in c and subclass-level label transfer in d. Each cell shows the fraction of nuclei with a given original label assigned to each transferred class in c and subclass in d label confirming reproducibility of fine-grained annotations across datasets. **e**, UMAP plots of over 2 million nuclei from the replication dataset, colored by dataset of origin with the percentage of total nuclei contributed by each dataset (left), six broad cell types (middle), and subclass labels (right).

**Supplementary Figure 10.**
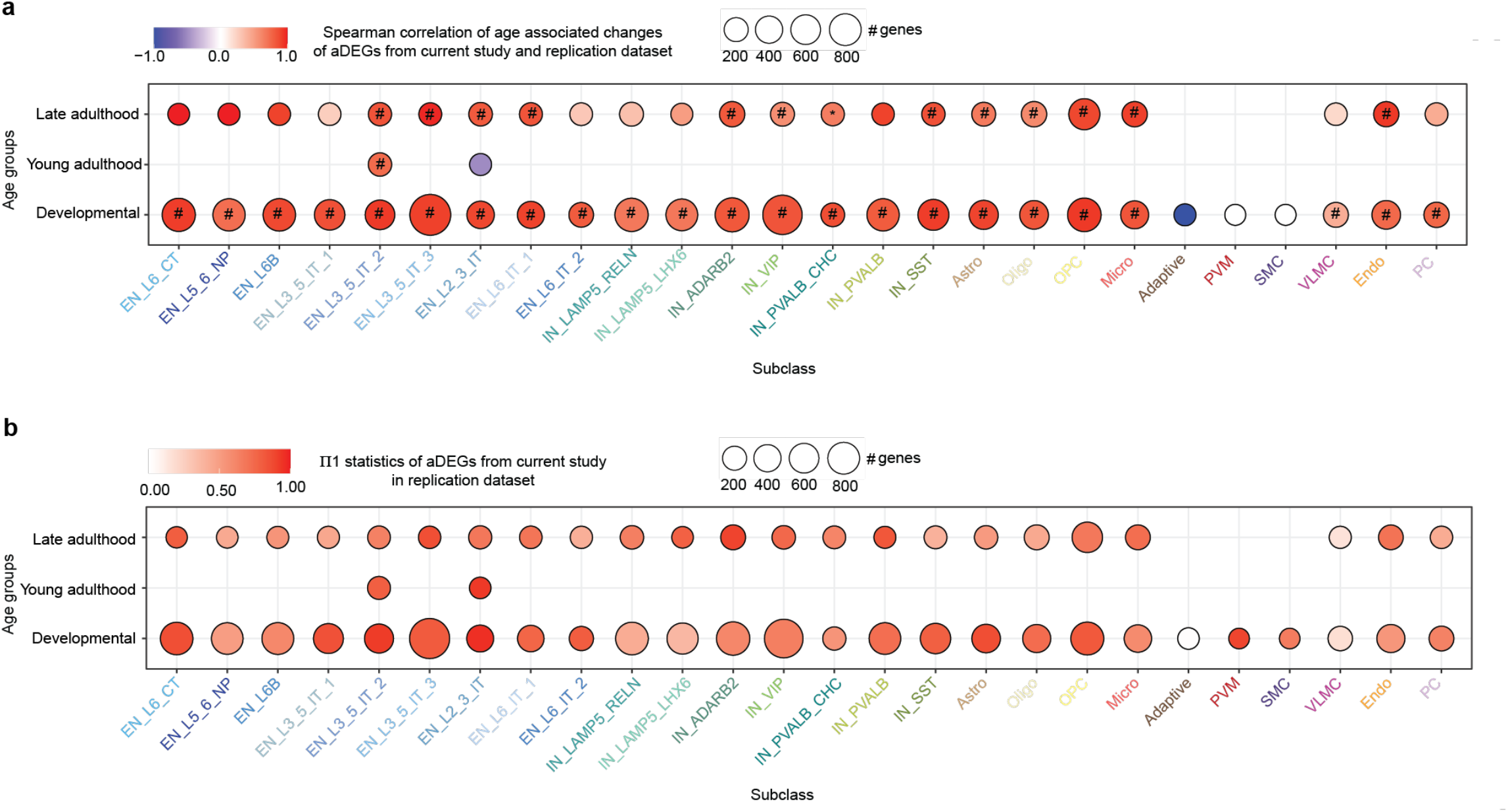
Replication of age-associated gene expression changes across subclasses and age groups. **a**, Bubble plot displaying the Spearman correlation of age-associated differentially expressed genes (aDEGs; FDR < 0.05) between the DLPFC lifespan and the replication dataset. Dot size corresponds to the number of shared aDEGs, and color represents the spearman correlation. **b**, Bubble plot showing the π1 statistics (estimate of replication rate) of aDEGs from the DLPFC lifespan dataset in the replication dataset. Dot size reflects the number of aDEGs tested, and color intensity indicates the replication signal (π1 ranges from 0 to 1). In all panels: “*” indicates spearman correlation p-value < 0.05 and “#” indicates significance after multiple testing correction across all subclasses.

**Supplementary Figure 11.**
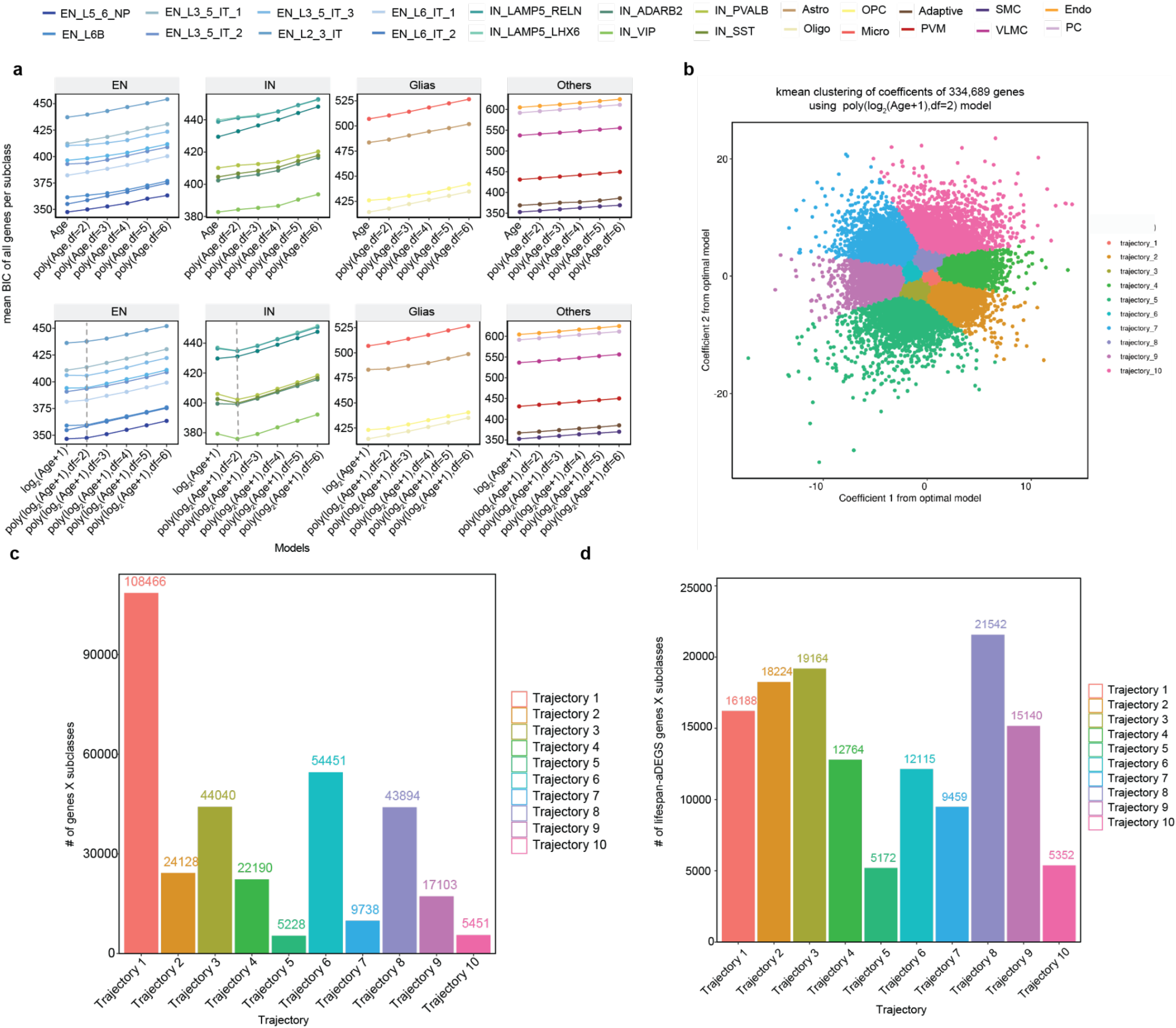
Optimal model selection and clustering of gene expression trajectories. **a**, Mean BIC of linear and non-linear models fitting on all genes. X-axis displays all models implemented in the pipeline. The dashed line shows the optimal model used for final clustering of trajectories. **b**, k means clustering of non-linear coefficients from the optimal model of 334,689 genes across all subclasses. **c-d**, Bar plot to show all genes (*n* = 334,689) and lifespan aDEGs (*n* =135,120) after FDR < 0.05 from all subclasses within each trajectory cluster.

**Supplementary Figure 12.**
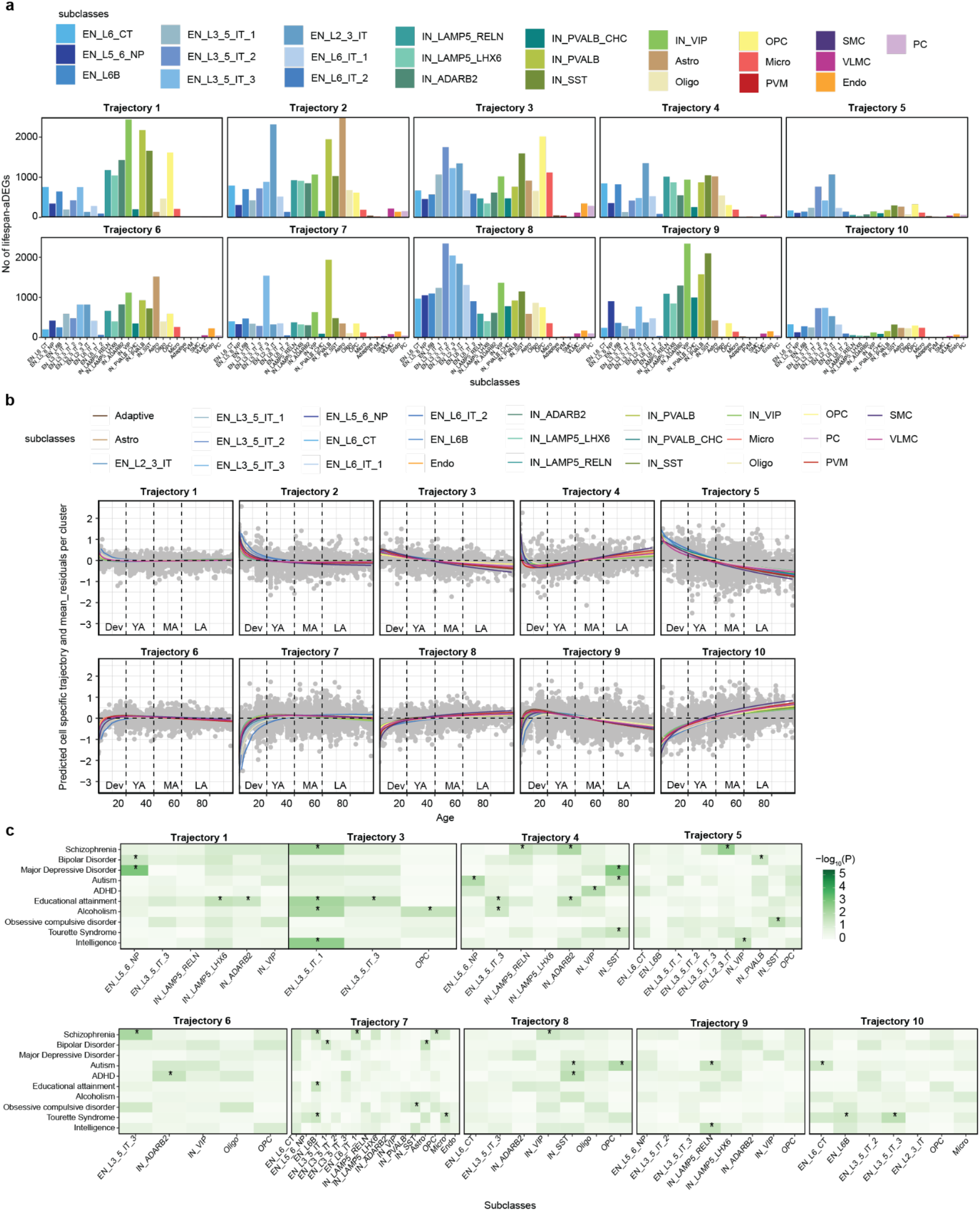
Subclass specificity of trajectories. **a**, Distribution of lifespan-aDEGs across 27 subclasses stratified by ten clusters. **b**, Plot of covariates corrected gene expression as a function age shown as gray points and fitted polynomial using the mean of coefficients from each subclass. The polynomial fit is shown in subclass specific colours. **c**, Association of subclass specific development aDEGs stratified by clusters of trajectories with risk genes for brain-related traits using MAGMA. * and # indicate nominal p-value < 0.05 and FDR < 0.05 from MAGMA enrichment. * and # indicate nominal p-value < 0.05 and FDR < 0.05 from MAGMA enrichment.

**Supplementary Figure 13.**
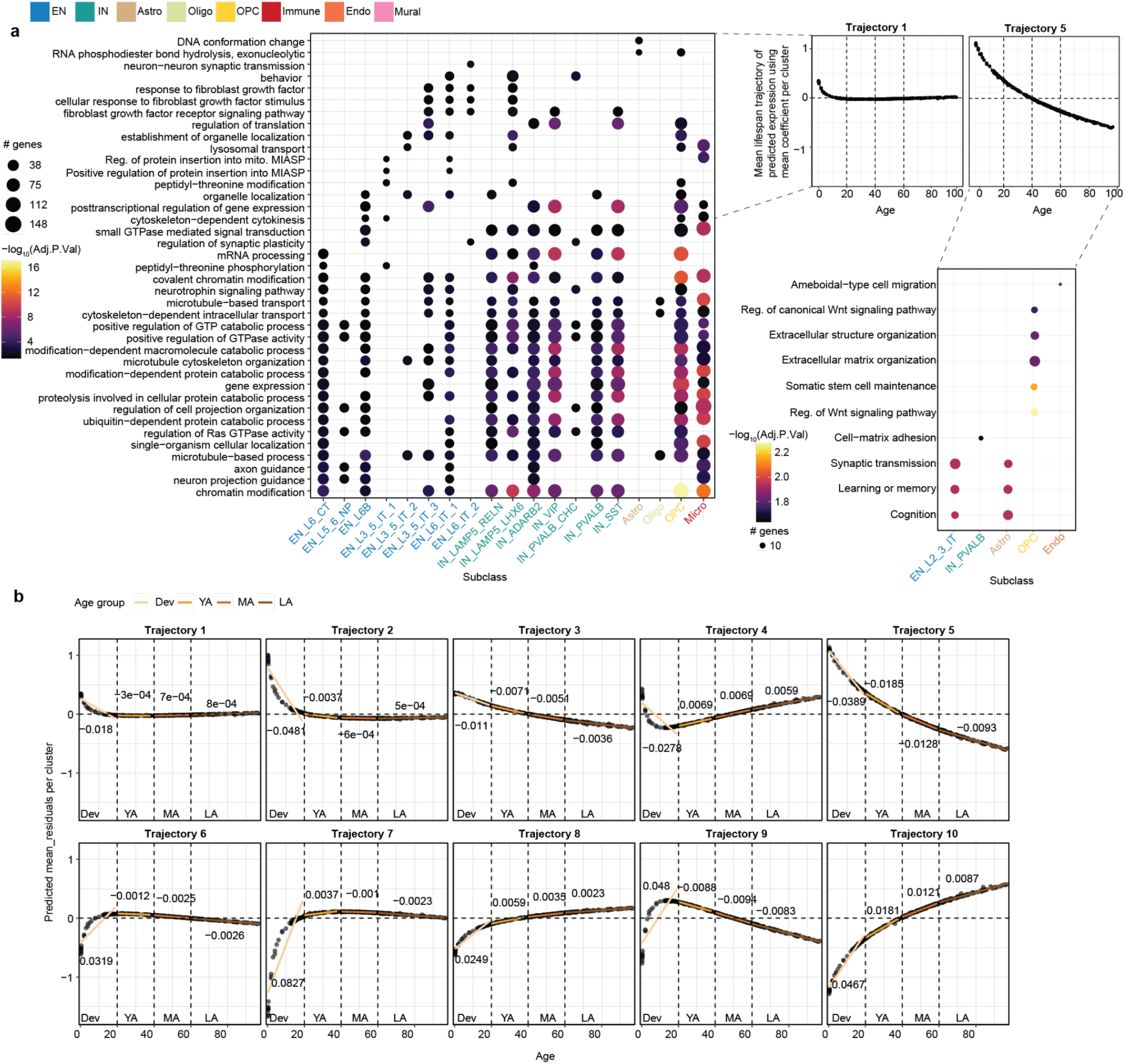
Biological mechanism exhibited by trajectories. **a**, Functional pathway analysis of subclass-specific genes in trajectory 1 (left) and trajectory 5 (right), highlighting subclasses significantly enriched for GO biological processes with an adjusted p-value < 0.05. **b**, Plot of predicted trajectory using the mean of coefficients of all genes within each trajectory. The linear fit across age groups shows the regression line from average trajectory to age from each age group. The text above the age-groups specific fitted line figure shows the beta from model: lm(expression∼Age+covariates). All beta values had p-value < 2e-16.

**Supplementary Figure 14.**
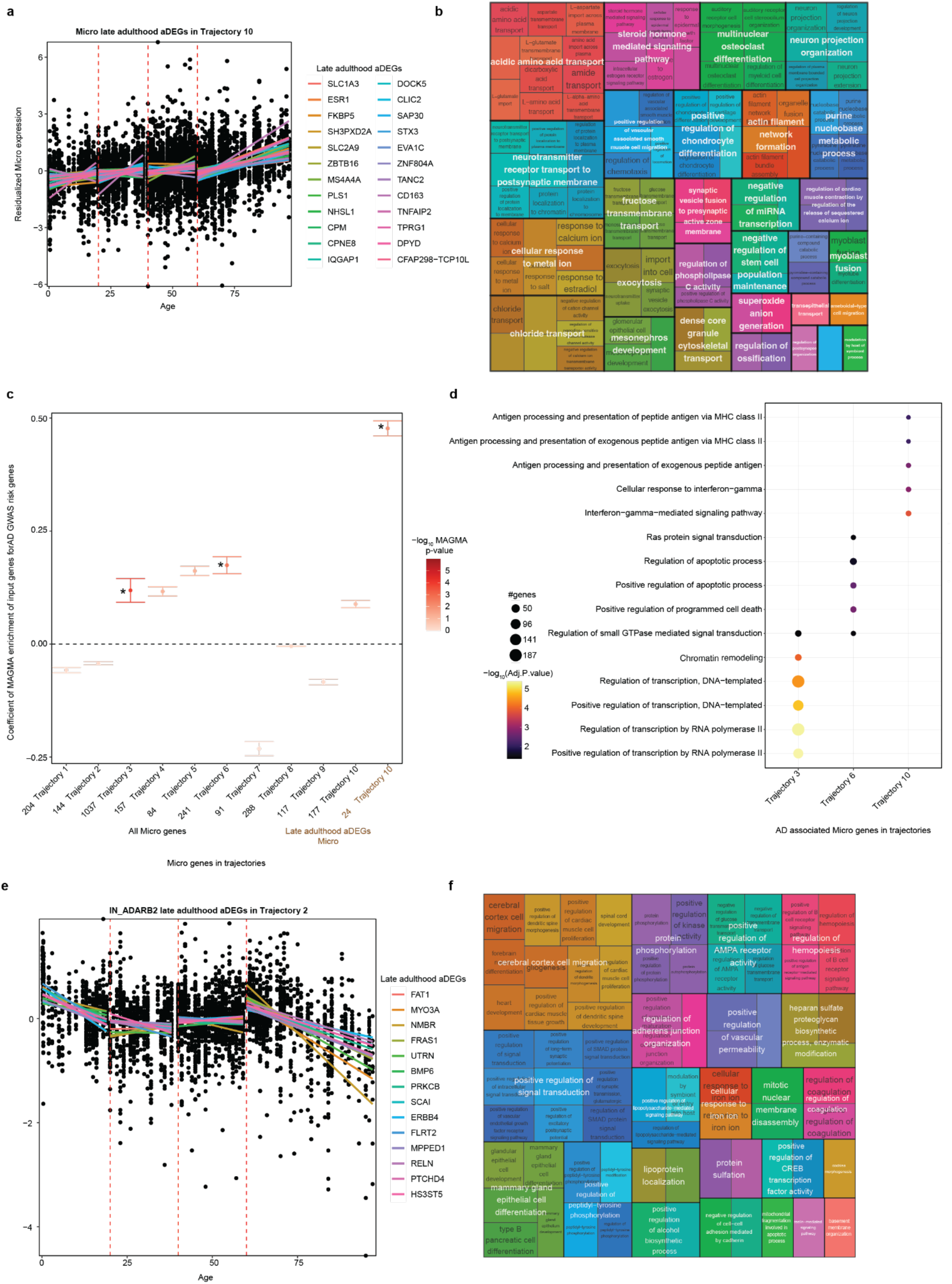
Late adulthood IN_ADARB2 and Micro aDEGs. **a**, Plot of Micro specific aDEGs residualized gene expression from trajectory 10 shown as black markers. Age related changes of every gene is shown as a colored regression line within each group. **b**, Functional pathway analysis of micro-specific aDEGs from trajectories 10 was conducted using rrvgo. The input GO IDs were obtained from enrichR analysis of aDEGs with an adjusted p-value < 0.10 for microglia. **c**, The coefficient of enrichment of AD GWAS risk genes for all micro genes from the 10 trajectories and late-adulthood specific aDEGs in trajectory 10 is shown in brown. **d**, Functional pathway analysis of all micro genes from trajectories 3, 6, and 10, which were significantly enriched for GO biological processes with an adjusted p-value < 0.05, is also included. **e**, Plot of IN_ADARB2 specific aDEGs residualized gene expression from trajectory 2 shown as black markers. Age related changes of every gene is shown as a colored regression line within each group. **f**, Functional pathway analysis of IN_ADARB2 aDEGs from trajectories 2 was conducted using rrvgo. The input GO IDs were obtained from enrichR analysis of aDEGs with an adjusted p-value < 0.05.

**Supplementary Figure 15.**
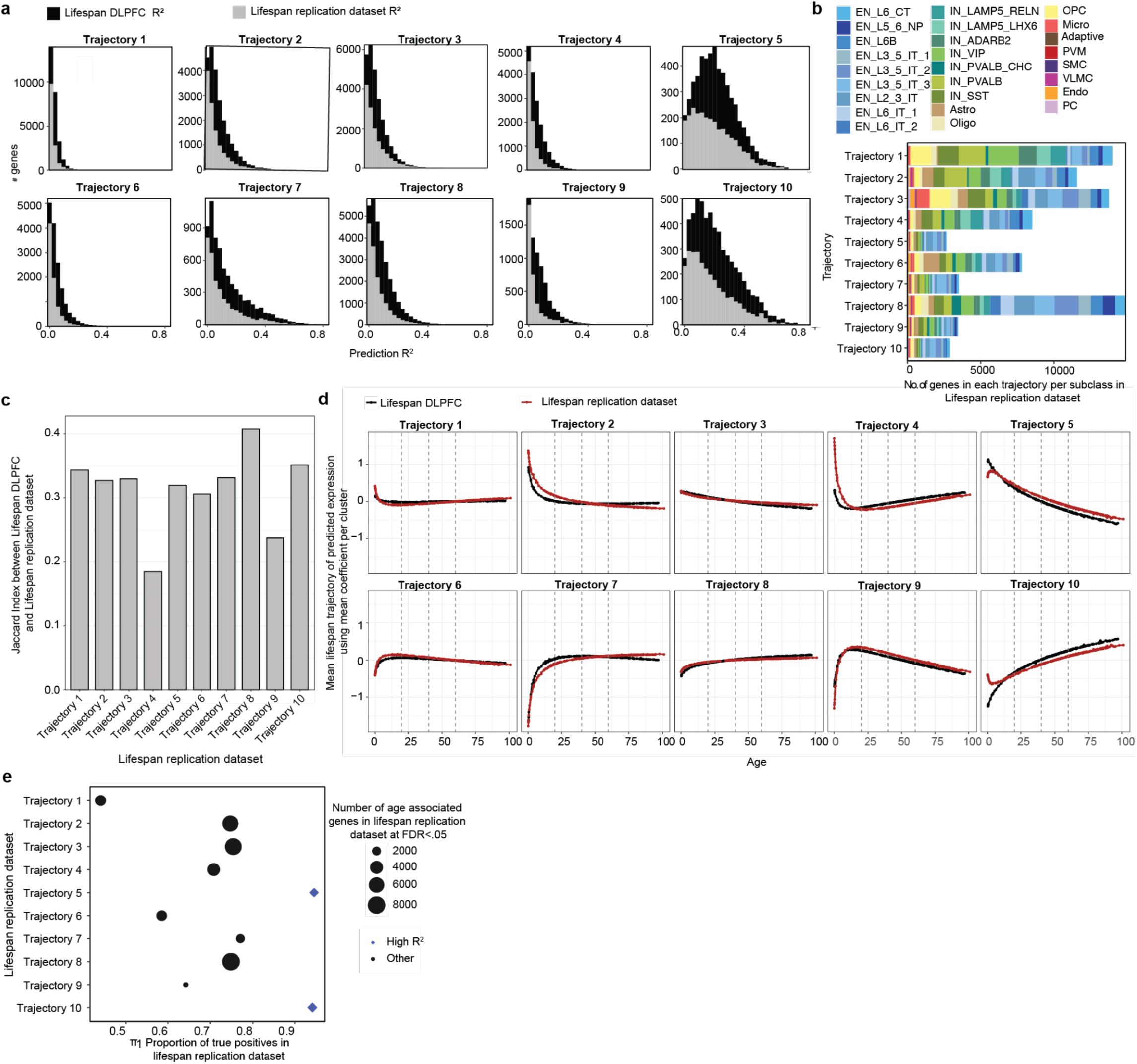
Replication of lifespan trajectories. **a**, Distribution of gene-level R^2^ values comparing predicted versus observed expression across all genes in each trajectory. R^2^ values are shown for both the discovery lifespan DLPFC dataset (black) and the lifespan replication dataset (gray). **b**, Barplot showing the number of genes per trajectory stratified by subclass in the Lifespan replication dataset using strategy two.**(c–e)** Trajectory patterns derived independently by re-running the full trajectory pipeline (Fig. 2a) on the Lifespan replication dataset **c**, Jaccard index between discovery and replication trajectory gene sets, summarizing the degree of gene-level overlap for each trajectory. **d**, Mean trajectory fits for each trajectory across donor age. Predicted expression trends from the discovery dataset (black) and the replication dataset (red). **e**, Scatterplot showing replication performance across trajectories. The x-axis shows the proportion of true positives (π_1_) from the discovery dataset that were recovered in the replication dataset. Dot size corresponds to the number of replicated age-associated genes at FDR < 0.05. Blue diamonds indicate trajectories with high R^2^ in both datasets (Trajectories 5 and 10).

**Supplementary Figure 16.**
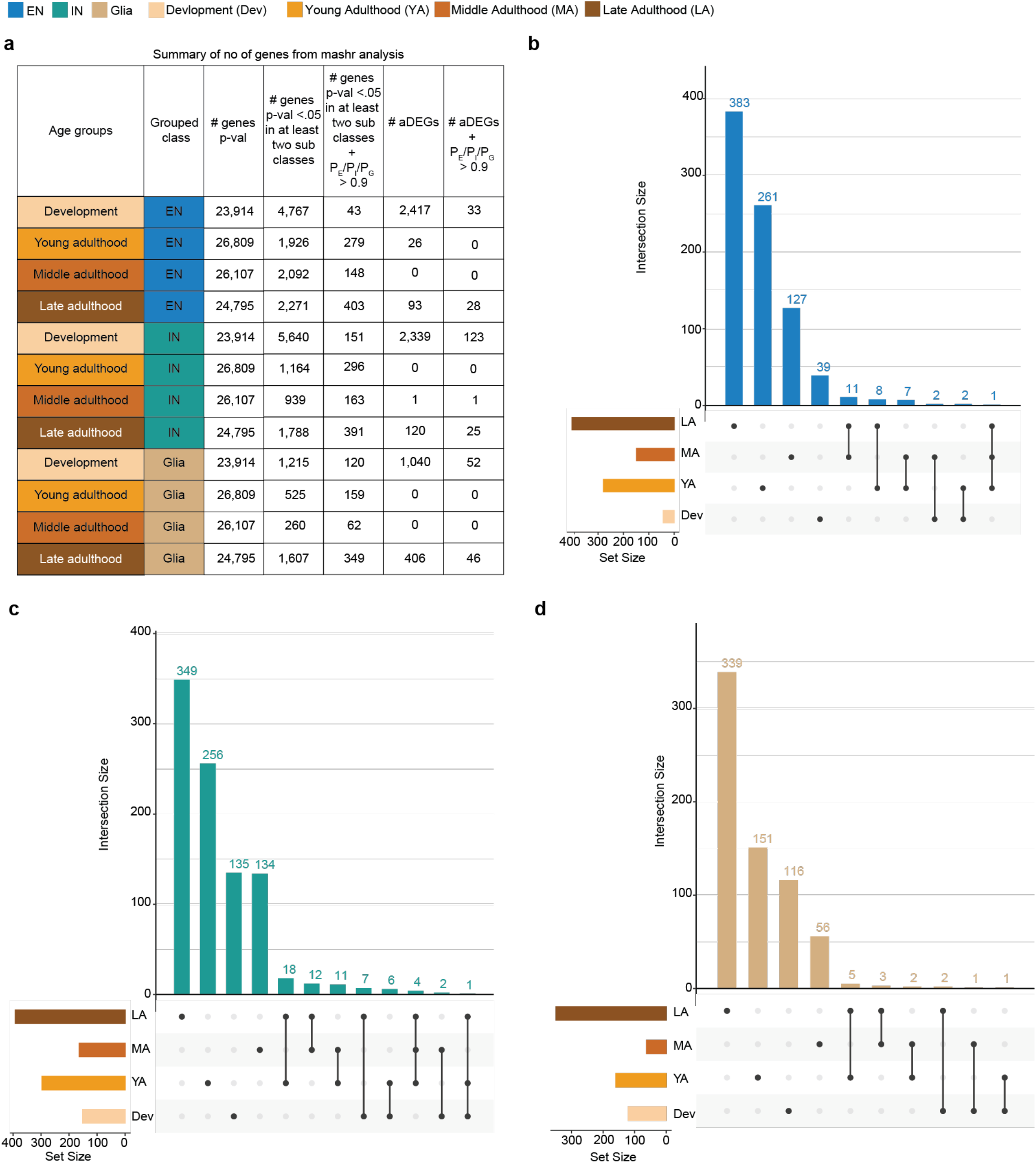
Summary of number of shared genes from *mashr* analysis. **a**, Table displaying the total number of genes to run the *mashr* analysis and shared genes at composite probability > 0.9. Table also displays the counts of genes that had age effect size at significant p-value < 0.5 in at least two subclasses. **b-d**, Upset plot to show the overlap of shared genes at composite probability > 0.9 across age groups in EN, IN and Glia grouped classes.

**Supplementary Figure 17.**
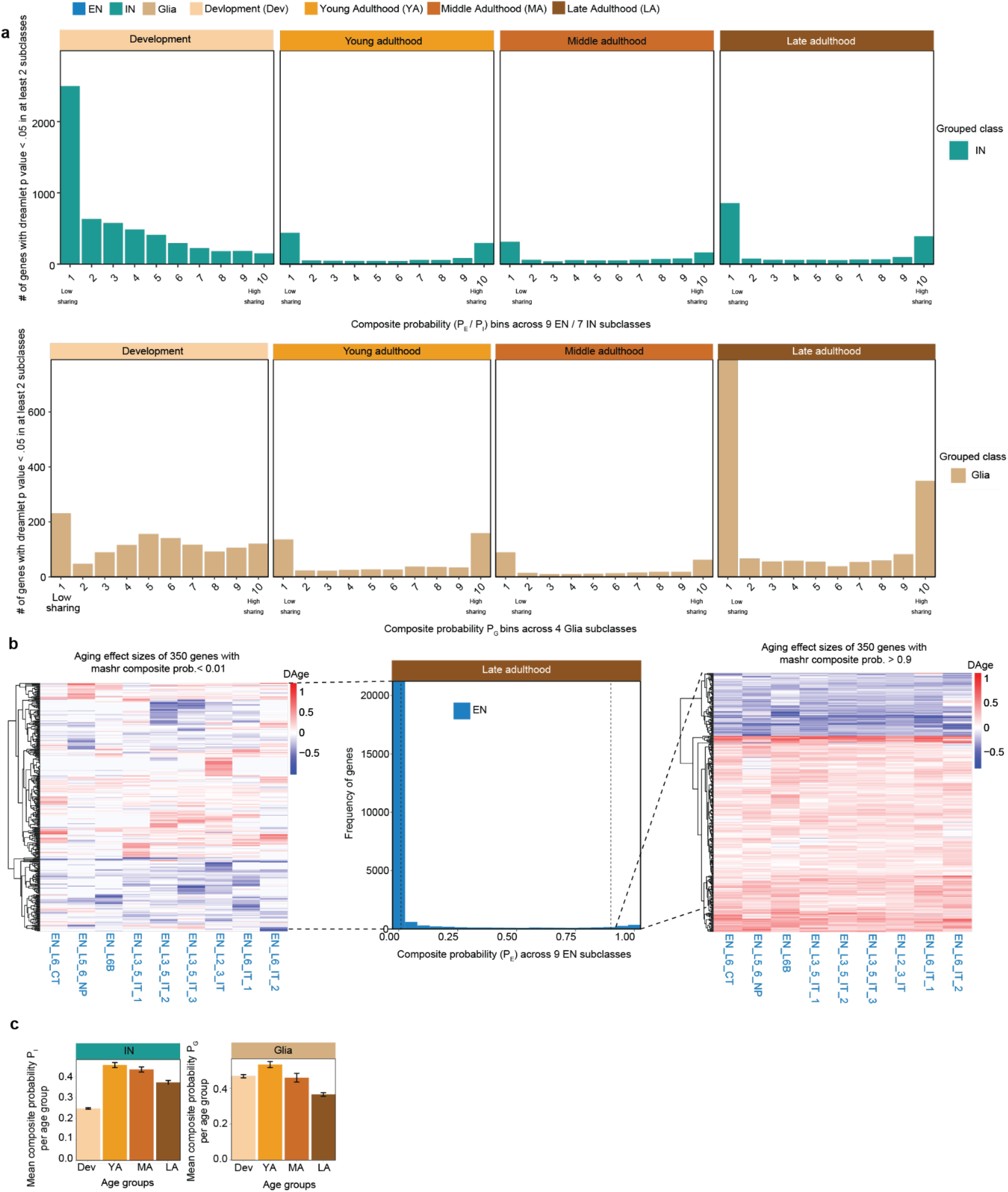
Degree of sharing across subclasses. **a**, Bar plot displaying the number of genes stratified into 10 equally sized bins based on composite probability (P_I_ and P_G_) values for IN and glia respectively, ranging from 0 to 1 from development to late adulthood group. The number of input genes to make these bar-plots is displayed in Fig. S16a. **b**, Heatmap of age associated effect sizes across 9 EN for genes with composite probability (P_E_ < 0.01 in left) and (P_E_ > 0.9 in right) across late adulthood. **c**, Mean P_I_ (left) and P_G_ (right) of genes in each age group. This plot is the mean of probabilities of genes shown in Fig. S17a.

**Supplementary Figure 18.**
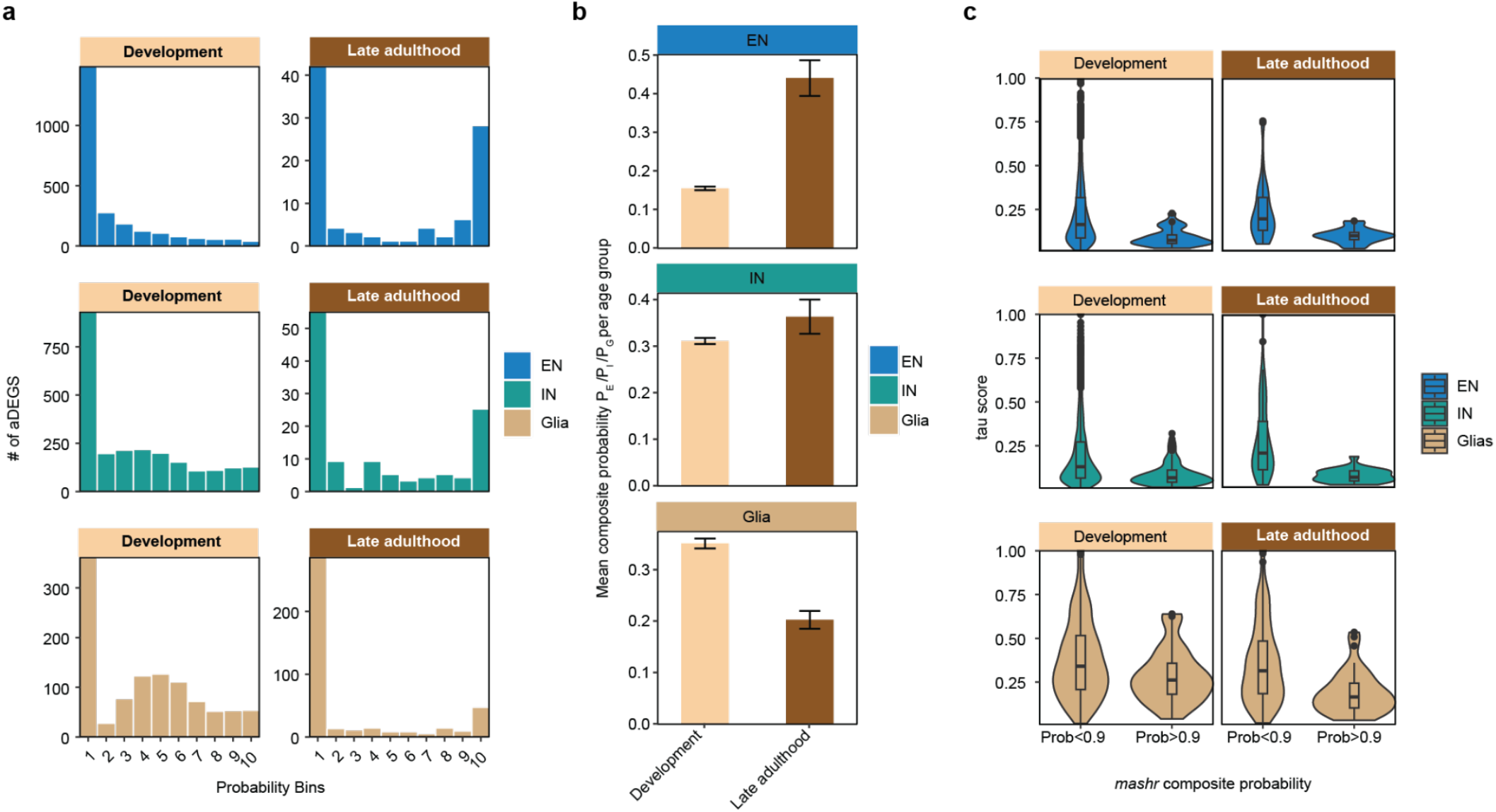
Composite probability of aDEGs. **a**, Bar plot displaying the number of aDEGs stratified into 10 equally sized bins based on composite probability (P_E_, P_I_, P_G_) values for EN, IN and glia ranging from 0 to 1 for development and late adulthood groups. **b**, Mean P_E_, P_I_, P_G_ of all development and late adulthood aDEGs. **c**, Violin plot of tau scores of genes that have age associated with nominal p-value from dreamlet tool in at least two subclasses. The x-axis is stratified by genes with P_E_, P_I_, P_G_ < 0.9 and > 0.9.

**Supplementary Figure 19.**
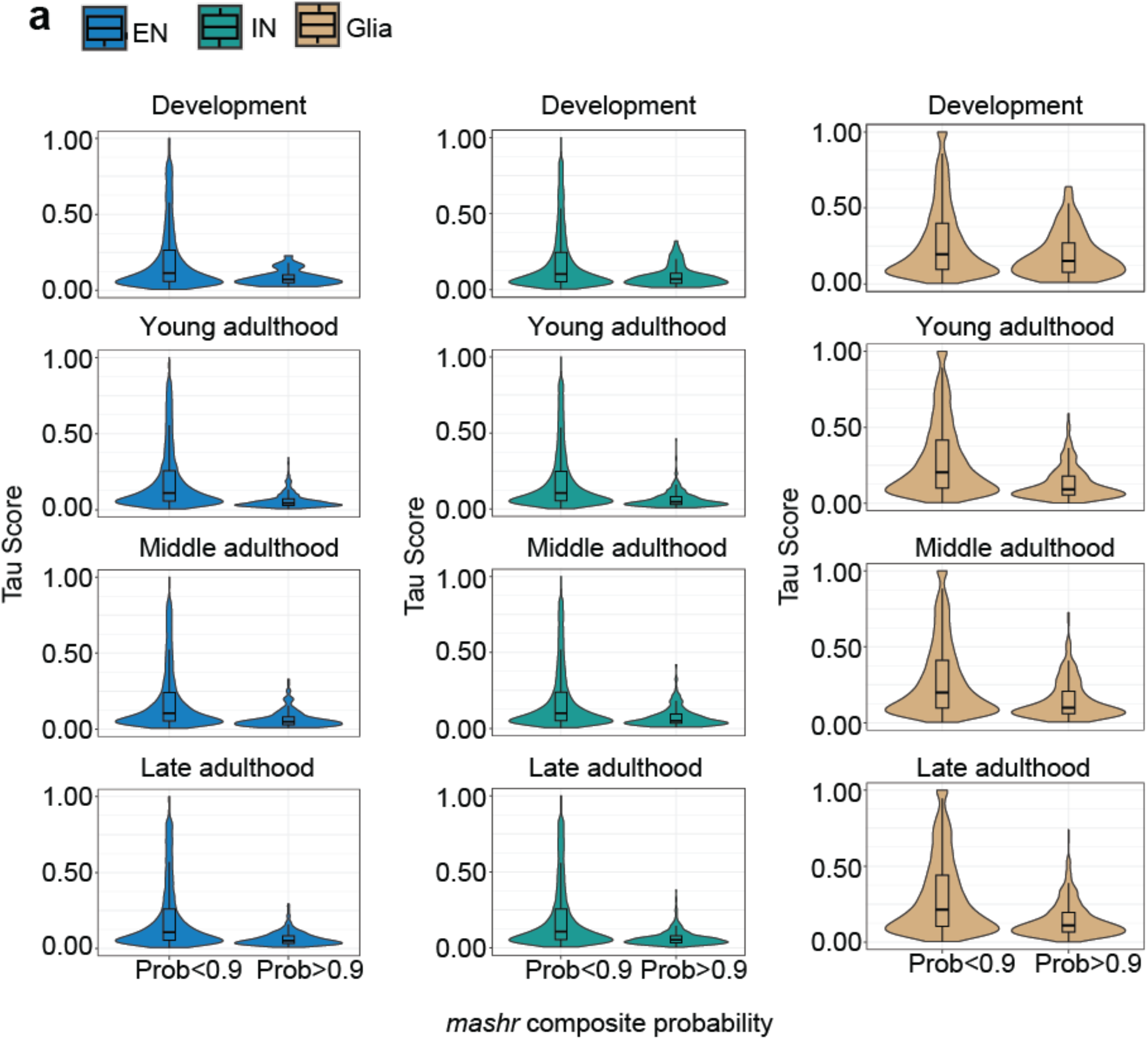
Distribution of cell specificity tau score. **a**, Violin plot of tau scores of genes that have age associated with nominal p-value from dreamlet tool in at least two subclasses. The x-axis is stratified by genes with P_E_, P_I_, P_G_ < 0.9 and > 0.9. These tau scores were obtained from 9 EN and 7 IN and 4 Glia expressions separately for each age group.

**Supplementary Figure 20.**
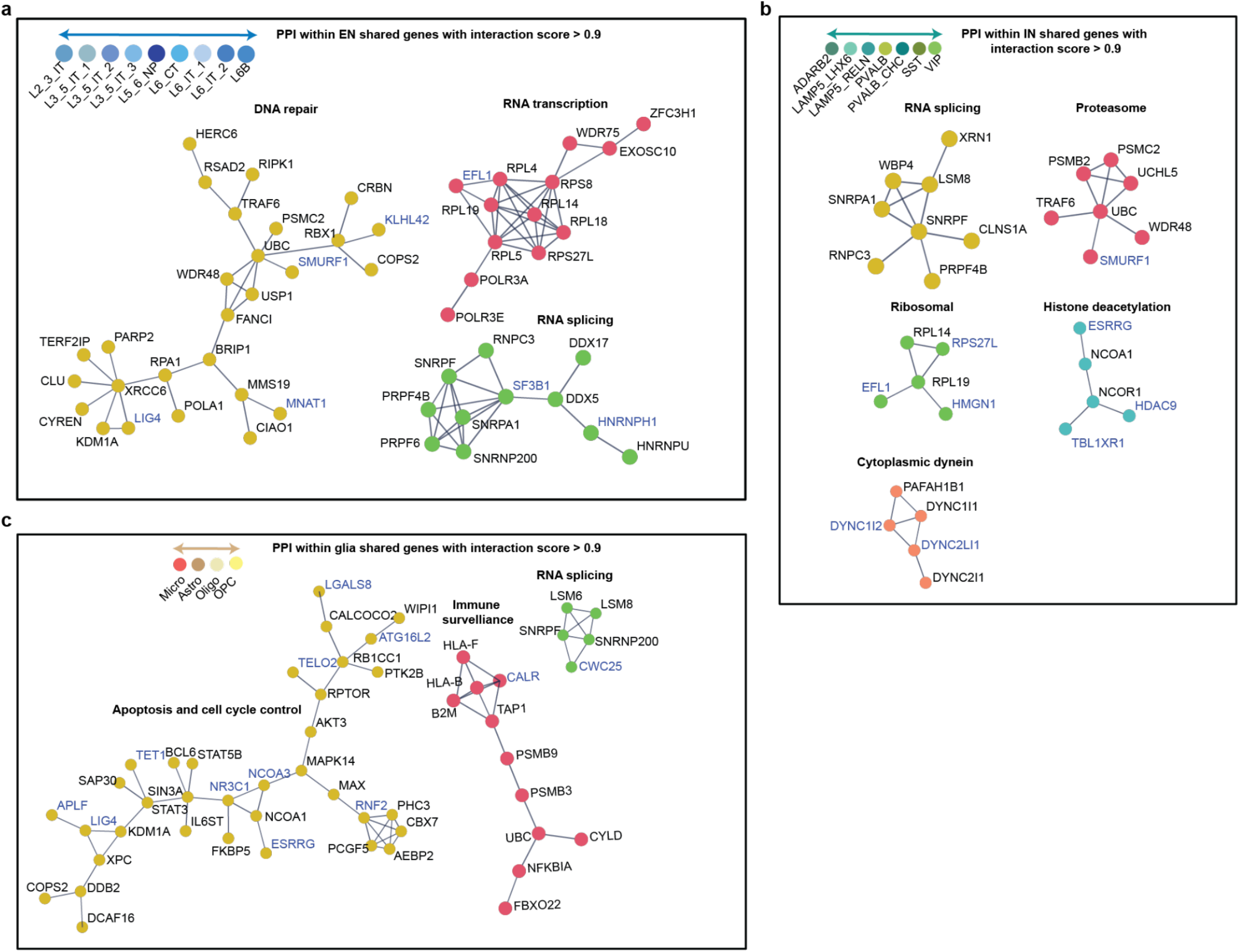
Protein-protein interactions among shared genes. **a-c** Identified clusters of networks of genes that show significant PPI interactions with score > 0.9 obtained from the analysis of shared genes within 9 EN, 7 IN and 4 glia classes. Genes highlighted in blue indicate downregulation with age and remaining are upregulated. EN clusters in b include genes crucial for 1) DNA repair, 2) RNA transcription, and 3) RNA splicing-specific complexes colored as gold, pink and mantis respectively. IN clusters in c include 1) spliceosome assembly, 2) the proteasome complex, 3) the histone deacetylase complex, and 4) components for maintaining cellular function and intracellular transport colored as yellow, pink, mantis, sea blue and salmon respectively. Glia clusters in d include 1) proteins involved in apoptosis and cell cycle control, 2) the MHC class I protein complex and peptide loading complex, and 3) RNA processing and splicing complexes colored as yellow, pink and mantis respectively.

**Supplementary Figure 21.**
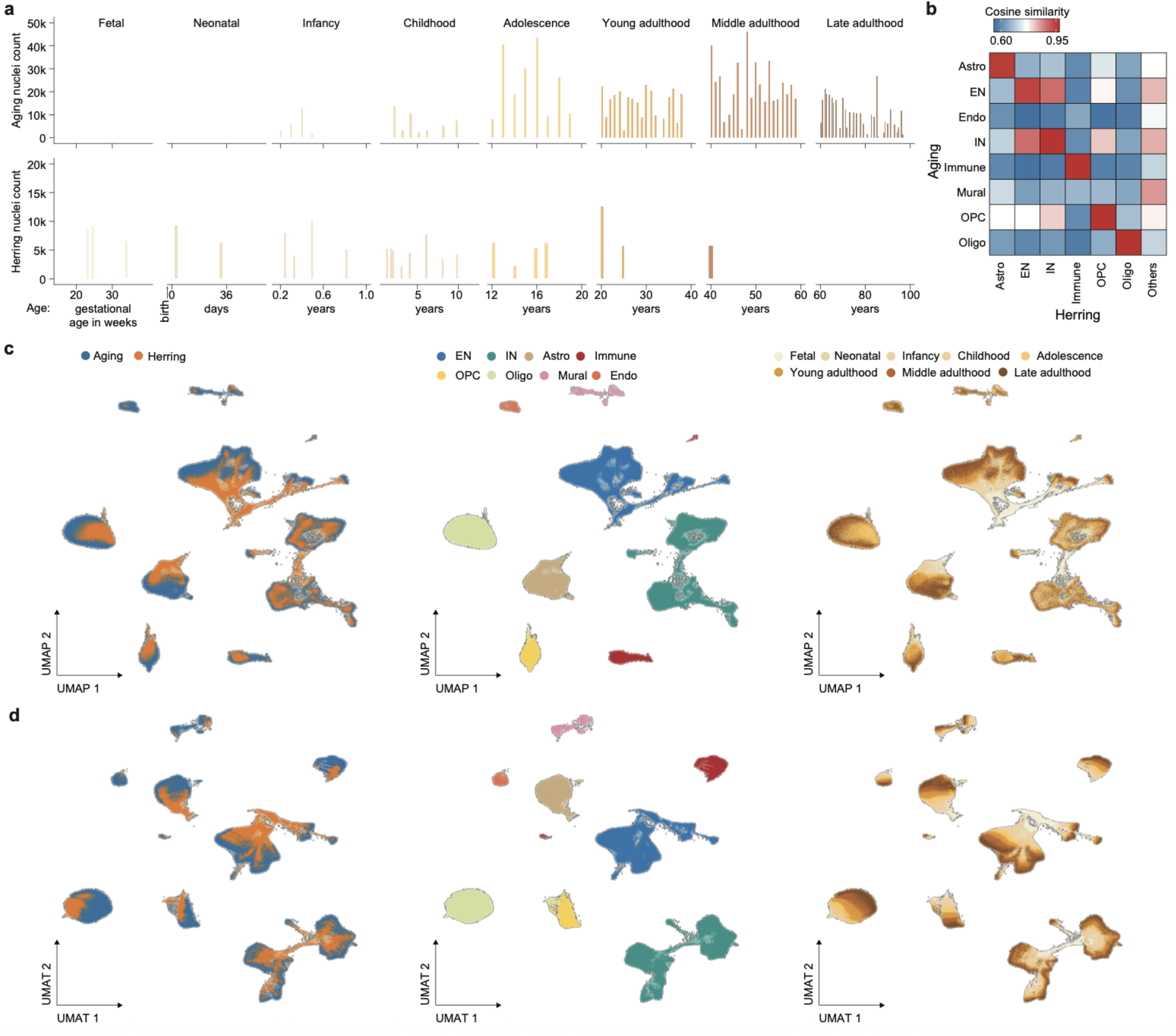
Characteristics and similarity of the combined snRNA-seq dataset. **a**, Nuclei counts in current (top) and previous (bottom) snRNA-seq datasets of the human DLPFC, categorized by age group and color-coded. **b**, Cell type similarity between the two datasets at the class level. **c,d**, Integration of the current dataset with previously published data. UMAP (**c**) and UMAT (**d**) representations of the combined snRNA-seq dataset colored by dataset (left), class (middle), and stage (right).

**Supplementary Figure 22.**
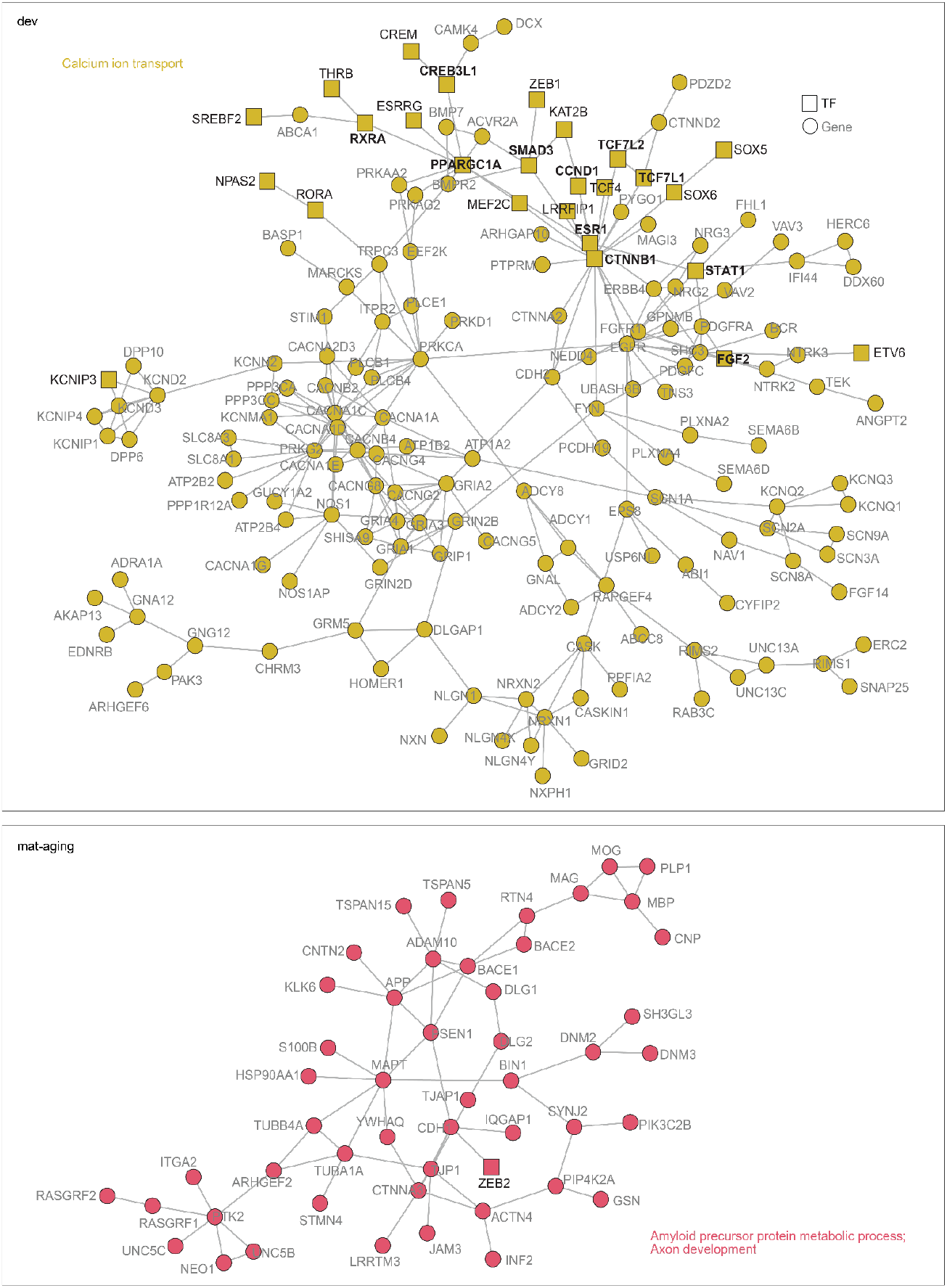
Regulatory networks for traDEG modules in oligodendrocytes. TF-gene regulatory networks inferred for Oligo traDEG modules, with enriched biological processes annotated based on network composition. Squares indicate TFs; circles indicate target genes. TF labels are shown in black; gene labels are shown in gray. Bolded TF names denote key regulators with high connectivity (connected components ≥ 5 nodes and degree > 3).

**Supplementary Figure 23.**
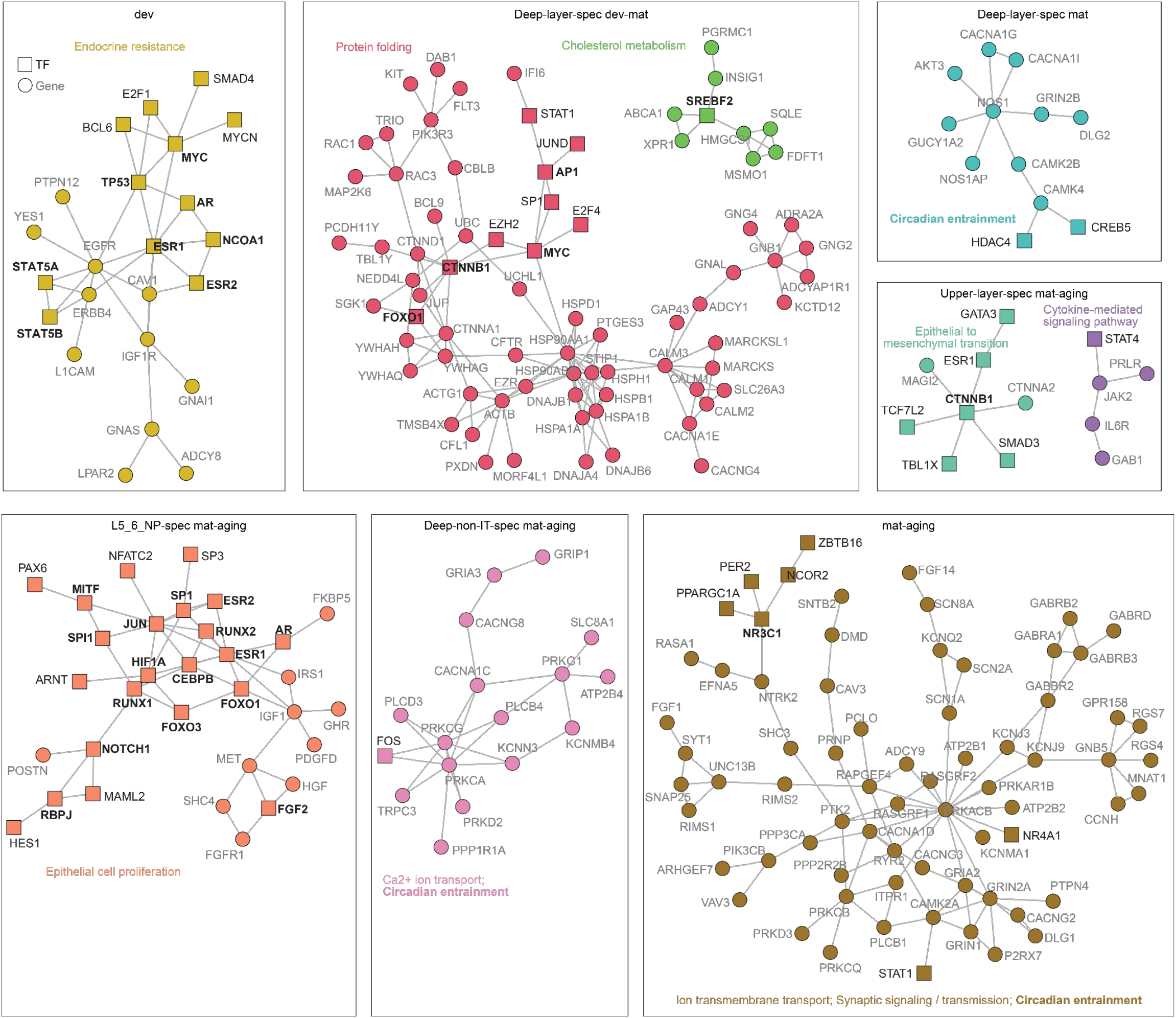
Regulatory networks for traDEG modules in excitatory neurons. TF-gene regulatory networks inferred for EN traDEG modules, with enriched biological processes annotated based on network composition. Squares indicate TFs; circles indicate target genes. TF labels are shown in black; gene labels are shown in gray. Bolded TF names denote key regulators with high connectivity (connected components ≥ 5 nodes and degree > 3).

**Supplementary Figure 24.**
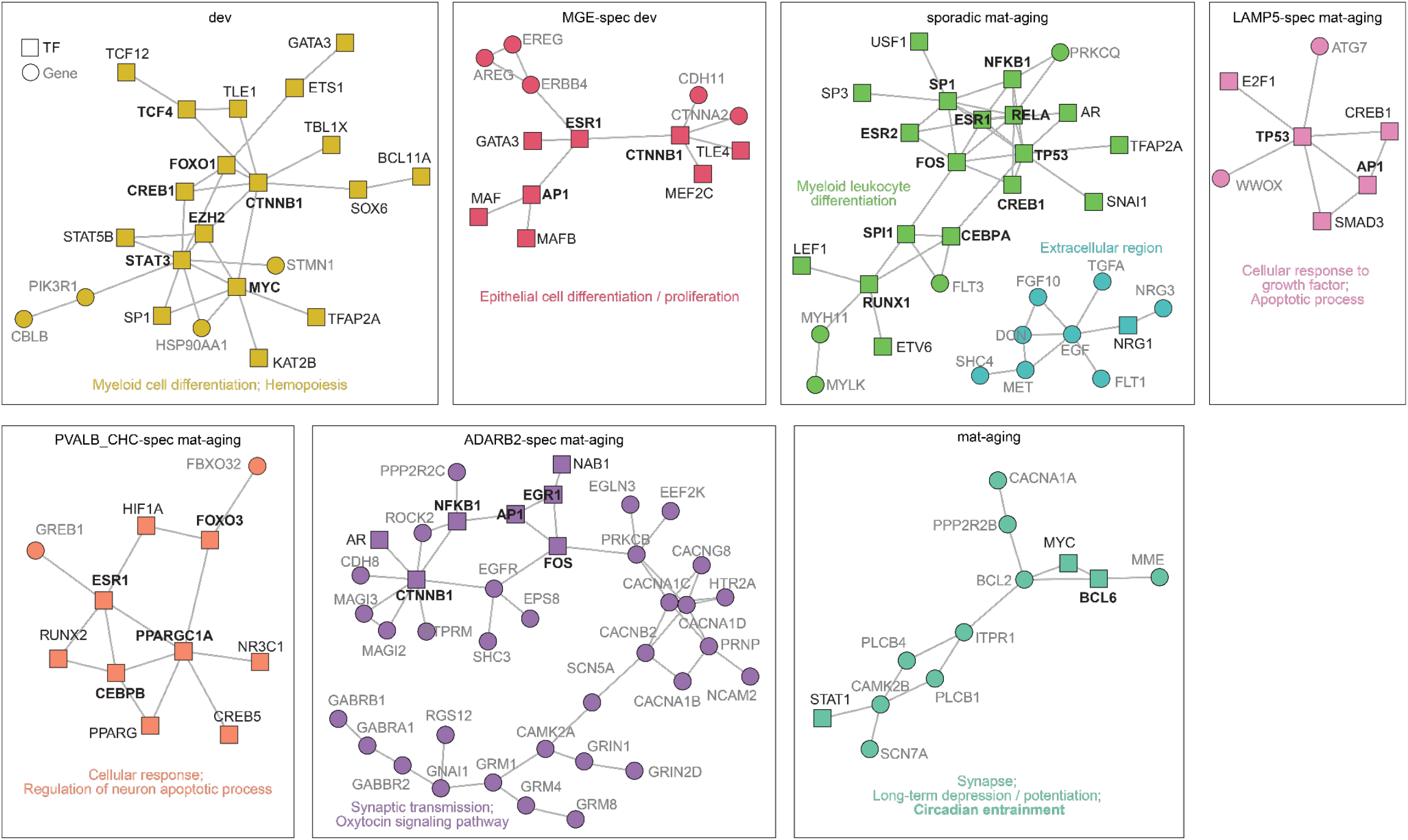
Regulatory networks for traDEG modules in inhibitory neurons. TF-gene regulatory networks inferred for IN traDEG modules, with enriched biological processes annotated based on network composition. Squares indicate TFs; circles indicate target genes. TF labels are shown in black; gene labels are shown in gray. Bolded TF names denote key regulators with high connectivity (connected components ≥ 5 nodes and degree > 3).

**Supplementary Figure 25.**
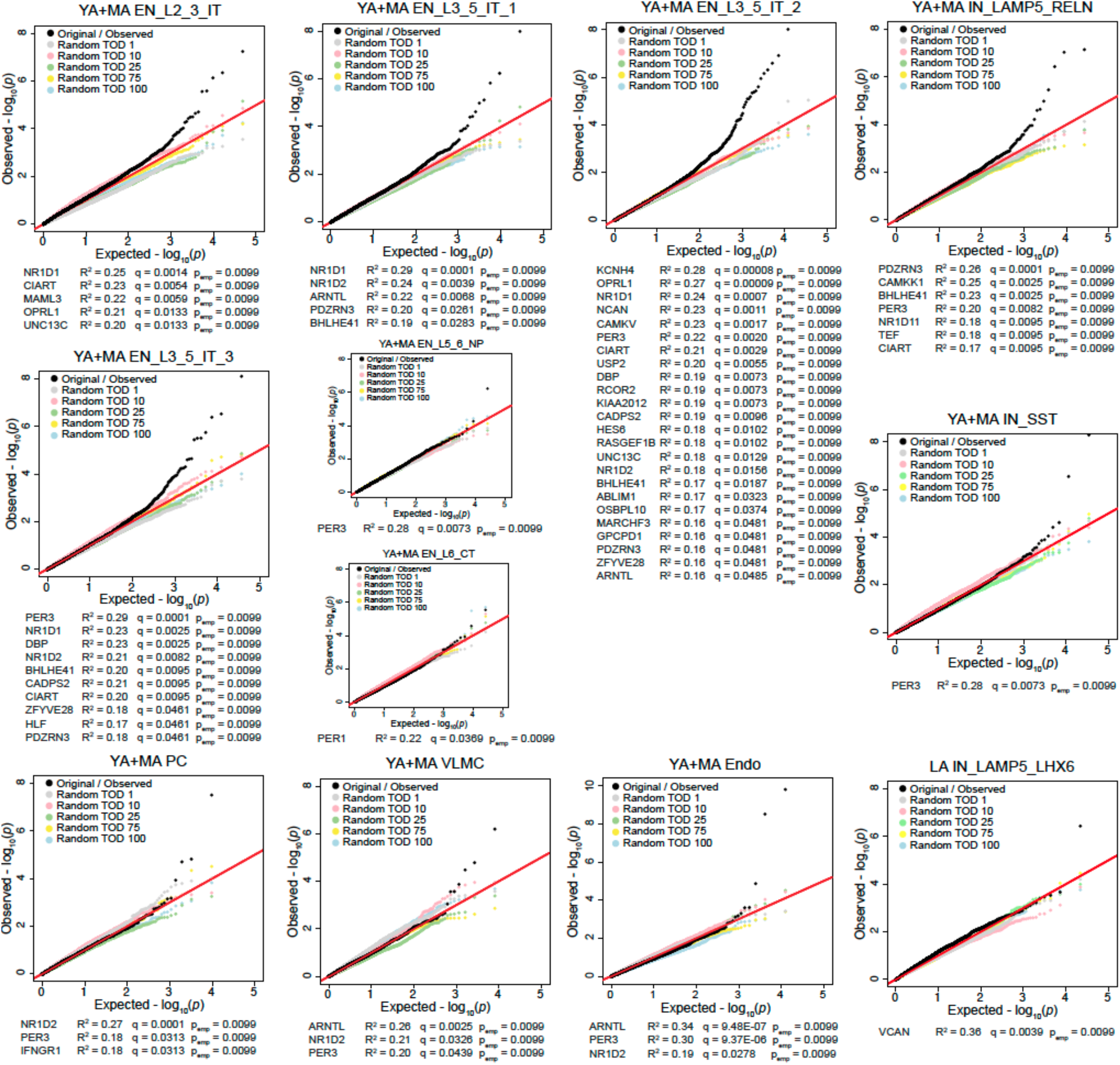
Q-Q Plots of representative rhythmicity analyses with randomized TOD. Q-Q Plots comparing the expected and observed p-value distributions of rhythmicity analyses. For each subclass, the original/observed distribution (black) is compared to 5 representative samples in which TOD was randomized. The subclasses shown had at least one gene that was significantly rhythmic (Adj.P.Val. ≤ 0.05) in the original/observed analysis. Below each Q-Q plot, the genes that were originally identified as rhythmic are listed, along with their goodness-of-fit (R2), adjusted p-value (q), and the final calculated empirical p-value.

**Supplementary Figure 26.**
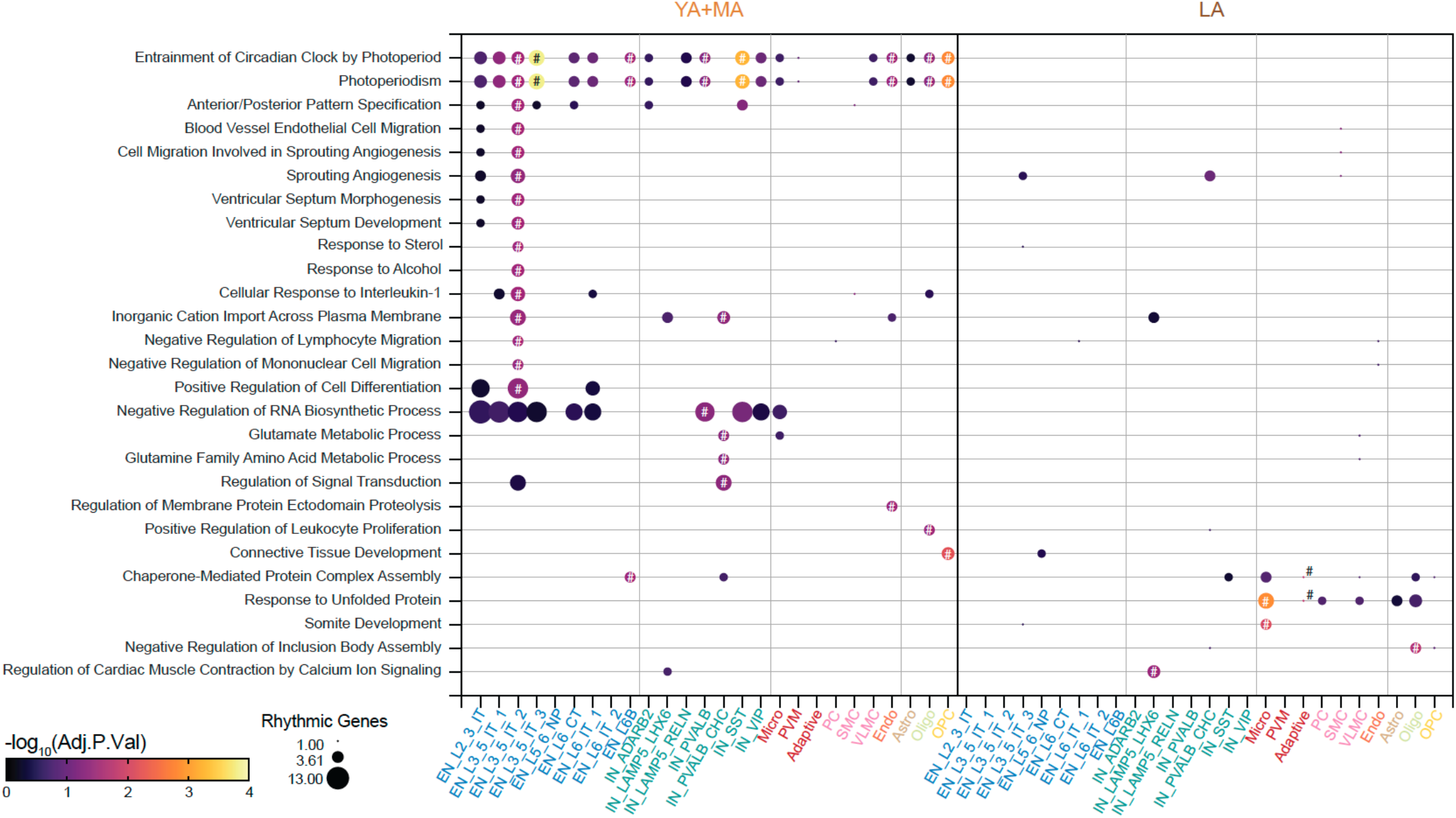
Pathways enriched in rhythmic genes. enrichR pathway analysis of rhythmic genes (p-value < 0.01) in YA+MA and LA subclasses. Pathways shown were identified as significantly enriched (adjusted p-value < 0.05, represented by #) in at least one subclass. Other points represent classes in which the pathway was only nominally significant (p < 0.01). The dashed box surrounds pathways that were driven by circadian clock genes.

**Supplementary Figure 27.**
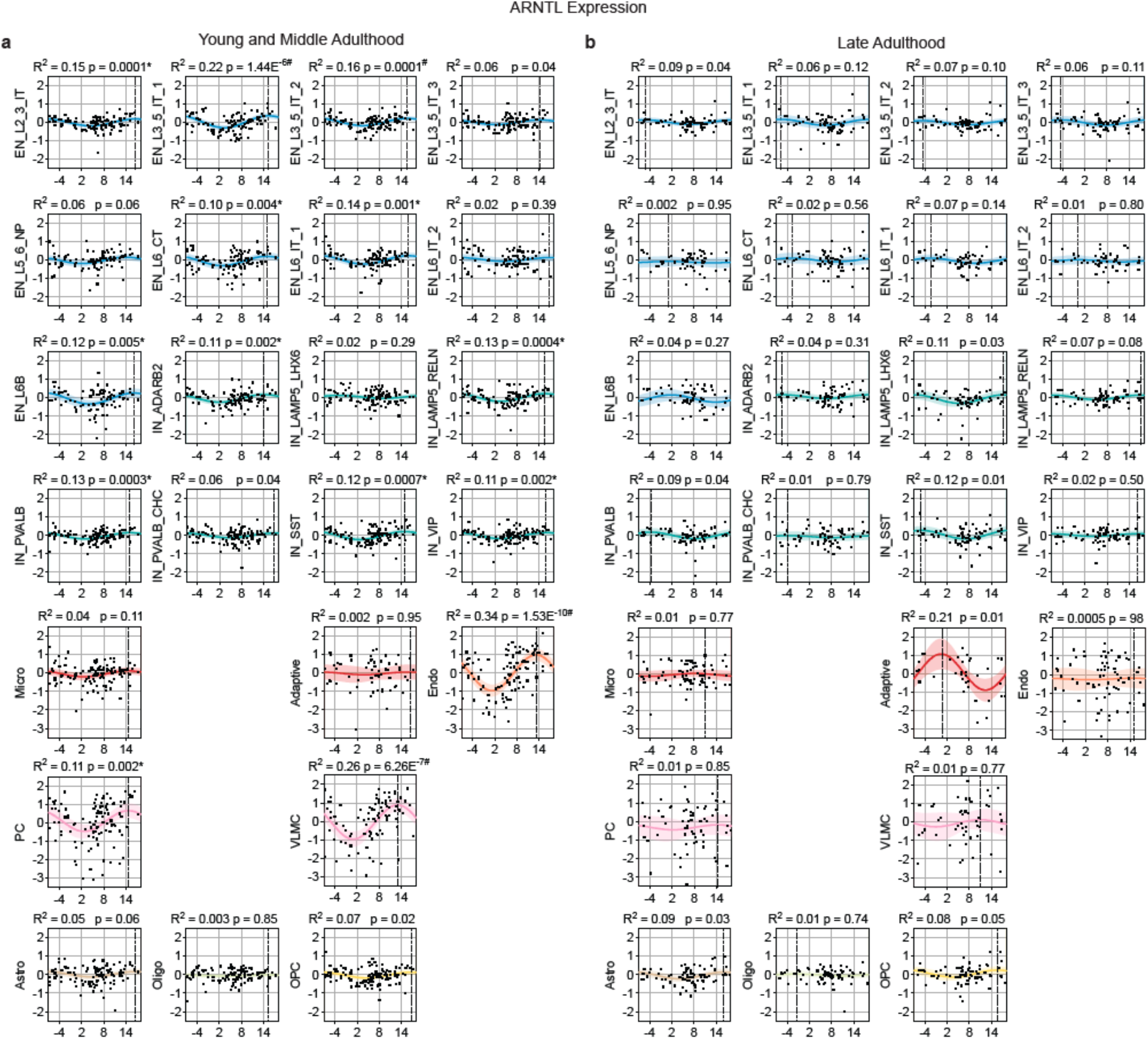
ARNTL rhythms. Plots of residualized gene expression across time of death (TOD) for the circadian gene ARNTL in **a**, YA+MA and **b**, LA subclasses. A dotted vertical line indicates the peak expression time for each curve.

**Supplementary Figure 28.**
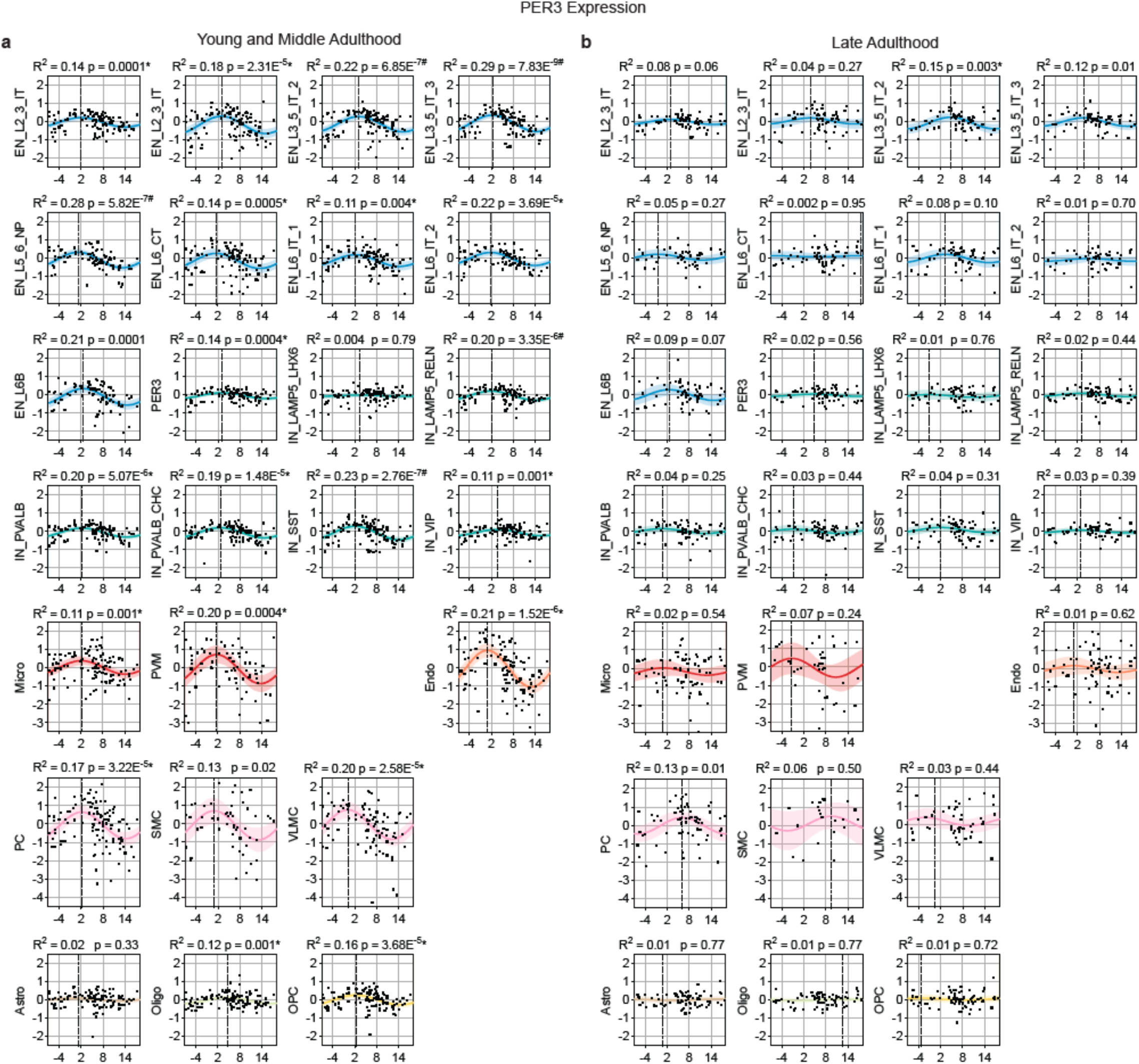
PER3 rhythms. Plots of residualized gene expression across time of death (TOD) for the circadian gene PER3 in **a**, YA+MA and **b**, LA subclasses. A dotted vertical line indicates the peak expression time for each curve.

### Captions for Supplementary table 1-4

**Supplementary Table 1** | Summary metadata of 284 donors ranges from 0 to 97 years.

**Supplementary Table 2** | List of GWAS traits and references.

**Supplementary Table 3** | Summary statistics of changes in nuclei composition using *crumblr* tool.

**Supplementary Table 4** | Tau scores of each gene across EN, IN and glia in four age groups.

### Captions for Supplementary Data 1-21

Supplementary Data can be accessed on synapse through accession code syn52396927

**Supplementary Data 1** | Metadata of 284 donors with neuropathology details.

**Supplementary Data 2** | Overall scDRS statistics from 1,307,674 nuclei and 284 donors for a subset of neurological, psychiatric, metabolic and immunological traits.

**Supplementary Data 3** | Variance explained by technical and biological covariates for each gene across 26 subclasses and four age groups.

**Supplemental Data 4** | Dreamlet age association summary statistics of genes for four age groups across 26 subclasses.

**Supplemental Data 5** | Functional pathway analysis of four age groups specific aDEGs across 26 subclasses.

**Supplemental Data 6** | Enrichment of brain and non-brain related risk genes from GWAS in four age groups specific aDEGs across 26 subclasses.

**Supplemental Data 7** | Lifespan age association summary statistics of each gene from a polynomial log model across 26 subclasses.

**Supplemental Data 8** | Functional pathway analysis of genes within each subclass across ten trajectories.

**Supplemental Data 9** | Enrichment of brain and non-brain related risk genes from GWAS in development and late adulthood aDEGs across ten trajectories.

**Supplemental Data 10** | Functional pathway analysis of development and late adulthood aDEGs across ten trajectories.

**Supplemental Data 11** | Composite *mashr* probabilities representing degree of sharing of a gene across 9 EN, 7 IN and 4 glial subclasses across four age groups.

**Supplemental Data 12** | Functional pathway analysis of shared genes across all cell classes and PPI cluster index in EN and IN from young and late adulthood, and in glia from late adulthood.

**Supplemental Data 13** | PPI clusters in EN and IN from young and late adulthood, and in glia from late adulthood from STRINGDB.

**Supplemental Data 14** | Clusters of traDEGs within Astro, Micro, OPC+Oligo, EN and IN lineages.

**Supplemental Data 15** | Pathway enrichment analyses of traDEG clusters within Astro, Micro, OPC+Oligo, EN and IN lineages.

**Supplemental Data 16** | Enrichment of brain and non-brain related risk genes from GWAS in traDEG clusters within Astro, Micro, OPC+Oligo, EN and IN lineages.

**Supplementary Data 17** | Rhythmic gene expression analyses.

**Supplementary Data 18** | Time of day randomization rhythmicity analyses

**Supplementary Data 19** | Gene expression rhythm parameter changes between YA+MA and LA within each subclass.

**Supplementary Data 20** | Gene expression rhythms - ΔR^2^ between YA+MA and LA within each subclass.

**Supplementary Data 21** | enrichR pathway analysis of rhythmic genes.

### Supplementary Notes

#### The PsychAD dataset

To enhance our ability to identify shared and distinct molecular pathways, as well as the causal variants and genes involved in various diseases of the brain, we generated a population-scale single-cell gene expression dataset in the postmortem human prefrontal cortex. The PsychAD cohort comprises 1,494 donors, each of which was subjected to single nucleus RNA-seq (snRNA-seq), generating more than 6.3 million individual nuclei (**Supplementary Fig. N1A**). Human brain specimens were obtained from three sources, Mount Sinai NIH Brain Bank and Tissue Repository (MSSM; 1,042 samples), NIMH-IRP Human Brain Collection Core (HBCC; 300 samples), and Rush Alzheimer’s Disease Center (RADC; 152 samples). For 1,381 (92%) donors, we also provide harmonized genotype data (either or both SNP array and whole genome sequencing). Importantly, less than 5% of the samples from either the MSSM^32^ or RADC^14,71^ cohort had previously been subjected to snRNA-seq. The PsychAD cohort consists of 1,074 donors affected by over 30 different disorders, including three represented by more than 100 cases (AD (*n* = 519), SCZ (*n* = 177), and DLBD (*n* = 112)) and three by at least 40 (vascular dementia (*n* = 85), BD (*n* = 72) and PD (*n* = 48)). In addition, the sample set also includes 420 neurotypical controls, as well as a number of cases with relatively understudied conditions, such as obsessive-compulsive disorder (*n* = 6), amyotrophic lateral sclerosis (*n* = 5), and progressive supranuclear palsy (*n* = 5). An important component of our data is the availability of phenotypic information on the nature and prevalence of neuropsychiatric symptoms (NPS). NPS frequently accompany AD and related dementias, and it has been estimated that, throughout the course of the disease, more than 80% of individuals will exhibit at least one NPS that significantly impacts their clinical outcomes^102^. So far, various studies have examined population data to characterize NPS along the AD continuum^103–105^. For example, depression and apathy are often the most observed symptoms in the early stages of AD, while delusions, hallucinations, and aggression become more prevalent as the disease advances^103^. However, research into the molecular basis of these NPS remains scarce, and we believe that our dataset provides a unique opportunity to explain their role in AD at a more granular level, leading to a better understanding of the broader disease and the potential identification of novel therapeutic targets.

To understand complex and heterogeneous human cortical tissues in a disease context, we first generated a robust cell type taxonomy that is invariant to the aging lifespan, disease phenotypes, and various sampling and technical biases. Following unified computational processing, quality controls, and batch normalization of more than 6.3 million individual nuclei, we used the cellular taxonomy of the primate DLPFC^106^ and human primary motor cortex^107^ as a baseline reference to annotate the cell types of the human DLPFC. The resulting cellular taxonomy was organized using three levels of taxonomic hierarchy, consisting of 8 broad cell classes, 27 subclasses, and 65 functional subtypes (**Supplementary Fig. N1B**). Each level of the hierarchy represents different slices of the clustering dendrogram. The top-level (class) defines 8 major cell types, including two broad neuronal cell types: glutamatergic excitatory (EN) and GABAergic inhibitory neurons (IN), three glial (Astrocyte, Oligodendrocyte, OPC), and three non-neuronal cell types (Immune, Mural, Endothelial). Subsequent levels (subclass and subtype) were derived by iteratively re-clustering the subset of cells by gene matrix using a new set of variable genes relevant to the particular cell type. For example, the various EN subclasses were distinguished by their laminar organization and axon projection characteristics (IT: intra-telencephalic, ET: extra-telencephalic, NP: near projecting, CT: corticothalamic, and L6B), while IN subclasses were denoted using characteristic marker genes. This approach allowed us to identify 10 subclasses of EN and 7 subclasses of IN. The 10 EN subclasses were further subdivided into 18 functionally distinct subtypes, while the 7 IN subclasses comprised 21 subtypes.

The release of this dataset by the PsychAD consortium is accompanied by a series of manuscripts that are submitted as a package to Nature Portfolio journals describing the cross-disorder analysis of transcriptomic vulnerability (Lee *et al*, Single-cell atlas of transcriptomic vulnerability across multiple neurodegenerative and neuropsychiatric diseases), genetic regulation of gene expression (Zeng *et al*, Single nucleus, multi-ancestry atlas of genetic regulation of gene expression in the human brain), and transcriptome-wide association studies (Venkatesh *et al*, Single-nucleus transcriptome-wide association study of human brain disorders). The consortium has also leveraged neurotypical controls to assemble a map of transcriptomic changes across the lifespan (Yang *et al*, A lifespan transcriptomic atlas of the human prefrontal cortex at single-cell resolution). Also, we performed two advanced analyses using emerging machine learning and AI approaches to detect phenotype-associated cells revealing potential novel cell subpopulations and expressed genes (PASCode: He *et al*, Phenotype Scoring of Population Scale Single-Cell Data Dissects Alzheimer’s Disease Complexity) and quantify personalized importance scores of genes, cell types and regulatory networks for various PsychAD phenotypes (iBrainMAP: Chandrashekar *et al*, Single-cell transcriptomic dissection of 1,494 human brains reveals personalized functional genomics for Alzheimer’s disease phenotypes). Furthermore, the computational scale and diversity of the generated data led to the development of several analytical tools, including *Dreamlet* for differential gene expression^28^ and *Crumblr* for the identification of cell types with significant shifts in cell type proportions between disease cases and controls (Hoffman *et al*, Fast, flexible analysis of compositional data with crumblr). Lastly, we have prepared a separate manuscript (Fullard *et al*, Population-scale cross-disorder atlas of the human prefrontal cortex at single-cell resolution) that offers a comprehensive overview of the clinical and demographic donor information, as well as detailed descriptions of the techniques used in snRNA-seq and SNP array sample preparation, including bioinformatics preprocessing and quality control methods applied to the resulting data. The PsychAD dataset is publicly available at the AMP-AD Knowledge Portal in the study-specific folder https://doi.org/10.7303/syn60084804. This repository includes sample metadata, as well as raw and processed sequencing data for snRNA-seq and genotyping. snRNA-seq data can be inspected using the CELLxGENE visualization tool at https://cellxgene.cziscience.com/collections/84ce6837-548d-4a1f-919f-0bc0d9a3952f.

**Supplementary Figure N1.**
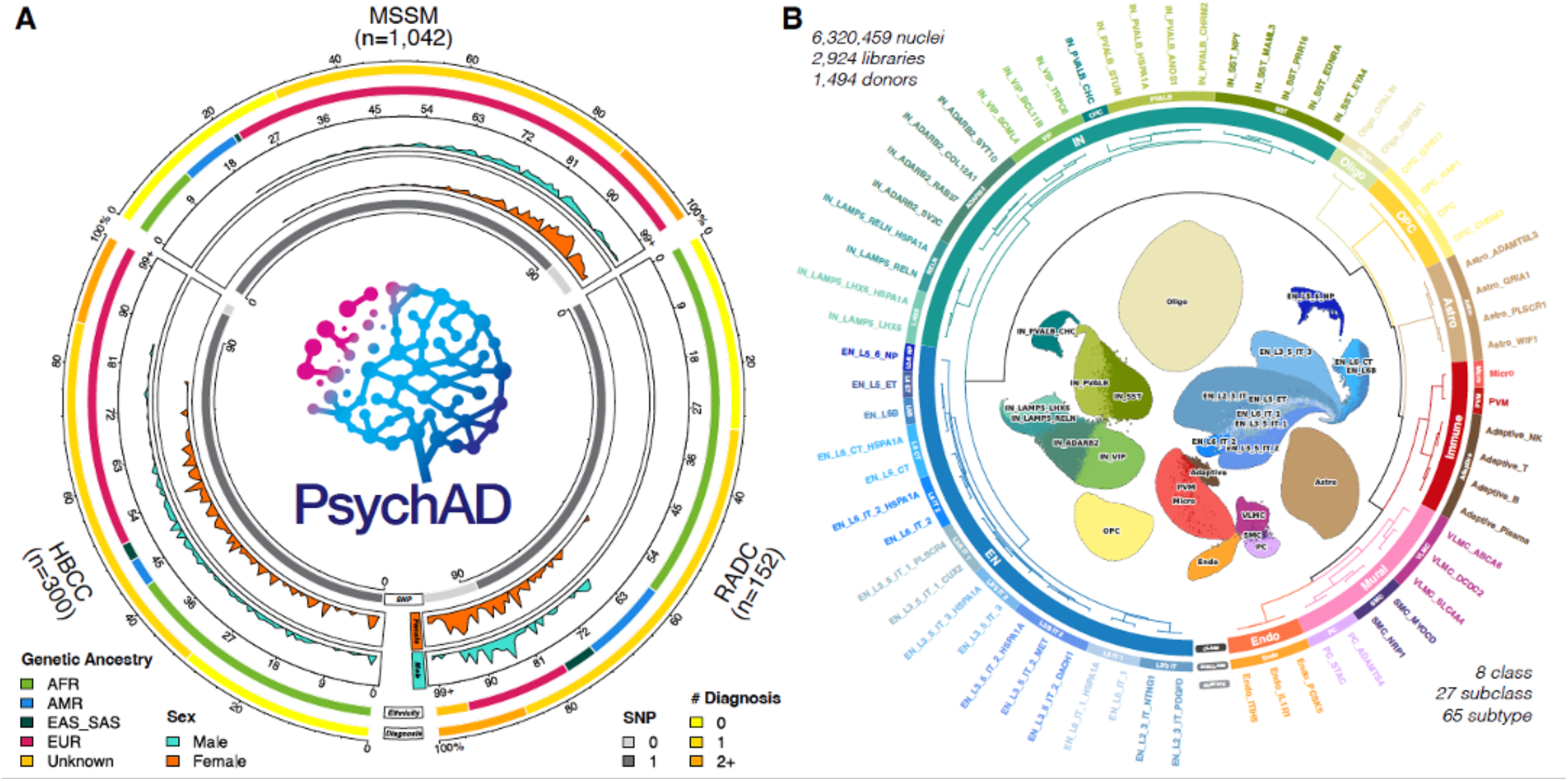
The PsychAD cohort. **A**, Organization of the PsychAD study cohort. Breakdown of brain donors by tissue repository, number of neurological diagnoses, genetic ancestry, age, sex, and availability of genotype data. **B**, Unified processing of the single-cell transcriptomics atlas and hierarchical structure of cellular taxonomy of the overall transcriptome. Taxonomic annotation of cellular phenotype at three levels of granularity; class (*n* = 8), subclass (*n* = 27), and subtype (*n* = 65).

## References

1. Nejati, V., Majdi, R., Salehinejad, M. A. & Nitsche, M. A. The role of dorsolateral and ventromedial prefrontal cortex in the processing of emotional dimensions. Sci. Rep. 11, 1971 (2021).

2. Barbey, A. K., Koenigs, M. & Grafman, J. Dorsolateral prefrontal contributions to human working memory. Cortex 49, 1195–1205 (2013).

3. Hertrich, I., Dietrich, S., Blum, C. & Ackermann, H. The Role of the Dorsolateral Prefrontal Cortex for Speech and Language Processing. Front. Hum. Neurosci. 15, 645209 (2021).

4. Smucny, J., Dienel, S. J., Lewis, D. A. & Carter, C. S. Mechanisms underlying dorsolateral prefrontal cortex contributions to cognitive dysfunction in schizophrenia. Neuropsychopharmacology 47, 292–308 (2022).

5. Girdhar, K. et al. Chromatin domain alterations linked to 3D genome organization in a large cohort of schizophrenia and bipolar disorder brains. Nat. Neurosci. 25, 474–483 (2022).

6. Pizzagalli, D. A. & Roberts, A. C. Prefrontal cortex and depression. Neuropsychopharmacology 47, 225–246 (2022).

7. Cain, A. et al. Multicellular communities are perturbed in the aging human brain and Alzheimer’s disease. Nat. Neurosci. 26, 1267–1280 (2023).

8. Velmeshev, D. et al. Single-cell analysis of prenatal and postnatal human cortical development. Science 382, eadf0834 (2023).

9. Zeng, B. et al. The single-cell and spatial transcriptional landscape of human gastrulation and early brain development. Cell Stem Cell 30, 851–866.e7 (2023).

10. Eze, U. C., Bhaduri, A., Haeussler, M., Nowakowski, T. J. & Kriegstein, A. R. Single-cell atlas of early human brain development highlights heterogeneity of human neuroepithelial cells and early radial glia. Nat. Neurosci. 24, 584–594 (2021).

11. Herring, C. A. et al. Human prefrontal cortex gene regulatory dynamics from gestation to adulthood at single-cell resolution. Cell 185, 4428–4447.e28 (2022).

12. Fullard, J. F. et al. Population-scale cross-disorder atlas of the human prefrontal cortex at single-cell resolution.

13. Lee, D. et al. Single-cell atlas of transcriptomic vulnerability across multiple neurodegenerative and neuropsychiatric diseases.

14. Mathys, H. et al. Single-cell atlas reveals correlates of high cognitive function, dementia, and resilience to Alzheimer’s disease pathology. Cell 186, 4365–4385.e27 (2023).

15. Hoffman, G. E. & Schadt, E. E. variancePartition: interpreting drivers of variation in complex gene expression studies. BMC Bioinformatics 17, 483 (2016).

16. Herring, C. A. et al. Human prefrontal cortex gene regulatory dynamics from gestation to adulthood at single-cell resolution. Cell 185, 4428–4447.e28 (2022).

17. Ham, S. & Lee, S.-J. V. Advances in transcriptome analysis of human brain aging. Exp. Mol. Med. 52, 1787–1797 (2020).

18. Chaudhari, P. R., Singla, A. & Vaidya, V. A. Early Adversity and Accelerated Brain Aging: A Mini-Review. Front. Mol. Neurosci. 15, 822917 (2022).

19. Chiou, K. L. et al. Multiregion transcriptomic profiling of the primate brain reveals signatures of aging and the social environment. Nat. Neurosci. 25, 1714–1723 (2022).

20. Muraoka, S. et al. Enrichment of Neurodegenerative Microglia Signature in Brain-Derived Extracellular Vesicles Isolated from Alzheimer’s Disease Mouse Models. J. Proteome Res. 20, 1733–1743 (2021).

21. Felsky, D. et al. Polygenic analysis of inflammatory disease variants and effects on microglia in the aging brain. Mol. Neurodegener. 13, 38 (2018).

22. Bahrami, S. et al. Shared Genetic Loci Between Body Mass Index and Major Psychiatric Disorders: A Genome-wide Association Study. JAMA Psychiatry 77, 503–512 (2020).

23. Marcus, M. D. & Wildes, J. E. Obesity: is it a mental disorder? Int. J. Eat. Disord. 42, 739–753 (2009).

24. Rajan, T. M. & Menon, V. Psychiatric disorders and obesity: A review of association studies. J. Postgrad. Med. 63, 182–190 (2017).

25. Leutner, M. et al. Obesity as pleiotropic risk state for metabolic and mental health throughout life. Transl. Psychiatry 13, 1–12 (2023).

26. Stiles, J. & Jernigan, T. L. The basics of brain development. Neuropsychol. Rev. 20, 327–348 (2010).

27. Count ratio uncertainty modeling base linear regression. https://diseaseneurogenomics.github.io/crumblr/.

28. Hoffman, G. E. et al. Efficient differential expression analysis of large-scale single cell transcriptomics data using dreamlet. bioRxiv 2023.03.17.533005 (2023) doi:10.1101/2023.03.17.533005.

29. Ling, E. et al. A concerted neuron-astrocyte program declines in ageing and schizophrenia. Nature 627, 604–611 (2024).

30. Emani, P. S. et al. Single-cell genomics and regulatory networks for 388 human brains. Science 384, eadi5199 (2024).

31. El Marroun, H. et al. Prenatal exposure to maternal and paternal depressive symptoms and brain morphology: A population-based prospective neuroimaging study in young children. Depress. Anxiety 33, 658–666 (2016).

32. Ruzicka, W. B. et al. Single-cell multi-cohort dissection of the schizophrenia transcriptome. Science 384, eadg5136 (2024).

33. Karczewski, K. J. et al. The mutational constraint spectrum quantified from variation in 141,456 humans. Nature 581, 434–443 (2020).

34. Lynch, G., Rex, C. S. & Gall, C. M. Synaptic plasticity in early aging. Ageing Res. Rev. 5, 255–280 (2006).

35. Arcos-Burgos, M., Lopera, F., Sepulveda-Falla, D. & Mastronardi, C. Neural Plasticity during Aging. Neural Plast. 2019, 6042132 (2019).

36. Izgi, H. et al. Inter-tissue convergence of gene expression during ageing suggests age-related loss of tissue and cellular identity. Elife 11, e68048 (2022).

37. Jin, K. et al. Brain-wide cell-type-specific transcriptomic signatures of healthy ageing in mice. Nature 638, 182–196 (2025).

38. Urbut, S. M., Wang, G., Carbonetto, P. & Stephens, M. Flexible statistical methods for estimating and testing effects in genomic studies with multiple conditions. Nat. Genet. 51, 187–195 (2019).

39. Lüleci, H. B. & Yılmaz, A. Robust and rigorous identification of tissue-specific genes by statistically extending tau score. BioData Min. 15, 31 (2022).

40. Szklarczyk, D. et al. The STRING database in 2023: protein–protein association networks and functional enrichment analyses for any sequenced genome of interest. Nucleic Acids Res. 51, D638–D646 (2022).

41. Molofsky, A. V. et al. Astrocytes and disease: a neurodevelopmental perspective. Genes Dev. 26, 891–907 (2012).

42. Köhler, S., Winkler, U. & Hirrlinger, J. Heterogeneity of astrocytes in grey and white matter. Neurochem. Res. 46, 3–14 (2021).

43. Lee, H.-G., Wheeler, M. A. & Quintana, F. J. Function and therapeutic value of astrocytes in neurological diseases. Nat. Rev. Drug Discov. 21, 339–358 (2022).

44. Kruyer, A., Kalivas, P. W. & Scofield, M. D. Astrocyte regulation of synaptic signaling in psychiatric disorders. Neuropsychopharmacology 48, 21–36 (2023).

45. Allen, N. J. & Lyons, D. A. Glia as architects of central nervous system formation and function. Science 362, 181–185 (2018).

46. Sanmarco, L. M. et al. Gut-licensed IFNγ+ NK cells drive LAMP1+TRAIL+ anti-inflammatory astrocytes. Nature 590, 473–479 (2021).

47. Linnerbauer, M., Wheeler, M. A. & Quintana, F. J. Astrocyte crosstalk in CNS inflammation. Neuron 108, 608–622 (2020).

48. Wheeler, M. A. et al. MAFG-driven astrocytes promote CNS inflammation. Nature 578, 593–599 (2020).

49. Romanos, J., Benke, D., Pietrobon, D., Zeilhofer, H. U. & Santello, M. Astrocyte dysfunction increases cortical dendritic excitability and promotes cranial pain in familial migraine. Sci. Adv. 6, eaaz1584 (2020).

50. Sofroniew, M. V. & Vinters, H. V. Astrocytes: biology and pathology. Acta Neuropathol. 119, 7–35 (2010).

51. Badia-I-Mompel, P. et al. decoupleR: ensemble of computational methods to infer biological activities from omics data. Bioinform. Adv. 2, vbac016 (2022).

52. Wang, L. et al. Molecular and cellular dynamics of the developing human neocortex. Nature 1–10 (2025).

53. Huuki-Myers, L. A. et al. A data-driven single-cell and spatial transcriptomic map of the human prefrontal cortex. Science 384, eadh1938 (2024).

54. Chen, X. et al. Microglia-mediated T cell infiltration drives neurodegeneration in tauopathy. Nature 615, 668–677 (2023).

55. Bartels, T., De Schepper, S. & Hong, S. Microglia modulate neurodegeneration in Alzheimer’s and Parkinson’s diseases. Science 370, 66–69 (2020).

56. Bellenguez, C. et al. New insights into the genetic etiology of Alzheimer’s disease and related dementias. Nat. Genet. 54, 412–436 (2022).

57. Greig, L. C., Woodworth, M. B., Galazo, M. J., Padmanabhan, H. & Macklis, J. D. Molecular logic of neocortical projection neuron specification, development and diversity. Nat. Rev. Neurosci. 14, 755–769 (2013).

58. Silbereis, J. C., Pochareddy, S., Zhu, Y., Li, M. & Sestan, N. The cellular and molecular landscapes of the developing human central nervous system. Neuron 89, 248–268 (2016).

59. Südhof, T. C. The cell biology of synapse formation. J. Cell Biol. 220, (2021).

60. Chen, M. B., Jiang, X., Quake, S. R. & Südhof, T. C. Persistent transcriptional programmes are associated with remote memory. Nature 587, 437–442 (2020).

61. Zhu, K. et al. Multi-omic profiling of the developing human cerebral cortex at the single-cell level. Sci. Adv. 9, eadg3754 (2023).

62. Forsyth, J. K. & Lewis, D. A. Mapping the consequences of impaired synaptic plasticity in schizophrenia through development: An integrative model for diverse clinical features. Trends Cogn. Sci. 21, 760–778 (2017).

63. Ma, Y. et al. Activity-Dependent Transcriptional Program in NGN2+ Neurons Enriched for Genetic Risk for Brain-Related Disorders. Biol. Psychiatry 95, 187–198 (2024).

64. Johnson, K. A. et al. Tourette syndrome: clinical features, pathophysiology, and treatment. Lancet Neurol. 22, 147–158 (2023).

65. Lowe, C. J., Reichelt, A. C. & Hall, P. A. The prefrontal cortex and obesity: A health neuroscience perspective. Trends Cogn. Sci. 23, 349–361 (2019).

66. Stowe, T. A. & McClung, C. A. How Does Chronobiology Contribute to the Development of Diseases in Later Life. Clin. Interv. Aging 18, 655–666 (2023).

67. Wolff, C. A. et al. Defining the age-dependent and tissue-specific circadian transcriptome in male mice. Cell Rep. 42, 111982 (2023).

68. Uddin, M. S., Yu, W. S. & Lim, L. W. Exploring ER stress response in cellular aging and neuroinflammation in Alzheimer’s disease. Ageing Res. Rev. 70, 101417 (2021).

69. Kim, Y. H. & Lazar, M. A. Transcriptional Control of Circadian Rhythms and Metabolism: A Matter of Time and Space. Endocr. Rev. 41, 707–732 (2020).

70. Li, M. et al. Integrative functional genomic analysis of human brain development and neuropsychiatric risks. Science 362, eaat7615 (2018).

71. Green, G. S. et al. Cellular dynamics across aged human brains uncover a multicellular cascade leading to Alzheimer’s disease. bioRxiv (2023) doi:10.1101/2023.03.07.531493.

72. Su, Y. et al. A single-cell transcriptome atlas of glial diversity in the human hippocampus across the postnatal lifespan. Cell Stem Cell 29, 1594–1610.e8 (2022).

73. Nowakowski, T. J. et al. Spatiotemporal gene expression trajectories reveal developmental hierarchies of the human cortex. Science 358, 1318–1323 (2017).

74. Reid, D. A. et al. Incorporation of a nucleoside analog maps genome repair sites in postmitotic human neurons. Science 372, 91–94 (2021).

75. Li, H., Wang, Z., Ma, T., Wei, G. & Ni, T. Alternative splicing in aging and age-related diseases. Translational Medicine of Aging 1, 32–40 (2017).

76. Lu, T. et al. Gene regulation and DNA damage in the ageing human brain. Nature 429, 883–891 (2004).

77. Bethlehem, R. A. I. et al. Brain charts for the human lifespan. Nature 604, 525–533 (2022).

78. Khodosevich, K. & Sellgren, C. M. Neurodevelopmental disorders-high-resolution rethinking of disease modeling. Mol. Psychiatry 28, 34–43 (2023).

79. Kang, H. J. et al. Spatio-temporal transcriptome of the human brain. Nature 478, 483–489 (2011).

80. Chen, C.-Y. et al. Effects of aging on circadian patterns of gene expression in the human prefrontal cortex. Proc. Natl. Acad. Sci. U. S. A. 113, 206–211 (2016).

81. Stoeckius, M. et al. Cell Hashing with barcoded antibodies enables multiplexing and doublet detection for single cell genomics. Genome Biol. 19, 224 (2018).

82. Kaminow, B., Yunusov, D. & Dobin, A. STARsolo: accurate, fast and versatile mapping/quantification of single-cell and single-nucleus RNA-seq data. bioRxiv 2021.05.05.442755 (2021) doi:10.1101/2021.05.05.442755.

83. Huang, X. & Huang, Y. Cellsnp-lite: an efficient tool for genotyping single cells. Bioinformatics (2021) doi:10.1093/bioinformatics/btab358.

84. Huang, Y., McCarthy, D. J. & Stegle, O. Vireo: Bayesian demultiplexing of pooled single-cell RNA-seq data without genotype reference. Genome Biol. 20, 273 (2019).

85. Fort, A. et al. MBV: a method to solve sample mislabeling and detect technical bias in large combined genotype and sequencing assay datasets. Bioinformatics 33, 1895–1897 (2017).

86. Li, B. et al. Cumulus provides cloud-based data analysis for large-scale single-cell and single-nucleus RNA-seq. Nat. Methods 17, 793–798 (2020).

87. Wolf, F. A., Angerer, P. & Theis, F. J. SCANPY: large-scale single-cell gene expression data analysis. Genome Biol. 19, 15 (2018).

88. Fleming, S. J. et al. Unsupervised removal of systematic background noise from droplet-based single-cell experiments using CellBender. Nat. Methods 20, 1323–1335 (2023).

89. Wolock, S. L., Lopez, R. & Klein, A. M. Scrublet: Computational Identification of Cell Doublets in Single-Cell Transcriptomic Data. Cell Syst 8, 281–291.e9 (2019).

90. Korsunsky, I. et al. Fast, sensitive and accurate integration of single-cell data with Harmony. Nat. Methods 16, 1289–1296 (2019).

91. Xu, C. et al. Probabilistic harmonization and annotation of single-cell transcriptomics data with deep generative models. Mol. Syst. Biol. 17, e9620 (2021).

92. Mathys, H. et al. Single-cell transcriptomic analysis of Alzheimer’s disease. Nature 570, 332–337 (2019).

93. Becht, E. et al. Dimensionality reduction for visualizing single-cell data using UMAP. Nat. Biotechnol. 37, 38–44 (2018).

94. de Leeuw, C. A., Mooij, J. M., Heskes, T. & Posthuma, D. MAGMA: generalized gene-set analysis of GWAS data. PLoS Comput. Biol. 11, e1004219 (2015).

95. Gayoso, A. et al. A Python library for probabilistic analysis of single-cell omics data. Nat. Biotechnol. 40, 163–166 (2022).

96. Pardo, B. et al. spatialLIBD: an R/Bioconductor package to visualize spatially-resolved transcriptomics data. BMC Genomics 23, 434 (2022).

97. 1000 Genomes Project Consortium et al. A global reference for human genetic variation. Nature 526, 68–74 (2015).

98. Müller-Dott, S. et al. Expanding the coverage of regulons from high-confidence prior knowledge for accurate estimation of transcription factor activities. Nucleic Acids Res. 51, 10934–10949 (2023).

99. Seney, M. L. et al. Diurnal rhythms in gene expression in the prefrontal cortex in schizophrenia. Nat. Commun. 10, 3355 (2019).

100. Xue, X. et al. DiffCircaPipeline: a framework for multifaceted characterization of differential rhythmicity. Bioinformatics 39, (2023).

101. Zhang, M. J. et al. Polygenic enrichment distinguishes disease associations of individual cells in single-cell RNA-seq data. Nat. Genet. 54, 1572–1580 (2022).

102. Lyketsos, C. G. et al. Prevalence of neuropsychiatric symptoms in dementia and mild cognitive impairment: results from the cardiovascular health study. JAMA 288, 1475–1483 (2002).

103. Lyketsos, C. G. et al. Neuropsychiatric symptoms in Alzheimer’s disease. Alzheimers. Dement. 7, 532–539 (2011).

104. Zhao, Q.-F. et al. The prevalence of neuropsychiatric symptoms in Alzheimer’s disease: Systematic review and metaanalysis. J. Affect. Disord. 190, 264–271 (2016).

105. Pless, A. et al. Understanding neuropsychiatric symptoms in Alzheimer’s disease: challenges and advances in diagnosis and treatment. Front. Neurosci. 17, 1263771 (2023).

106. Ma, S. et al. Molecular and cellular evolution of the primate dorsolateral prefrontal cortex. Science 377, eabo7257 (2022).

107. BRAIN Initiative Cell Census Network (BICCN). A multimodal cell census and atlas of the mammalian primary motor cortex. Nature 598, 86–102 (2021).

